# Plasma Microbial Cell-free DNA Sequencing from Over 15,000 Patients Identified a Broad Spectrum of Pathogens

**DOI:** 10.1101/2023.01.03.22283605

**Authors:** Sarah Y. Park, Eliza J Chang, Nathan Ledeboer, Kevin Messacar, Martin S. Lindner, Shivkumar Venkatasubrahmanyam, Sivan Bercovici, Judith C. Wilber, Marla Lay Vaughn, Bradley A. Perkins, Frederick S. Nolte

**Affiliations:** Karius, Redwood City, CA; Medical College of Wisconsin, Milwaukee, WI; University of Colorado, Children’s Hospital Colorado, Aurora, CO.

**Keywords:** microbial cell-free DNA, high-throughput nucleic acid sequencing, liquid biopsy for infectious diseases, metagenomics

## Abstract

Microbial cell-free DNA (mcfDNA) sequencing is an emerging infectious disease diagnostic tool which enables unbiased pathogen detection from plasma. The Karius Test®, a commercial mcfDNA sequencing assay developed by and available since 2017 from Karius, Inc. (Redwood City, CA), detects and quantifies mcfDNA as molecules/μl in plasma. The commercial sample data and results for all tests conducted from April 2018 through mid-September 2021 were evaluated for laboratory performance metrics, reported pathogens, and data from test requisition forms. A total of 18,690 reports were generated from 15,165 patients in a hospital setting among 39 states and the District of Columbia. The median time from sample receipt to reported result was 26 hours (IQR 25–28), and 96% of samples had valid test results. Almost two-thirds (65%) of patients were adults, and 29% at the time of diagnostic testing had ICD10 codes representing a diverse array of clinical scenarios. There were 10,752 (58%) reports that yielded at least one taxon for a total of 22,792 detections spanning 701 unique microbial taxa. The 50 most common taxa detected included 36 bacteria, 9 viruses, and 5 fungi. Opportunistic fungi (374 *Aspergillus* spp., 258 *Pneumocystis jirovecii*, 196 *Mucorales*, and 33 dematiaceous fungi) comprised 861 (4%) of all detections. Additional diagnostically challenging pathogens (247 zoonotic and vector borne pathogens, 144 *Mycobacteria*, 80 *Legionella* spp., 78 systemic dimorphic fungi, 69 *Nocardia* spp., and 57 protozoan parasites) comprised 675 (3%) of all detections. We report the largest cohort of patients tested using plasma mcfDNA sequencing. The wide variety of pathogens detected by plasma mcfDNA sequencing reaffirm our understanding of the ubiquity of some infections while also identifying taxa less commonly detected by conventional methods.

## INTRODUCTION

Sequencing microbial cell-free DNA (mcfDNA) in plasma represents integration of progress in genomic sequencing, computation analyses, and recognition of cell-free DNA as a clinically useful blood analyte (1–3). Over the last four decades, PCR-based tests, specifically multiplexed broad syndromic panels, have made welcomed contributions to infectious disease diagnostics but fall short of desired performance including breadth of pathogen detection and require samples of infected tissue or body fluid (4). Broad range PCR testing arguably facilitates considering a wider range of potential pathogens but is still only limited to bacteria and fungi (5). A recent meta-analysis, which included 20 studies that satisfied the Quality Assessment of Diagnostic Accuracy Studies (6) to assess the diagnostic accuracy of next generation sequencing in distinguishing infectious diseases, concluded that this group of technologies demonstrated satisfactory diagnostic performance for infections and yielded an overall detection rate superior to conventional methods (7). Four of the studies included in this review employed plasma mcfDNA sequencing. Moreover, early experience with plasma mcfDNA sequencing suggests this new approach, especially when applied early in a patient’s clinical course and for specific use cases, has potential to improve upon the above-noted shortcomings (8, 9). Plasma mcfDNA sequencing enables unbiased pathogen detection through noninvasive sampling with rapid turnaround, creating opportunities to enhance diagnosis of bloodstream and deep-seated infections (10–12). This is urgently needed particularly among immunocompromised patients who are often the most vulnerable to serious and frequently life-threatening infections.

The Karius Test® is an analytically and clinically validated mcfDNA sequencing test, commercially available for US inpatients since 2017 as a laboratory developed test from Karius, Inc. The test can identify and quantitate molecules/µl (MPM) mcfDNA in plasma from >1,500 bacteria, DNA viruses, fungi, and parasites. The analytical and clinical validation of the test was previously reported (12). Since the time of this study, others have reported how this unbiased test may contribute to the diagnosis and management of life-threatening infections in defined patient populations, specifically immunocompromised patients early in their clinical course, by creating the potential to minimize invasive procedures (13), reducing time to specific etiologic diagnosis of infections compared with standard of care (SOC) microbiological testing (14), and, in some cases, optimizing antimicrobial therapy (15, 16). In contrast, several retrospective, observational reviews of Karius Test utilization concluded that in routine clinical practice the diagnostic and clinical impact of the test was limited, which highlights the need for diagnostic stewardship to optimize implementation and maximize clinical utility in specific patient populations (17–19).

Plasma mcfDNA sequencing for infectious disease diagnosis performance at scale with respect to time to results, quality metrics, positivity rates, and diversity of taxa detected has not been previously reported. Here we review the results for a large commercial laboratory testing cohort of over 18,000 plasma samples from over 15,000 patients in a hospital setting with the primary objective to provide additional insights about the breadth and depth of microbial identifications. In the course of doing so, we also describe the current test performance metrics and characterize clinical use based on the limited available data.

## MATERIAL AND METHODS

### Commercial laboratory test cohort

The Karius Test results for patients from across the United States were evaluated for reported pathogens and patient data (including basic demographics, ordering clinician, and ICD-10 codes if provided) obtained from the test request forms (TRF) for all samples tested from April 1, 2018 through mid-September, 2021. Laboratory performance metrics were gathered for all samples collected from April 1, 2018 through the end of September, 2021. Diagnosis codes submitted via TRFs were summarized at both the chapter level and Clinical Classifications Software Refined Categories (20, 21). Immunocompromising conditions were then flagged using definitions published by the Agency for Healthcare Research and Quality (22–24).

### The Karius Test

Plasma mcfDNA sequencing was performed as previously described (12) in the Karius clinical laboratory, certified under the Clinical Laboratory Improvement Amendments of 1988 and accredited by the College of American Pathologists. Briefly, whole-blood samples were collected in either BD Vacutainer plasma preparation tubes (PPTs) or K2-EDTA tubes. After plasma has been separated from cells, the sample is stable at ambient temperature for 96 hours and at -20°C for 6 months. Upon receipt at Karius, controls for carry-over, sequencing bias, metagenomic sequencing quality, and sample mix-ups were added to the sample. Proprietary chemistries were used to enrich samples for mcfDNA without preselecting pathogens to test. Automated DNA extraction and sequencing library preparation protocols were optimized for high speed and low pathogen bias. Single-end, 76-cycle sequencing was performed on NextSeq 500 instruments (Illumina, San Diego, CA) with an average of >20 million reads/sample. Double-unique dual indexes were used to ensure robust sample demultiplexing. Sequencing data were processed using a proprietary analytical pipeline, and microbial reads were aligned to a database comprising >20,000 curated assemblies from >16,000 species of which >1,500 taxa are reported, including bacteria, DNA viruses, fungi, and parasites (https://kariusdx.com/the-karius-test/pathogen-list/).

Microorganisms present in statistically significant amounts were reported as a concentration of their mcfDNA expressed as MPM, a unique, absolute quantification capability of the Karius Test shown in preliminary work to correlate well with single-analyte quantitative PCR measurements (25, 26). The reports also contained median and range of MPM values observed for each microorganism reported in the last 1,000 specimens, as MPM values from different microbes are not comparable, and a reference interval determined from 675 asymptomatic donors for comparison. We routinely analyzed the raw data for mcfDNA from potential pathogens, including those present at levels below our standard laboratory report thresholds. For this study, we focused on microorganisms identified in statistically significant amounts.

Notable improvements in the test wet bench procedures and analytical pipeline, as may be anticipated, occurred during the study period. We used the operational classes of pathogens described by Relman, Falkow, and Ramakrishnan (27). The operational classes include obligate, commensal, zoonotic, and environmental pathogens.

### Data analytics

Data analysis and visualization were conducted using Python v. 3.9.7, pandas v. 1.3.4, matplotlib v. 3.4.3, and seaborn v. 0.11.2 (28–30). Given the taxonomy and nomenclature for some genera continue to evolve, we selected three (*Legionella*, *Nocardia*, and *Mycobacterium*) to examine species detections, especially multiple species co-detections, more closely.

## RESULTS

### Test cohort

A total of 19,739 samples meeting collection and transport requirements were tested from 16,172 patients in a hospital setting in 39 states and the District of Columbia during the study period. The median time from sample receipt at the Karius laboratory to reported result was 26 hours (IQR 25–28), and 96% of samples had valid test results. A summary of key performance metrics for the test in this production data set are shown in **Table 1**. These metrics were not significantly different (two-sided t-test p-value >0.01) from those reported for the first 2,000 clinical samples run by the Karius clinical laboratory and reported in the initial validation study (12). Infectious disease and hematology/oncology providers represented most ordering clinicians, 64% (n=9,804) and 14% (n=2,132), respectively, for the 15,424 specimens with a National Provider Identifier indicated. We were able to capture and analyze 18,690 reports from 15,165 patients. Twelve percent (n=1,839) of patients had at least one repeat test during the study interval. Almost two thirds (65%, n=9,798) of patients were adults (i.e., age >18 years). More than a quarter (29%, n=4,423) of patients at the time of diagnostic testing had ICD10 codes representing a diverse array of clinical scenarios indicated in their TRFs (**Table 2**). Eighteen percent (n=797) of these patients were indicated as immunocompromised (IC); 717 (16%) had fever; and 230 (5%) had sepsis.

**Table 1.** Plasma mcfDNA sequencing test performance metrics in production, April 2018–end of September 2021.

| Metric | Production<sup>1</sup> | Validation<sup>2</sup> |
| --- | --- | --- |
| Sample acceptance rate | 98% | 98% |
| Total yield | 96% | 98% |
| First pass yield | 91% | 91% |
| Results delivered next operational day | 90% | 90% |

<sup>1</sup>Based on 20,087 clinical samples received during this study.
<sup>2</sup>Based on 2,000 clinical samples tested as part of the initial Karius Test validation study (12)

**Table 2.**
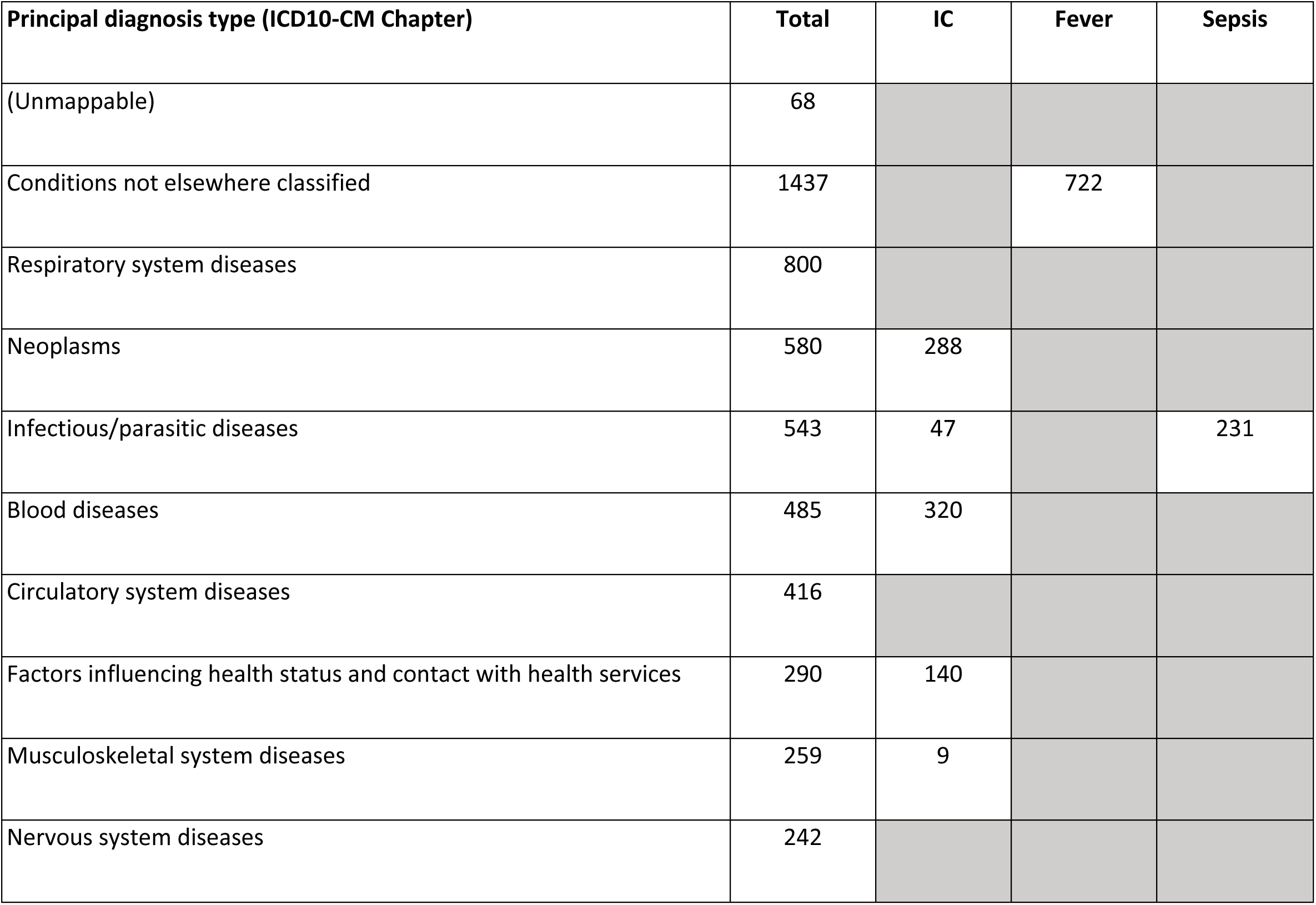
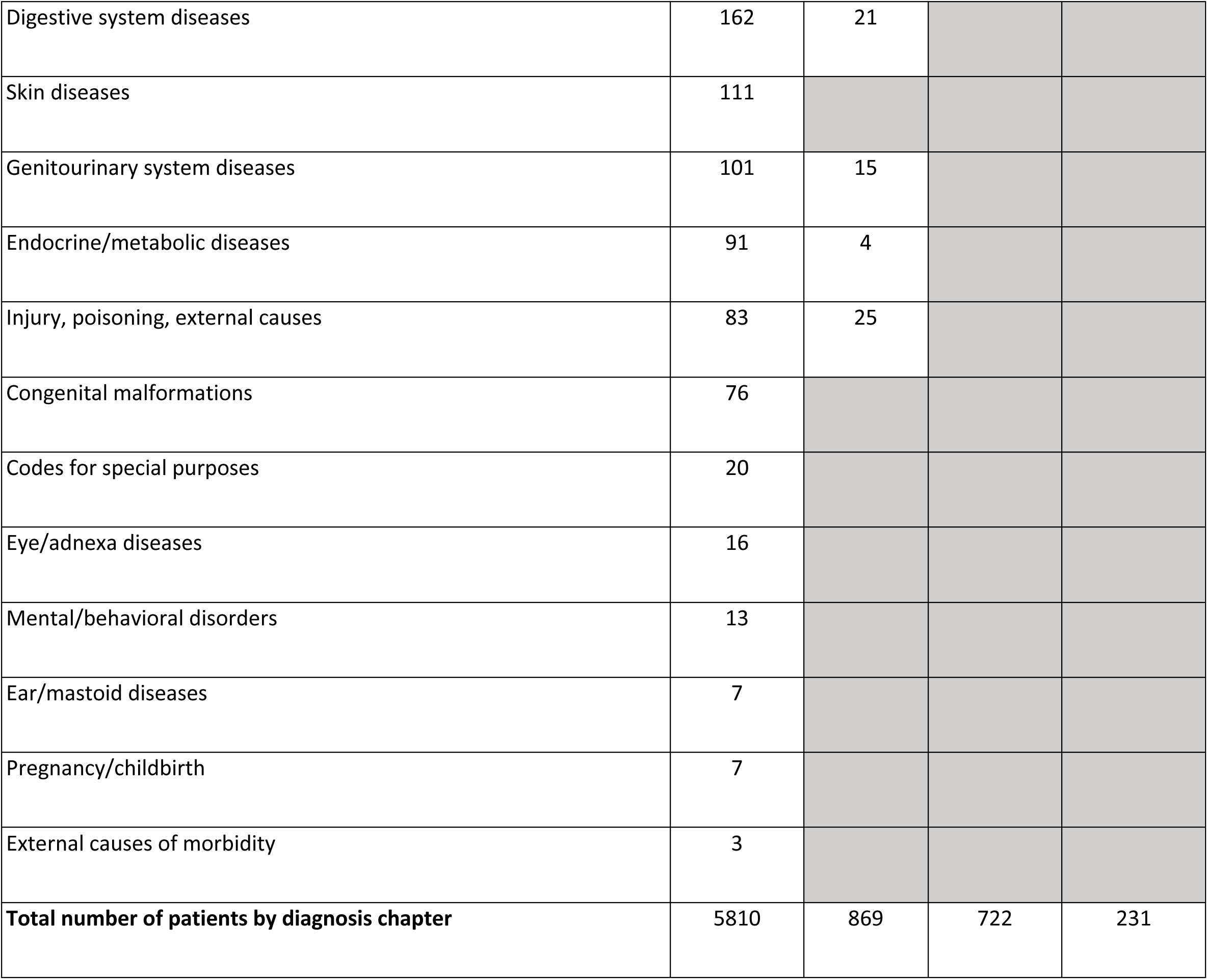
ICD10 codes by principal diagnosis type for those patients with ICD10 codes indicated on Karius TRFs (N=4,423), April 2018–September 2021.

### Taxa detection and quantification

Of samples yielding a valid result, 7,938 (42%) reported a negative test with no pathogens identified. The remaining 10,752 (58%) Karius Test reports from 8,849 patients had at least one microbe identified (5,531 [30%] only one) representing 701 unique microbial taxa (526 [75%] bacteria, 103 [15%] fungi, 47 [7%] viruses, and 24 [3%] parasites) and a total of 22,792 detections. The overall frequency of detection for each of these groups is shown in **Fig. 1**, and the number of detected taxa counts per report for all positive reports is shown in **Fig. 2**. All the quality control metrics were met for taxa quantification in MPM for 9,690 (90%) samples with positive results. A complete list of all taxa reported along with their frequency of detection and median and IQR for the MPM values are given in **Supplemental Table 1**.

**Fig. 1.**
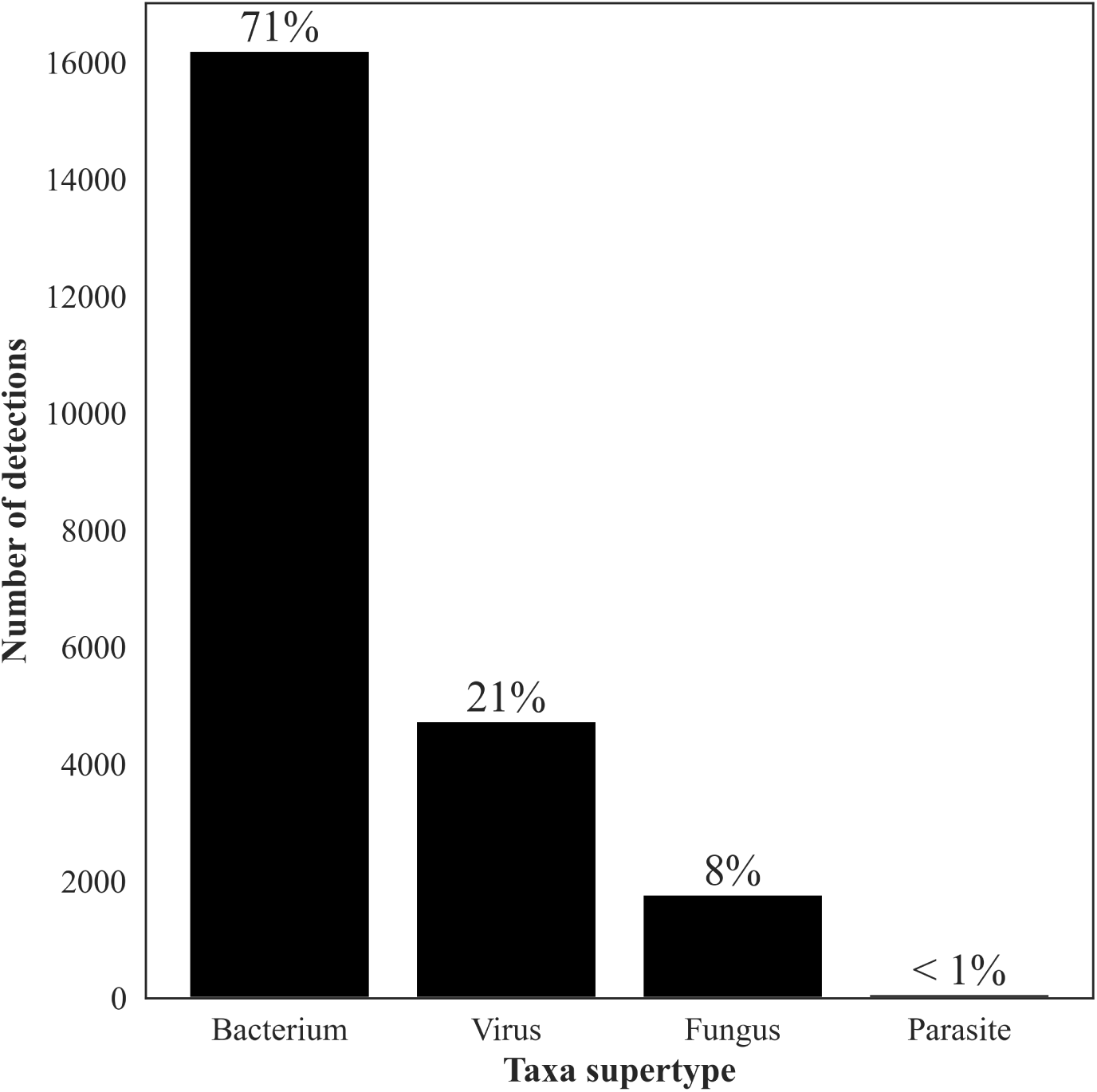
Number of detections by the Karius test of the different super groups of taxa, Apr 2018– Sept 2021, N=22,792: bacteria, 16,221; viruses, 4,737; fungi, 1,758; parasites, 70. Percentages reflect proportion of total number of detections.

**Fig. 2.**
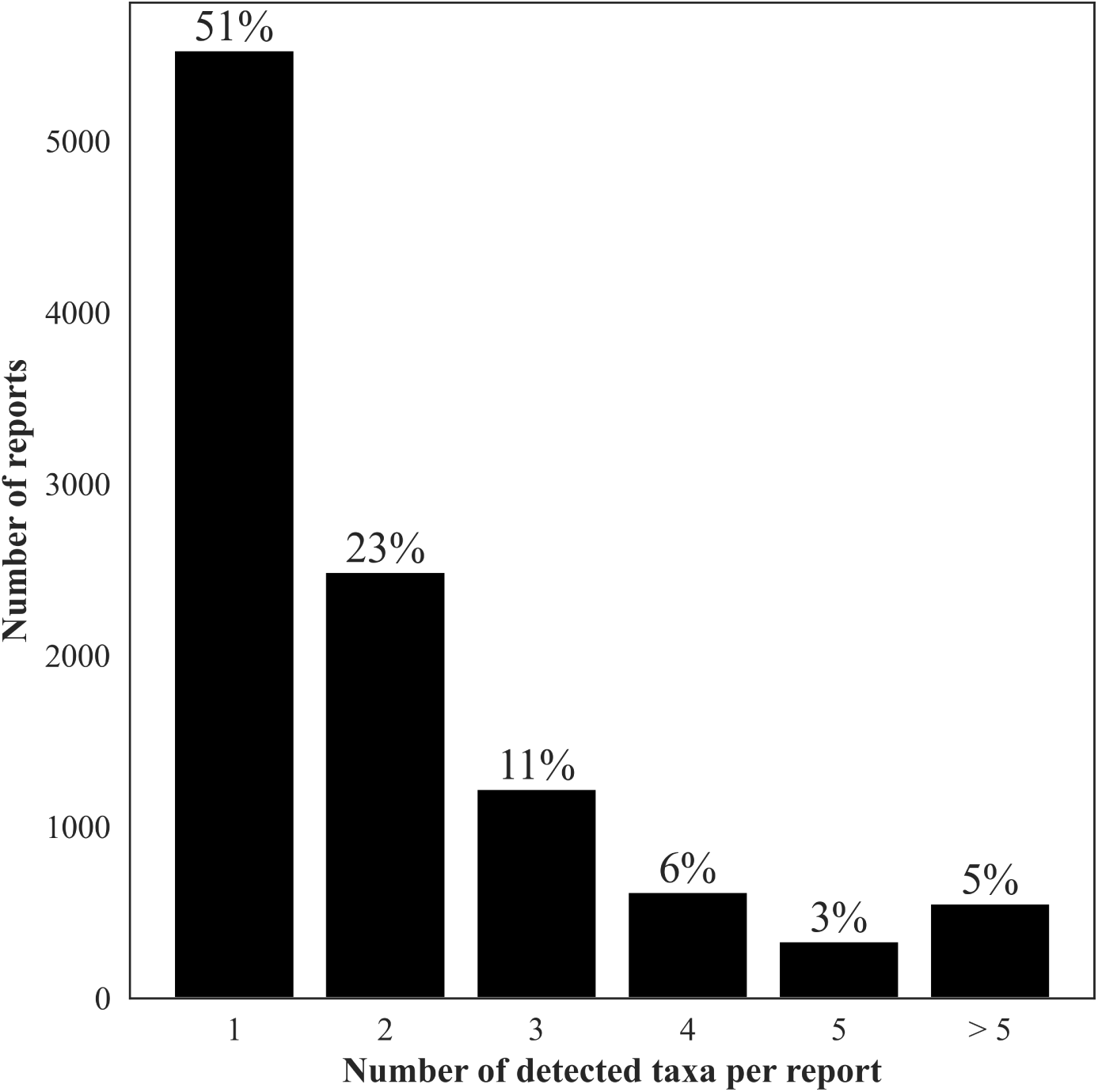
Number of detected taxa counts per report for all positive reports, Apr 2018–Sept 2021. Percentages reflect the proportion of all positive reports (N=10,752).

### Top 50 reported taxa

The top 50 reported taxa and the median, range, and IQR of MPM for each taxon are shown in **Table 3**. They included 36 bacteria, 9 viruses, and 5 fungi. Together the top 50 taxa included a broad range of commensal and environmental pathogens and represented 15,692 detections (69% of all detections).

**Table 3.** Top 50 taxa detected by sorted microorganism group (bacteria, fungi, and viruses) and then alphabetically by taxon name with number detected and median and IQR of molecules/µl (MPM) values for each taxon, April 2018–September 2021. TRF = test report form. IC = immunocompromised. Fever: any ICD10 starting with “R50”; sepsis: any ICD10 starting with “A41”; IC: any ICD10 annotated as immunocompromised from AHRQ code list. Each TRF could contain up to 2 ICD10 codes, and each patient had between 1–5 unique ICD10 codes. For the study period, there were 15,165 patients with a positive or negative report (18,690 reports).

| **Taxon name** | **Number detected** | **Median (MPM)** | **IQR (MPM)** |
| --- | --- | --- | --- |
| **Bacteria** |  |  |  |
| *Acinetobacter haemolyticus* | 199 | 142.97 | 192.68 |
| *Bacteroides fragilis* | 217 | 264.74 | 1,046.15 |
| *Bacteroides ovatus* | 133 | 141.43 | 476.14 |
| *Bacteroides thetaiotaomicron* | 146 | 221.79 | 742.60 |
| *Bacteroides vulgatus* | 392 | 153.01 | 413.32 |
| *Enterobacter cloacae complex* | 343 | 821.34 | 6,742.30 |
| *Enterococcus faecalis* | 582 | 617.90 | 3,285.15 |
| *Enterococcus faecium* | 510 | 687.63 | 3,958.33 |
| *Escherichia coli* | 1088 | 684.92 | 3,623.17 |
| *Fusobacterium nucleatum* | 299 | 460.25 | 1,922.08 |
| *Haemophilus influenzae* | 288 | 157.09 | 425.59 |
| *Haemophilus parainfluenzae* | 178 | 284.92 | 1,121.19 |
| *Helicobacter pylori* | 207 | 217.44 | 601.83 |
| *Klebsiella pneumoniae* | 501 | 910.19 | 5,889.84 |
| *Lactobacillus fermentum* | 165 | 256.13 | 1,058.08 |
| *Lactobacillus rhamnosus* | 281 | 344.33 | 1,250.14 |
| *Prevotella melaninogenica* | 509 | 208.52 | 545.66 |
| *Prevotella oris* | 170 | 517.78 | 2,094.07 |
| *Pseudomonas aeruginosa* | 817 | 1,005.18 | 6,874.18 |
| *Rothia dentocariosa* | 110 | 117.02 | 256.72 |
| *Rothia mucilaginosa* | 454 | 177.53 | 369.36 |
| *Serratia marcescens* | 99 | 492.72 | 2,956.54 |
| *Staphylococcus aureus* | 561 | 838.21 | 5,665.43 |
| *Staphylococcus epidermidis* | 716 | 579.58 | 2,689.68 |
| *Staphylococcus haemolyticus* | 92 | 1,137.08 | 5,450.01 |
| *Stenotrophomonas maltophilia* | 169 | 1,983.18 | 11,864.14 |
| *Streptococcus intermedius* | 100 | 354.95 | 1,048.47 |
| *Streptococcus mitis* | 356 | 226.15 | 897.48 |
| *Streptococcus oralis* | 140 | 235.54 | 730.42 |
| *Streptococcus parasanguinis* | 186 | 152.35 | 298.03 |
| *Streptococcus pneumoniae* | 214 | 975.48 | 11,933.10 |
| *Streptococcus pyogenes* | 95 | 1,064.08 | 9,773.54 |
| *Streptococcus salivarius* | 104 | 253.73 | 799.10 |
| *Streptococcus thermophilus* | 184 | 190.84 | 339.33 |
| *Veillonella dispar* | 162 | 140.88 | 269.85 |
| *Veillonella parvula* | 256 | 202.30 | 476.57 |
| **Fungi** |  |  |  |
| *Aspergillus fumigatus* | 186 | 300.10 | 723.60 |
| *Candida albicans* | 260 | 624.63 | 2,074.97 |
| *Candida glabrata* | 113 | 611.85 | 1,449.20 |
| *Candida tropicalis* | 103 | 784.49 | 3,997.85 |
| *Pneumocystis jiroveci* | 258 | 2,452.78 | 18,084.89 |
| **Viruses** |  |  |  |
| *BK polyomavirus* | 354 | 472.38 | 2,677.18 |
| Cytomegalovirus | 1275 | 633.93 | 3,889.67 |
| Epstein-Barr virus | 902 | 272.34 | 852.32 |
| Herpes simplex virus type 1 | 479 | 754.78 | 3,208.06 |
| Human herpesvirus 6B | 468 | 518.74 | 3,268.72 |
| Human herpesvirus 7 | 98 | 82.74 | 342.37 |
| Human adenovirus B | 100 | 3,184.75 | 69,714.71 |
| Human adenovirus C | 171 | 573.87 | 10,160.48 |
| Torque teno virus | 135 | 484.19 | 1,935.97 |

The distribution of these taxa are as follows. There were 11,023 detections of bacteria including 11 anaerobes (2,730, 25%), 8 *Streptococcus* spp. (1,379, 12%), 4 *Enterobacterales* (2,031, 18%), *3 Staphylococcus* spp. (1,369, 12%)., 2 *Rothia* spp. (564, 5%), 2 *Haemophilus* spp. (466, 4%), 2 *Enterococcu*s spp. (1,092, 10%), and 1 each of *Acinetobacter haemolyticus* (199, 2%), *Pseudomonas aeruginosa* (817, 7%), *Stenotrophomonas maltophilia* (169, 1%), and *Helicobacter pylori* (207, 2%). There were 3,982 viral detections of 9 different viruses that included 1,275 (32%) cytomegalovirus, 902 (23%) Epstein-Barr virus, 479 (12%) herpes simplex virus 1, 468 (12%) human herpes virus 6B, 354 (9%) BK polyoma virus, 171 (4%) human adenovirus C, 135 (3%) torque teno virus (TTV), 100 (3%) human adenovirus B, and 98 (3%) human herpes virus 7. Finally, there were 920 detections of fungi comprising 260 (28%) *Candida albicans*, 258 (28%) *Pneumocystis jirovecii*, 186 (20%) *Aspergillus fumigatus*, 113 (12%) *Candida glabrata*, and 103 (11%) *Candida tropicalis*.

### Difficult to diagnose uncommon pathogens

The SOC methods for the organisms listed below have considerable shortcomings including, but not limited to, sensitivity and specificity, comprehensiveness, accuracy, time to result, and/or local availability.

#### Bacteria

The frequency distribution of the number of detections of *Legionella*-like organisms (n=80) is shown in **Fig. 3**. Forty-one percent of these detections were the most recognized pathogen, *L. pneumophila*. Two reports contained co-detections of two different species (*L. brunensis*, 400 MPM and *hackeliae*, 270 MPM; and *L. feeleii*, 78,508 MPM and *L. tunisiensis*, 76,445 MPM, respectively). Neither *L. brunensis* nor *L. tunisiensis* has been associated with human disease (https://specialpathogenslab.com/legionella-species/), and the MPM values for the co-detections in each report were similar.

**Fig. 3.**
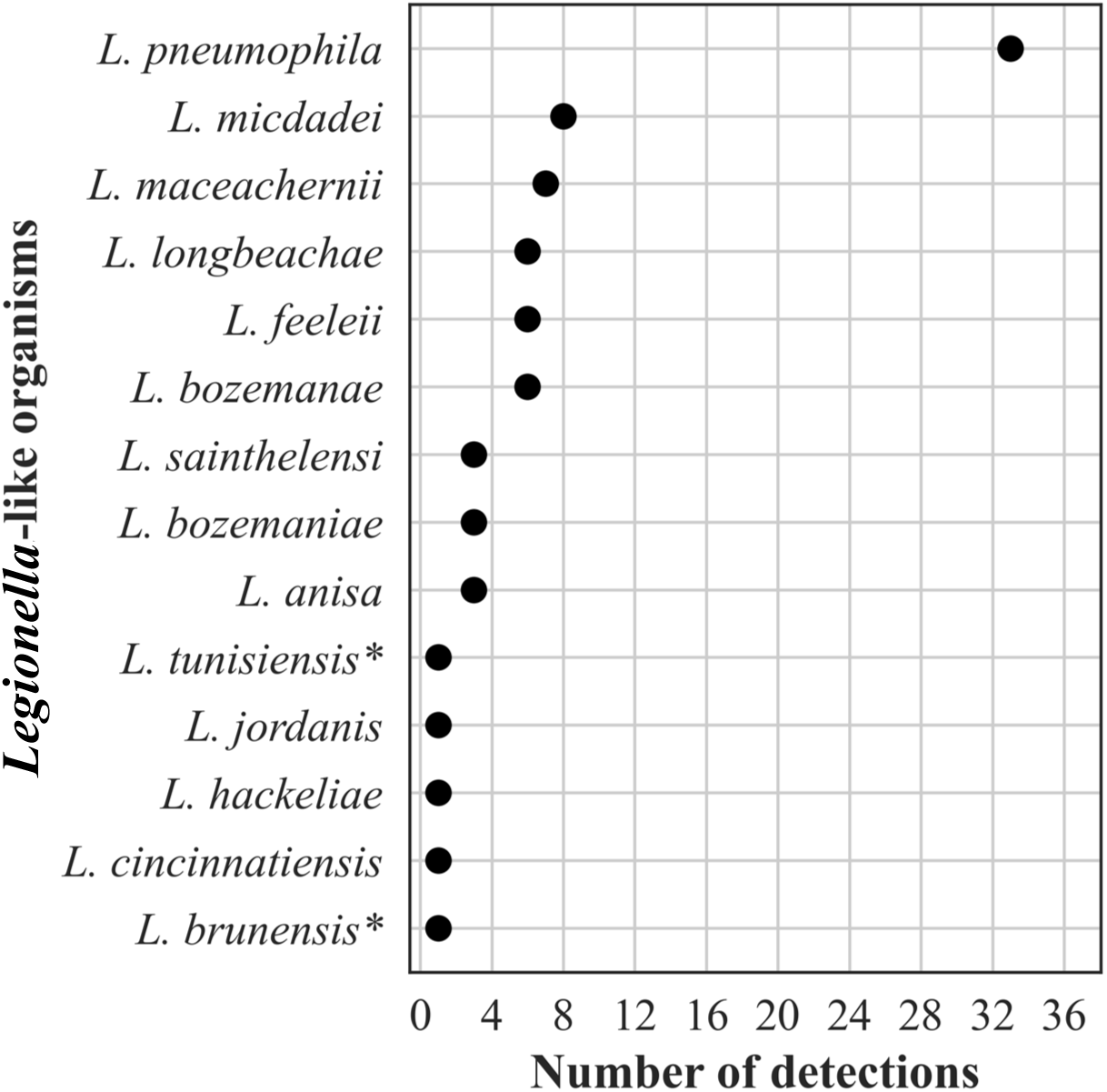
Frequency distribution of different *Legionella*-like organisms detected, n=80 (< 1% of all bacterial detections, N=16,203). *Indicates species not previously detected in humans.

The frequency distribution of *Nocardia* spp. detections, n=76, is shown in **Fig. 4**. Plasma mcfDNA sequencing detected 25 of the approximately 100 validly named species. Of the 8 species reported to be isolated more frequently from patients, 7 were detected (**Fig. 4**). One species, *N. cyriacigeorgica,* dominated with 19 (25%) detections. Of the 69 patient reports represented by the *Nocardia* spp. detections, 12 (17%) reported ≥2 concurrent species (range 2–5). All the co-detections were reported with similar MPM values, and 8 (67%) were co-detections of closely related species (4 *N. exalbida*/*gam*kensis, 3 *N. elegans*/*nova*/*africana*, 1 *N. kruczakiae/violaceofusca/aobensis*) previously reported to be indistinguishable by mcfDNA sequencing (31, 32).

**Fig. 4.**
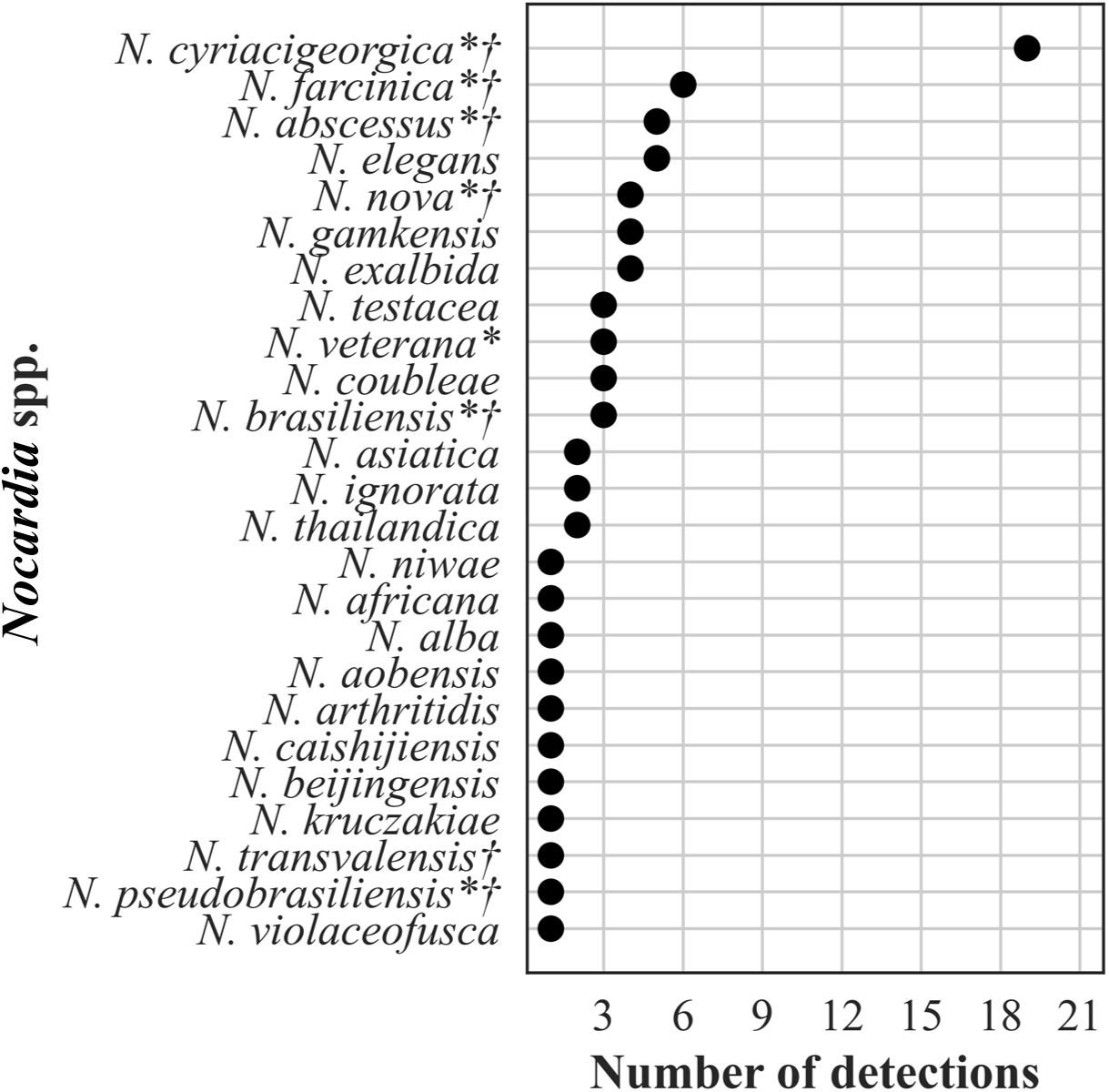
Frequency distribution of Nocardia spp. detections, n=76 (< 1% bacterial detections, N=16,203). *Indicates species more frequently isolated from clinical specimens (69). †Indicates species-specific susceptibility patterns (54).

The frequency distribution of the 156 *Mycobacterium* spp. detections is shown in **Fig 5**. Plasma mcfDNA sequencing detected 107 (69%) slowly growing mycobacteria (SGM) and 49 (31%) rapidly growing mycobacteria (RGM). The frequency of species distributions of these three genera showed similarities in that several species were predominant, followed by long tails of uncommon to single species detections. We reported ≥2 concurrent species (range 2–6) in 6 (4%) of the 144 reports including *Mycobacterium* spp. All the co-detections were reported with similar MPM values. Three of the reports contained *M. avium complex*/*chimera* and one each *M. avium* complex/*celatum*/*kyorinense*, *M. brisbanense*/*mucogenicum*/*obuense*, and *M. chubuense*/*elephantis*/*flavescens*/*goodii*/*holsaticum* /*phlei*. The co-detections of multiple *Legionella*, *Nocardia*, and *Mycobacterium* spp., respectively, within the same patient are shown in **Supplemental Table 2** .

**Fig. 5.**
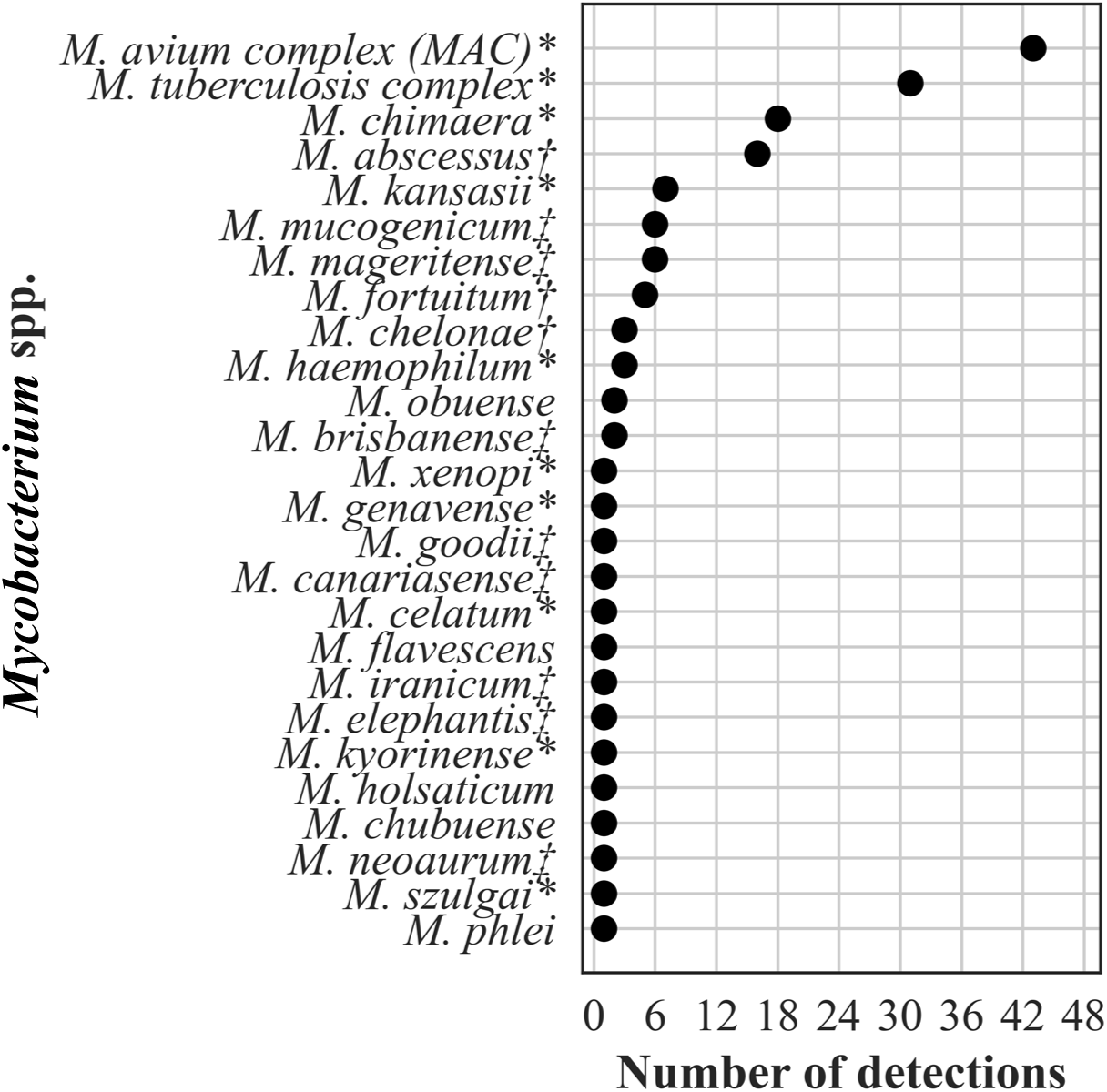
Frequency distribution of *Mycobacterium* spp. detections, n=156 (1% of all bacterial detections, N=16,203). *Indicates slowly growing mycobacteria of established clinical significance (70). †Indicates rapidly growing mycobacteria considered common human pathogens. ‡Indicates rapidly growing mycobacteria considered less common or rare human pathogens (71).

The frequency distribution of the 247 (3% of all bacterial detections) zoonotic and vector borne bacterial detections are shown in **Fig. 6**. *Bartonella henselae* predominated with 90 (36%) detections.

**Fig. 6.**
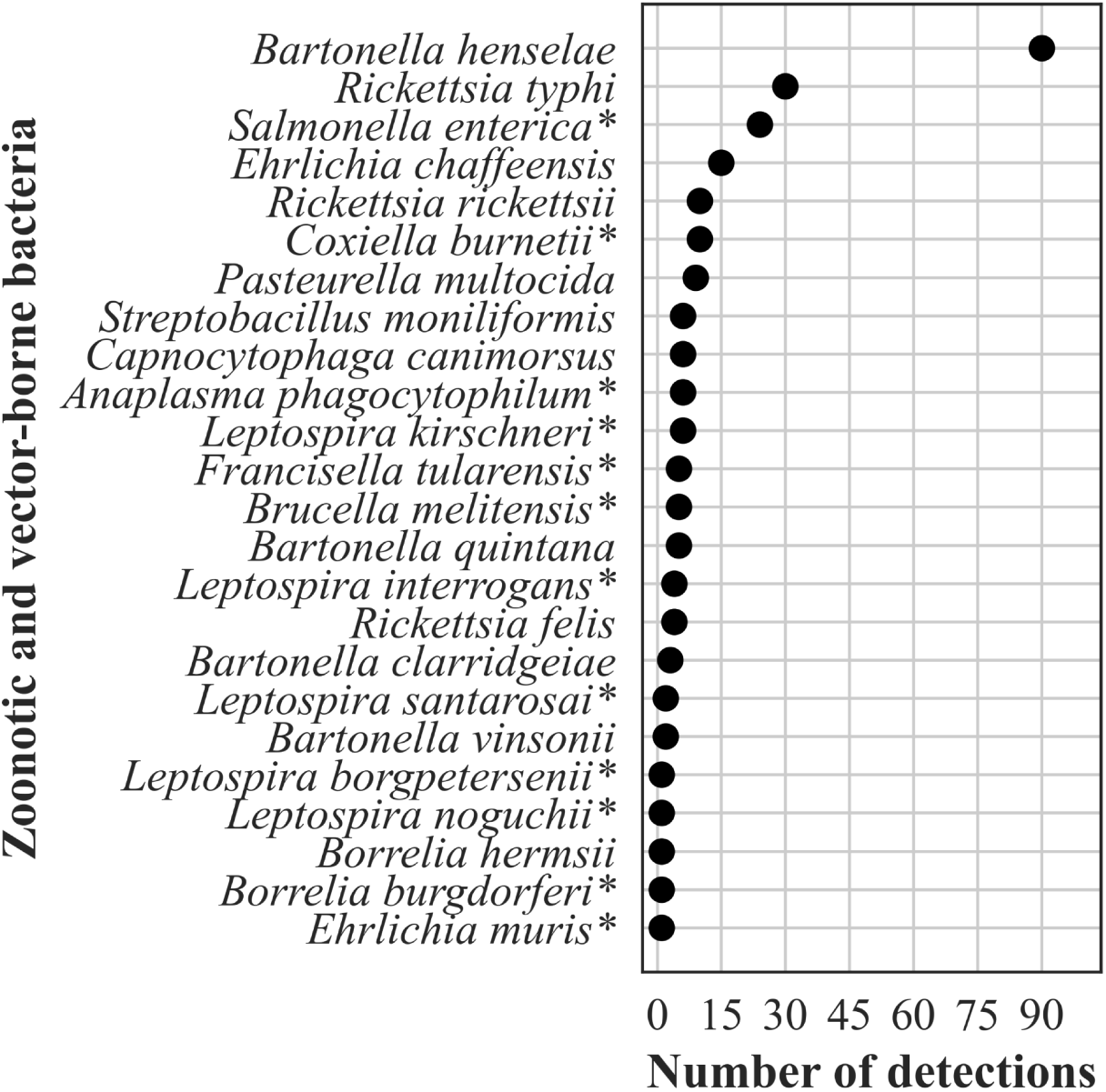
Frequency distribution of zoonotic and vector borne bacteria detections, n=247 (2%of all bacterial detections, N=16,203). *Indicates bacteria causing a nationally notifiable infectious disease (72).

#### Fungi

The frequency distribution of the 632 *Candida* spp. detections is shown in **Fig 7**; 374 *Aspergillus* spp. detections in **Fig 8**; 196 detections in the order *Mucorales* in **Fig. 9**; 78 detections of the systemic dimorphic fungi in **Fig. 10;** and 33 detections of dematiaceous fungi in **Fig. 11**. We detected 9 microsporidia, including 5 *Enterocytozoon bieneusi* and one each of *E. cuniculi*, *E. hellem*, *Anncaliia algerae*, and *Vittaforma corneae*. In addition, *Pneumocystis jirovecii* (258 detections) was among the top 50 taxa detected.

**Fig. 7.**
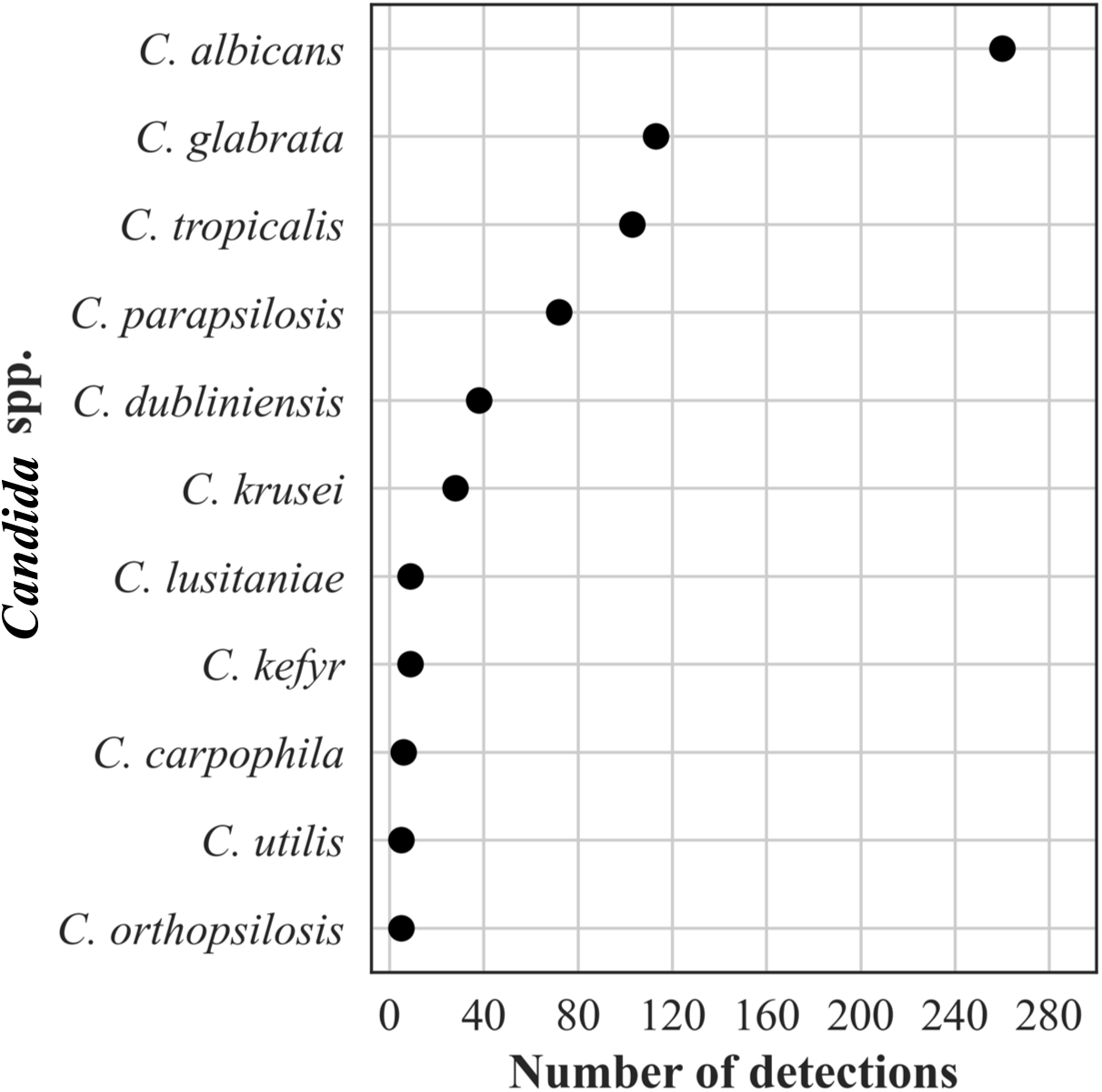
Frequency distribution of *Candida* spp. detections, n=648 (36% of all fungal detections, N=1,776).

**Fig. 8.**
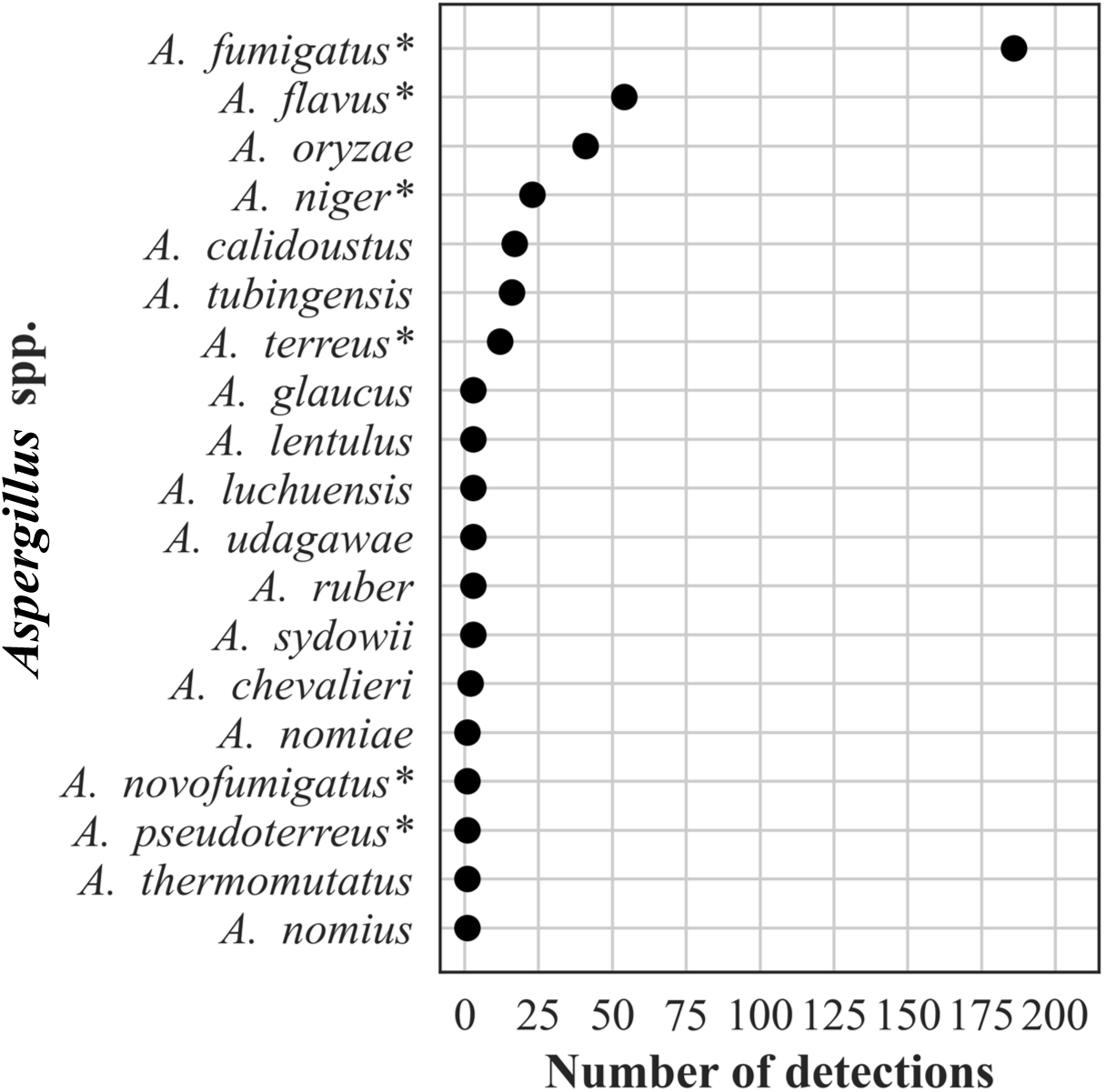
Frequency distribution of Aspergillus spp. detections, n=374 (21% of all fungal detections, N=1,776). *Indicates most common pathogenic species.

**Fig 9.**
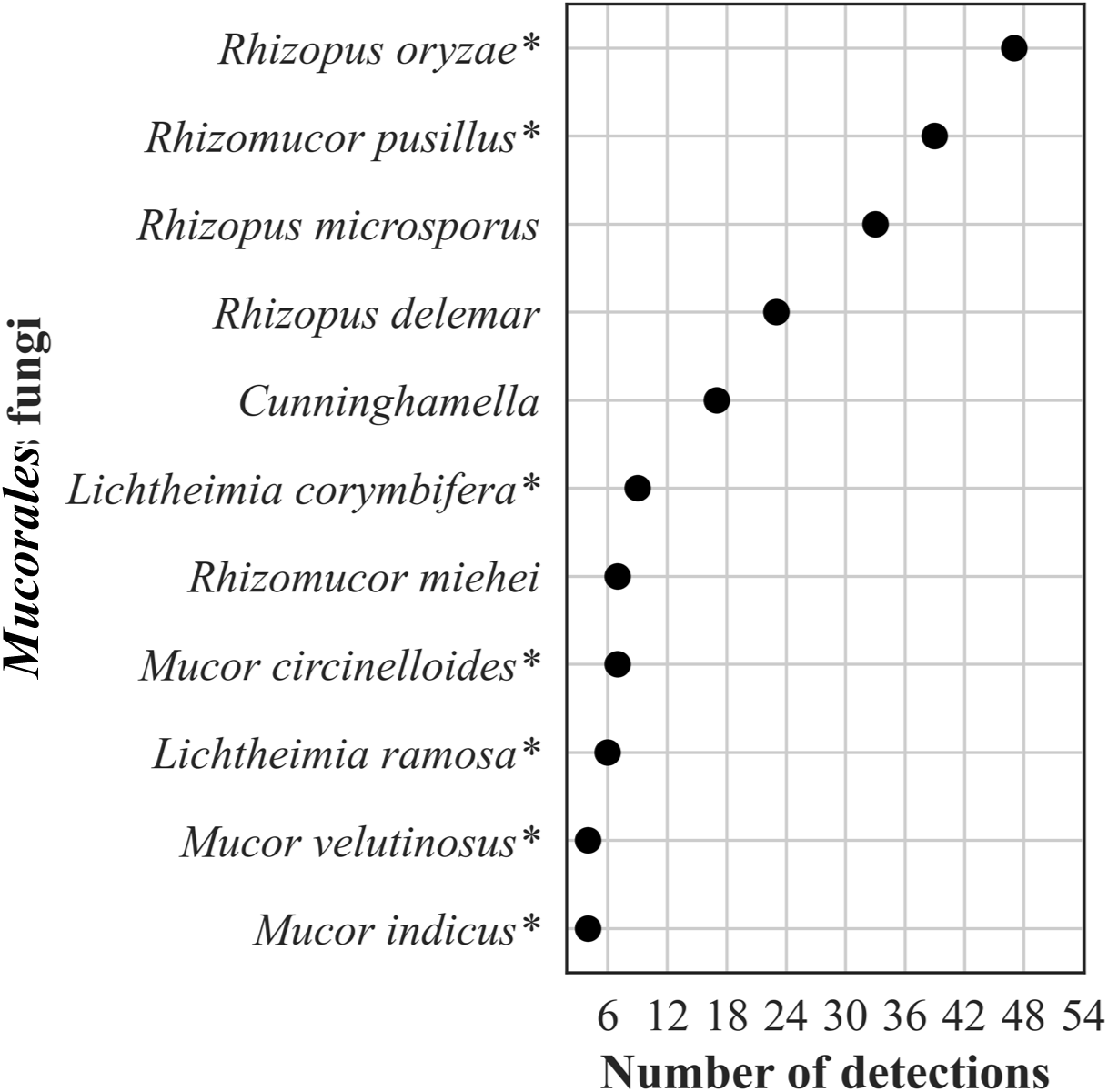
Frequency distribution of detections in the order Mucorales, n=196 (11% of all fungal detections, N=1,776). *Indicates taxa implicated in human mucormycosis (73).

**Fig 10.**
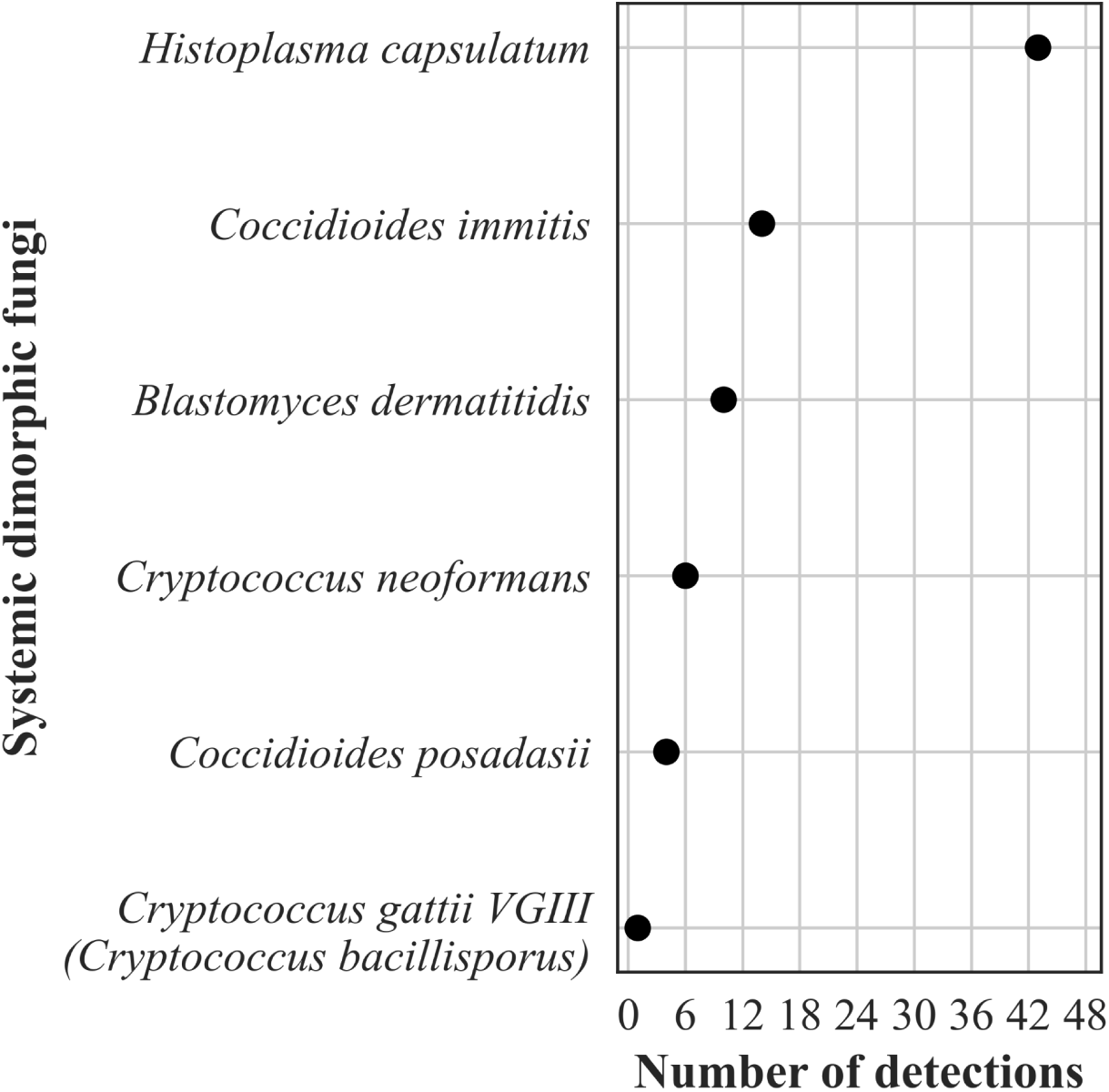
Frequency distribution of detections of systemic dimorphic fungi, n=78 (4% of all fungal detections, N=1,776).

**Fig 11.**
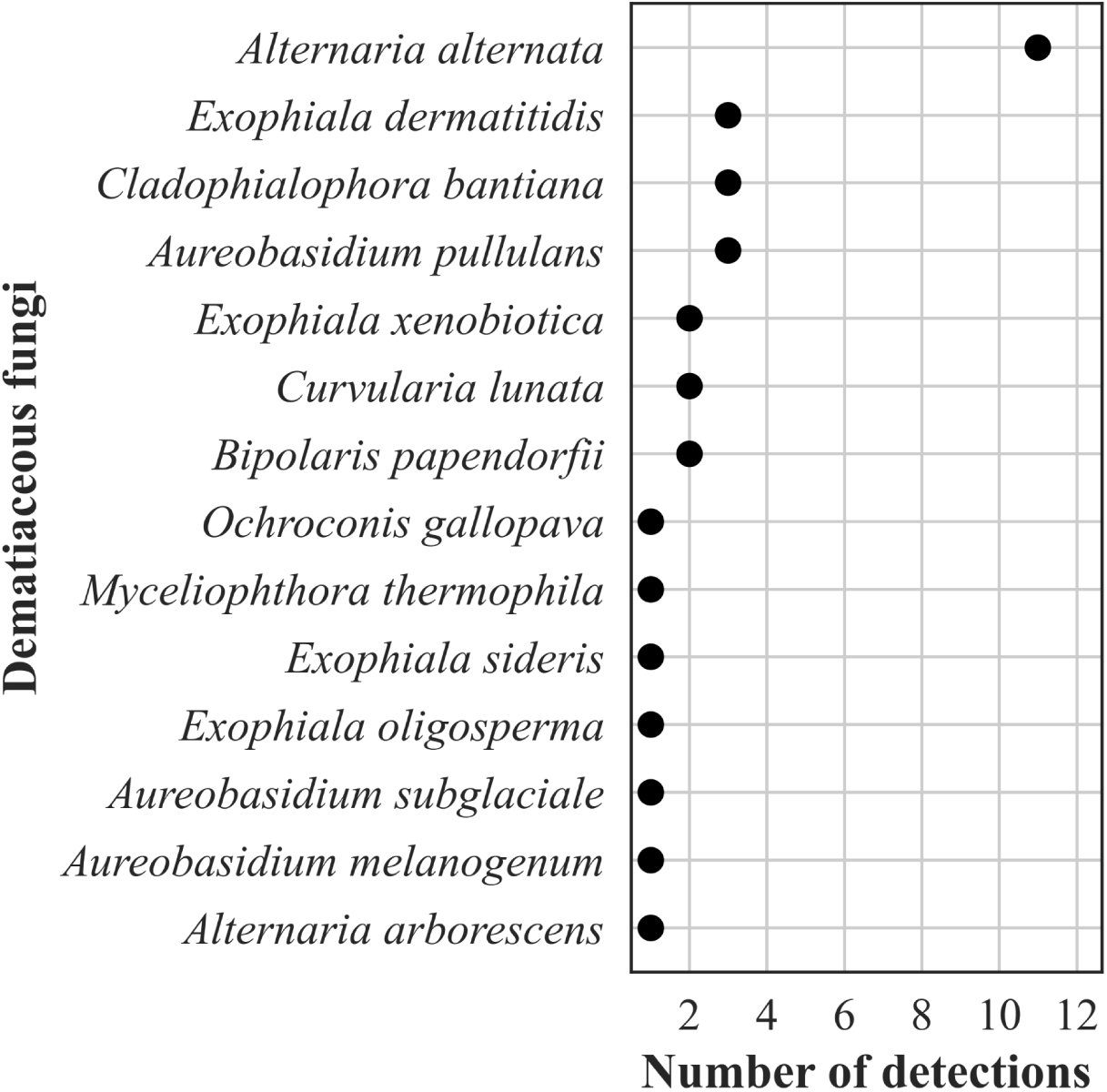
Frequency distribution of detections of dematiaceous fungi, n=33 (2% of all fungal detections, N=1,776).

#### Eukaryotic parasites

The frequency distribution of the 57 (89% of 64 parasite detections) protozoa is shown in **Fig. 12**. Among the protozoan parasite detections, 68% were *Toxoplasma gondii*, and 14% were pathogenic amoebae. Among the 7 (11%) helminthic parasites, we detected 4 nematodes (all *Strongyloides stercoralis*), 2 cestodes (both *Echinococcus multilocularis*), and 1 trematode (*Schistosoma mansoni*).

**Fig 12.**
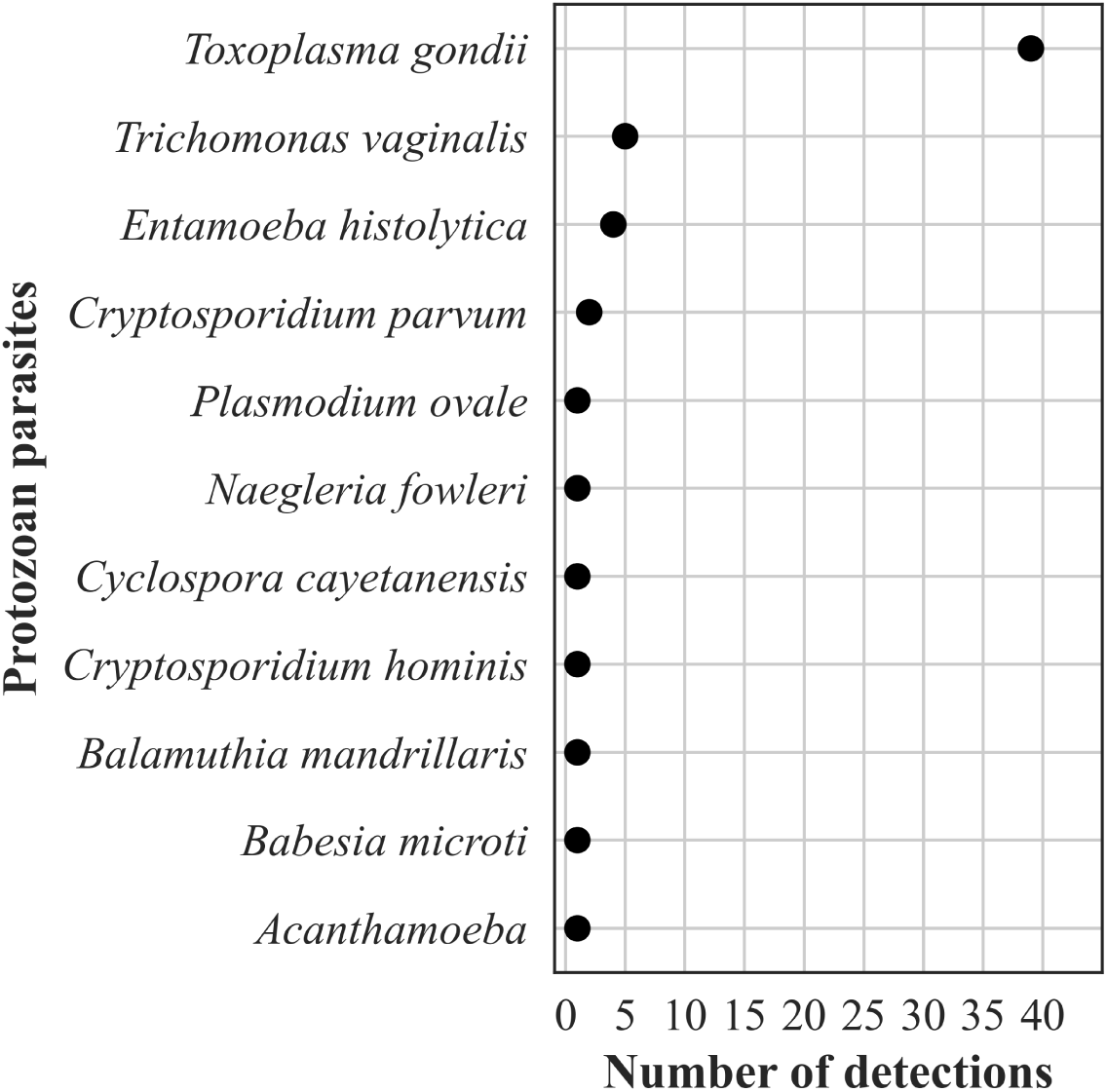
Frequency distribution of detections of protozoan parasites, n=57 (89% of all eukaryotic parasite detections, N=64).

## DISCUSSION

We report the largest testing cohort of patients in which mcfDNA was identified and quantified. The key test performance metrics in this large cohort mirror what was reported with a much smaller cohort in the initial validation study (10), demonstrating that the Karius Test is robust and can be performed at scale in a clinically relevant time frame. These mcfDNA sequencing data reaffirm the ubiquity of some infections, as commonly expected microbes were detected in most patients while less common microbes were rarely detected. However, notably, the unbiased approach of the plasma mcfDNA sequencing made those “rare” detections possible, whereas conventional diagnostics require targeting specific organisms. Identifying optimal utilization, both clinical indications and timing, of the plasma mcfDNA sequencing in future studies to augment clinical decision-making and integration into current testing algorithms (17, 18, 33–35) will serve to improve the utility of the test. Examples of such studies include two recently completed prospective, observational clinical trials of the use of the Karius Test to diagnose pneumonia in immunocompromised patients (NCT04047719) (36) and infections in stem cell transplant inpatients and outpatients over time (NCT02804464) (26), respectively.

Our finding that 58% of the Karius Test reports identified ≥1 pathogen is substantially lower than the 70–85% reported for the test when applied to well-defined clinical uses (13, 14, 37). These findings support the need for diagnostic stewardship and additional clinical studies demonstrating how yield differs and should be carefully interpreted according to the specific patient population and disease prevalence considered.

The Karius Test quantitates detections in MPM, which can be influenced by several factors specific to the microbial genome—e.g., turnover rate and genome size (12). Confounding patient variables (e.g., infection site, therapeutic interventions, and immune status) may also influence this measure. Generally higher concentrations have been found in definite infections, but since considerable overlap of MPM values exists in unlikely infections and in the asymptomatic cohort, threshold values for definite infections have not been established (12). However, following the decay of mcfDNA quantitatively by serial testing may have important implications for individual patient management in assessing the effectiveness of antimicrobial therapy and other medical or surgical interventions (8, 38, 39), but needs further study.

Notable among the most common detections were the many commensal bacterial and fungal pathogens that cause serious and often invasive infections in patients with relevant risk factors (e.g., immunocompromised). Among the viruses most detected by the Karius Test (e.g., *Herpesviridae*), all could represent latent infection, reactivation, or active infections depending on the respective clinical conditions; regardless, the detection of these viruses may be of particular concern for immunocompromised patients for whom they may cause considerable morbidity (40).

A key benefit of unbiased mcfDNA sequencing identified in this study is the ability to detect diagnostically challenging microbes such as opportunistic and systemic dimorphic fungi and zoonotic and vector borne pathogens (41). These pathogens often carry a sense of urgency for the management of the individual patient and even for the public’s health, as they may be associated with considerable morbidity and potential mortality. As some are uncommonly expected pathogens, they present a major challenge for clinicians in considering and ordering appropriate SOC testing to capture all possible pathogens. For the laboratories, the microbiologic diagnosis of these infections often represents a major challenge for SOC methods as has been described by others (42, 43).

The absence of certain pathogens such as the emerging, multidrug resistant *C. auris* among the detections may arguably reflect the relatively few cases occurring during the study period in the geographic locations or patient populations of the test cohort and reflects the rarity of clinical cases in the United States (44). Plasma mcfDNA sequencing has the potential to contribute to the greater understanding of the geographic distribution of microbes and even overall disease surveillance, as demonstrated by the systemic dimorphic fungi detections. At 43 detections (55% of dimorphic fungal identifications), *Histoplasma capsulatum* predominated, very likely related to its wide geographic range and opportunities for environmental exposure and mirrors what is known about the epidemiology and incidence of systemic dimorphic fungal infections (45). The relative frequency of detections of *Coccidioides immitis* and *posadasii* and *Blastomyces dermatidis* may have been influenced by the geographic bias in this study sample cohort. Of the nine *Cryptococcus* spp. detected, *gattii* was detected only once compared with *neoformans*, reflecting its restricted geographic distribution and the overall rarity of infections in the United States (46).

Among the rarely detected microbes during the study period are some deserving further mention. The detection of *Legionella*, an obligate pathogen, signals public health concern, whether community or hospital acquired. *L. pneumophila* serogroup 1 (LP1) is estimated to cause 84% of community acquired Legionnaire’s disease (LD) (47); however, other serogroups of *L. pneumophila* and other *Legionella* spp. may cause 60% of hospital acquired LD (48). Urinary antigen detection is the most common diagnostic in the United States and Europe; yet, given its specificity for LP1, as many as 40–50% of patients with non-LP1 legionellosis could be missed (49, 50). Plasma mcfDNA sequencing provides comprehensive testing for *Legionella* and *Legionella*-like organisms in one diagnostic test and could thereby expand the known LD epidemiology, particularly in nosocomial cases.

*Nocardia* have 8 species-specific drug susceptibility patterns (51). While accurate species identification can predict antimicrobial susceptibility patterns, molecular methods are required for accurate identifications but are not widely available. The Karius Test detected in our cohort 7 of the 8 species with recognized susceptibility patterns and can provide results more rapidly than existing approaches to species identification.

Finally, culture for mycobacteria, while a complicated and lengthy process, is considered the gold standard, supplemented by direct detection of *M. tuberculosis* complex by nucleic acid amplification tests (NAATs) in many laboratories. However, NAATs for the direct detection of nontuberculous mycobacteria are not widely available, and accurate identification to species level from cultured isolates remains challenging for most laboratories. The *M. chimaera* may be overrepresented in our cohort since the Karius Test was optimized for its detection following reports of infections occurring post-surgeries employing contaminated cardiopulmonary bypass devices (52). However, these detections demonstrate the capability of mcfDNA sequencing to provide comprehensive identification of these important obligate and commensal pathogens directly from plasma and provides increased diagnostic value to the above-mentioned SOC methods (53).

Plasma mcfDNA sequencing offers a non-invasive means of detecting microbial infection and capturing species diversity, potentially revealing new insights on genetic complexity not resolved by current taxonomic classification. Still, our findings highlight the need to expand currently available genomic sequencing libraries as many species across various genera remain undiscovered or undescribed (54, 55). The species co-detections demonstrated among the genera we highlighted, *Legionella*, *Nocardia*, and *Mycobacteria*, specifically those occurring at similar MPM values, suggest detection of a single divergent strain, a species not in the Karius Test database, or, alternatively, true polymicrobial detections. Similar challenges exist for broad range PCR testing (5, 56) and occurred with adoption of proteomic identification by MALDI-TOF mass spectrometry (57).

This study has several limitations preventing us from directly comparing and fully elucidating plasma mcfDNA sequencing impact on patient care. Orthogonal testing data (including type, timing, and results) were not available for comparison with the Karius Test results; nor was information regarding antimicrobial or other therapies or procedures. Also, the clinical context for using the Karius Test was provided in only 29% of patients; that information was voluntarily provided, may have been incomplete, and would reflect the clinician’s perspective at the time they ordered the test vs. the final diagnosis.

However, others have reported potential increased diagnostic yield of plasma mcfDNA sequencing compared with SOC tests in certain clinical scenarios (14–16, 58). Some have noted the test to be commonly applied in managing severely ill, especially IC, patients (59), who are more likely to be infected with unusual, and difficult to diagnose pathogens (60, 61).

Constraints of the underlying data structure prevented analyzing the data from those patients who had repeated testing to determine whether these tests were performed serially to diagnose new suspected infections or to monitor therapy response or conduct pathogen surveillance among immunocompromised patients as suggested by others (8, 26, 62–64). Further, the underlying data structure limited our ability to stratify the results by notable improvements to the bioinformatics pipeline over the course of the study period. Nevertheless, the unique data from this large testing cohort provides a wealth of information to help improve our understanding of infecting pathogens.

Unbiased plasma mcfDNA sequencing can potentially enhance patient outcomes by direct and timely recognition of pathogens in specific clinical scenarios as well as benefit the overall public health by increasing our understanding of the epidemiology of emerging infectious diseases such as monkeypox virus or *Borrelia miyamotoi* (M.S. Lindner, K. Brick, N. Noll, S.Y. Park, *et. al.*, submitted for publication; L.A. Rubio, A.M. Kjemtrup, G.E. Marx, S. Cronan, *et. al.*, submitted for publication, respectively). Further, this powerful, novel diagnostic tool may facilitate medical advances through recognizing previously unanticipated pathogens, as noted when mcfDNA sequencing was leveraged in a research use only modality to identify porcine cytomegalovirus infection in a patient who had received a genetically modified porcine-to-human cardiac transplant (65). As with any advanced diagnostic tool, careful, timely clinical application with expert guidance and interpretation of its results as well as appropriate diagnostic stewardship will optimize its application to offer even greater clinical impact. In addition, the development of robust clinical outcomes studies to evaluate the clinical impact and cost effectiveness of plasma mcfDNA sequencing for specific clinical indications to guide use remains a top diagnostic stewardship priority.

## Data Availability

All data produced in the present work are contained in the manuscript.

## ACKNOWLEDGEMENTS

We thank the talented, experienced, and dedicated clinical laboratory operations and analytics teams at Karius for generating the test results and sequencing analyses, respectively, that were foundational to this study. We also thank T. Matthew Hill, PharmD, PhD, formerly with Karius, Inc., and Asim A. Ahmed, MD for their early contributions to the paper. This study was supported by Karius. Nathan Ledeboer, PhD and Kevin Messacar, MD, PhD both serve as uncompensated consultants for Karius on this manuscript. Their relationship with Karius has been reviewed and approved by the Medical College of Wisconsin and the University of Colorado, Children’s Hospital of Colorado, respectively, in accordance with their conflict of interest policies.

---

**Supplemental Table 1.**
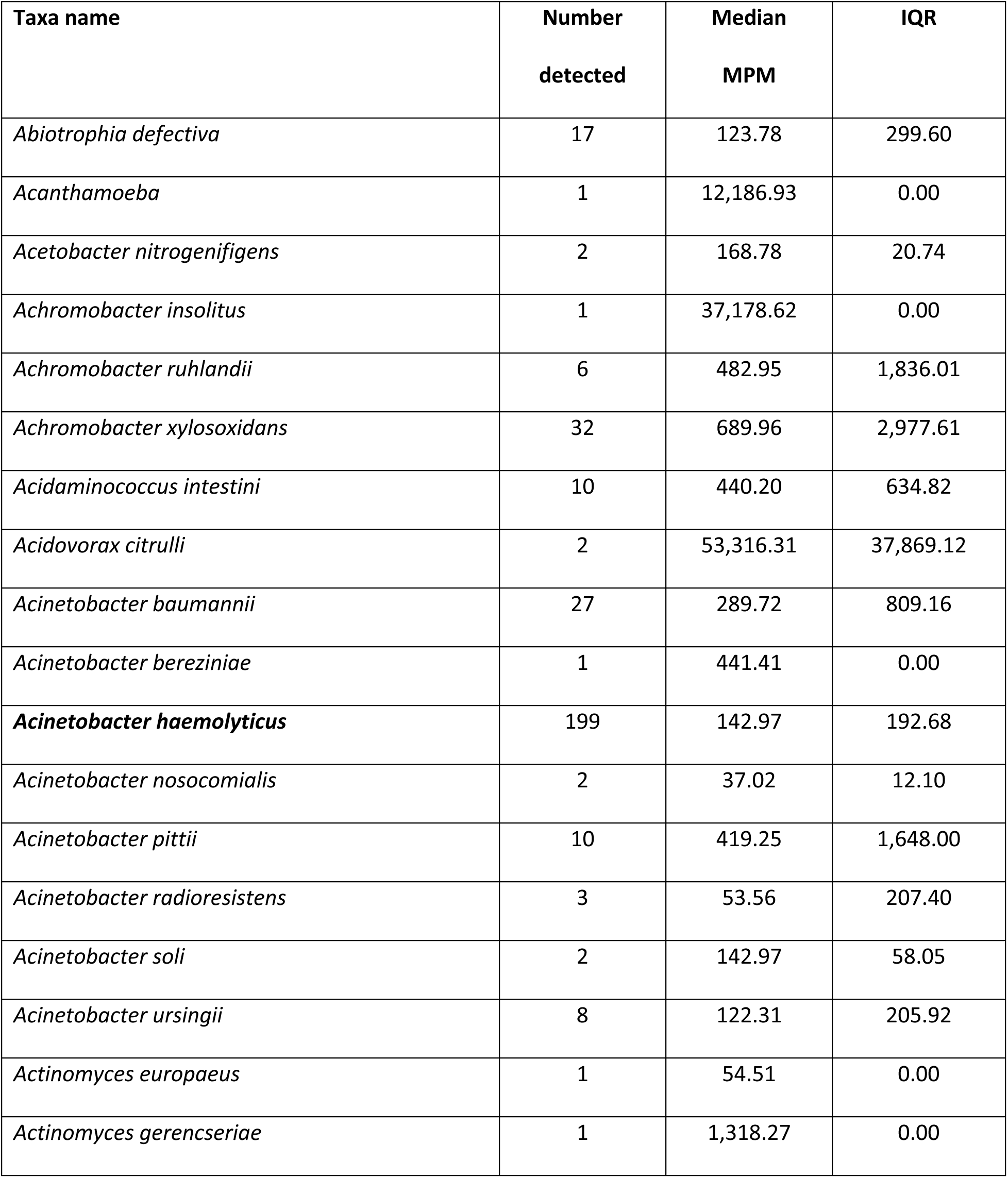
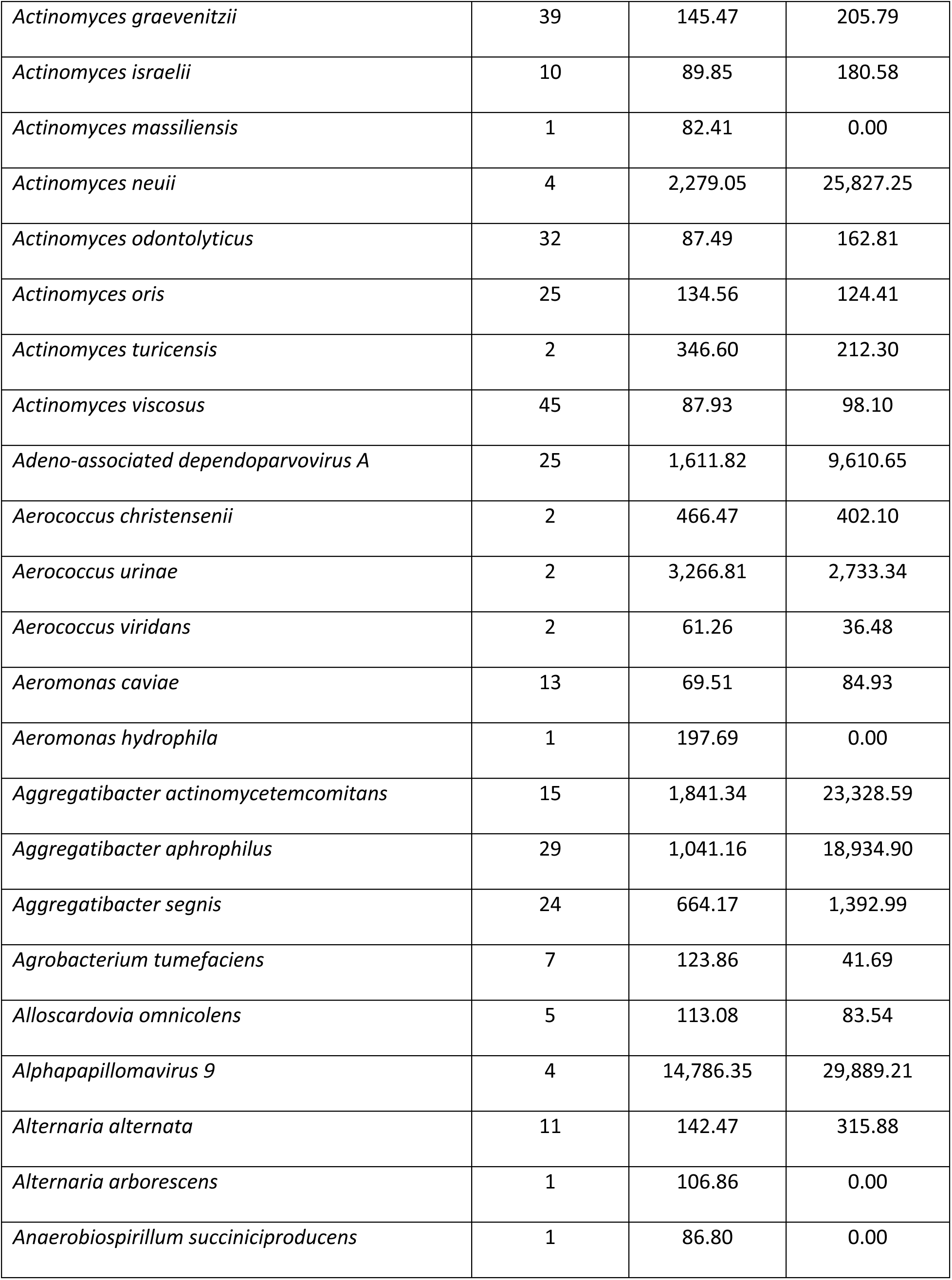
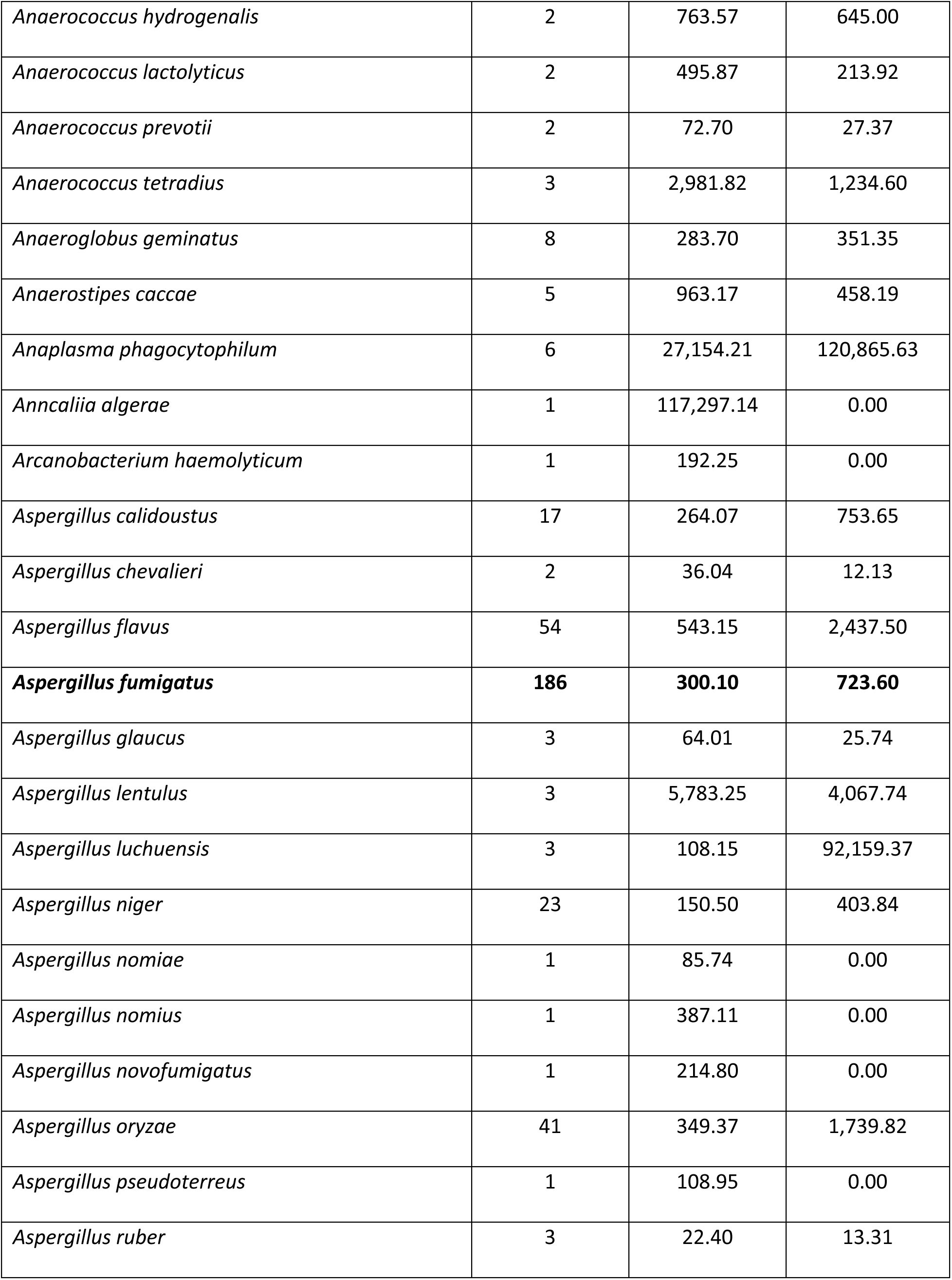
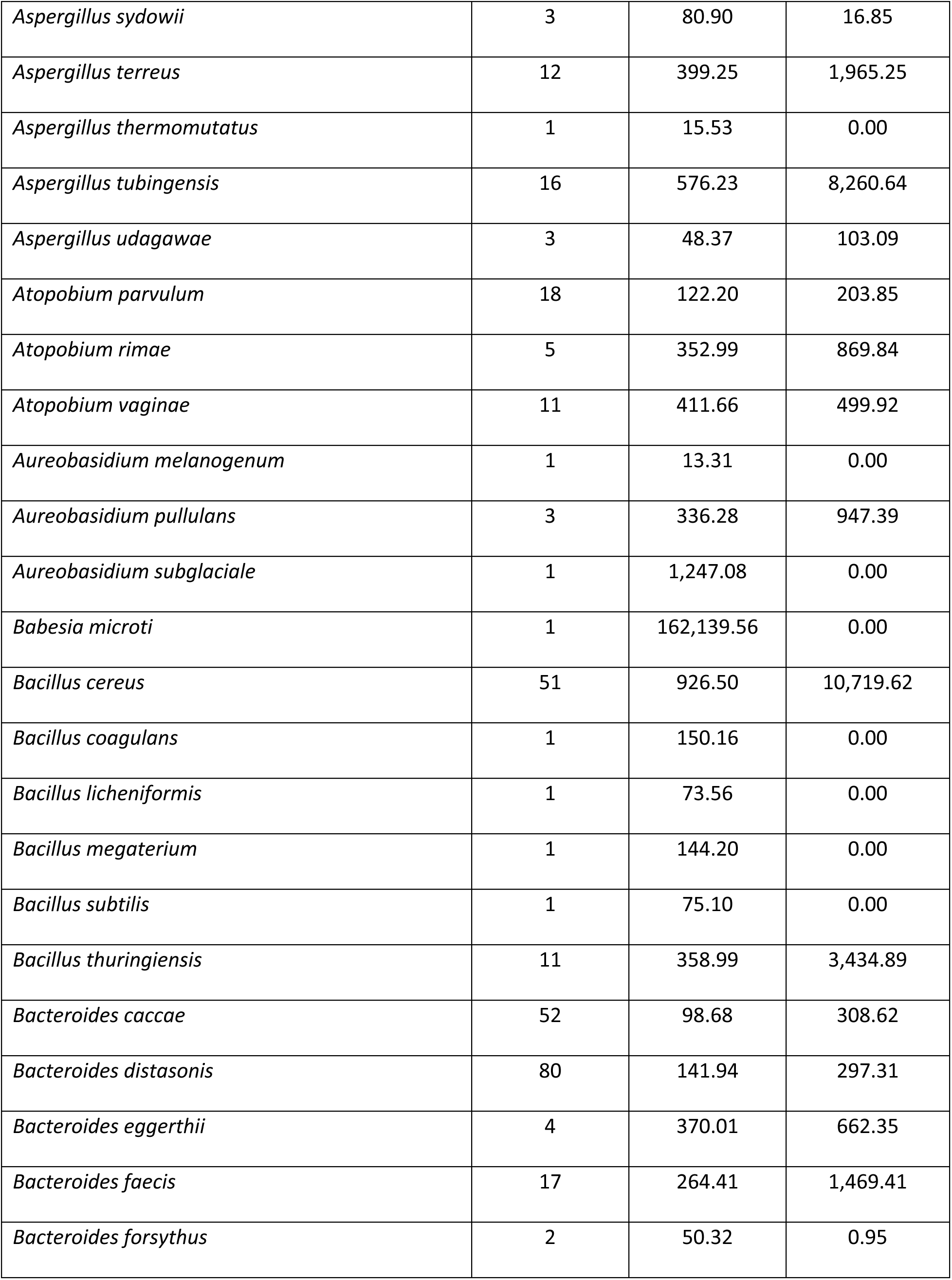
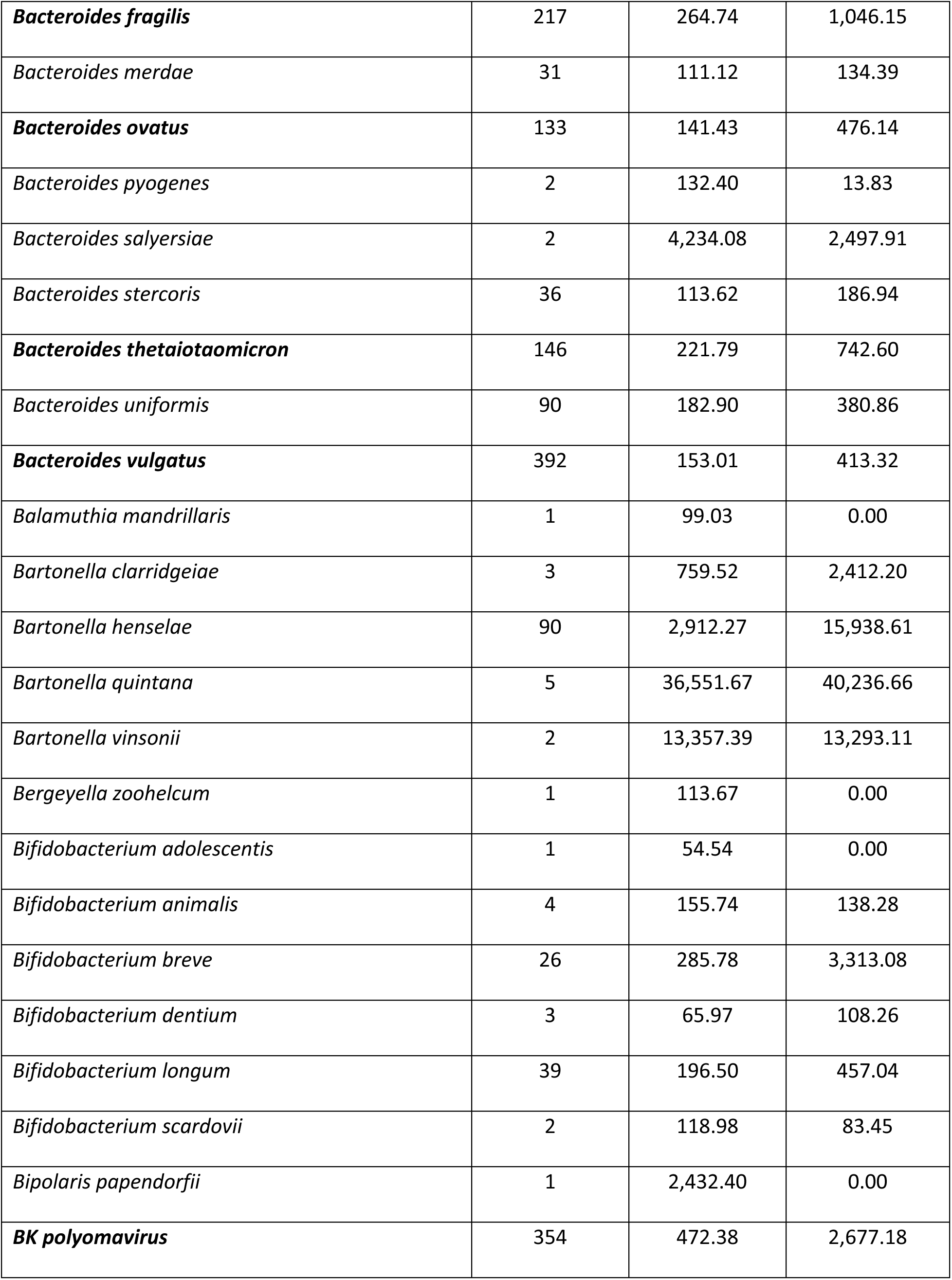
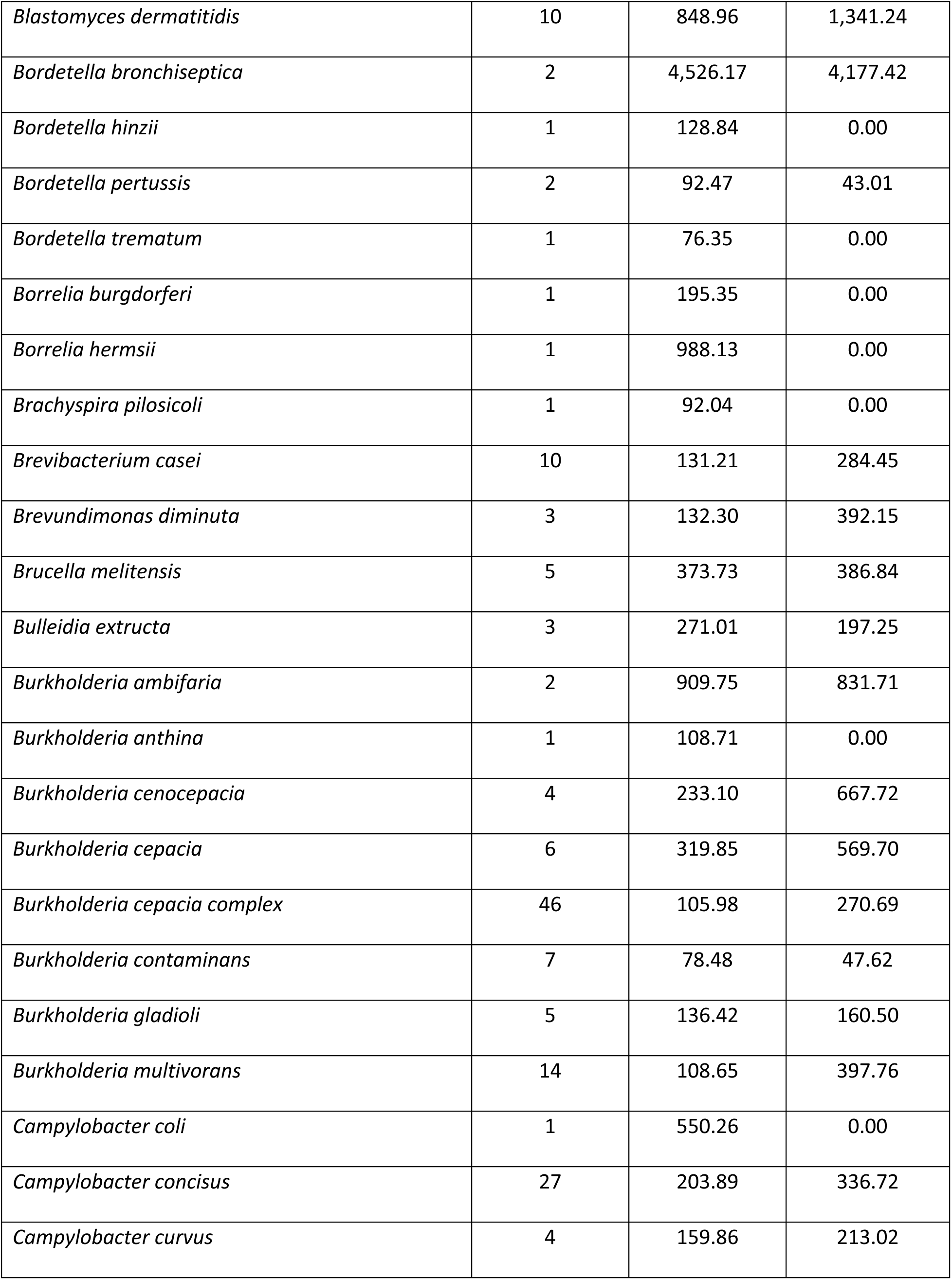
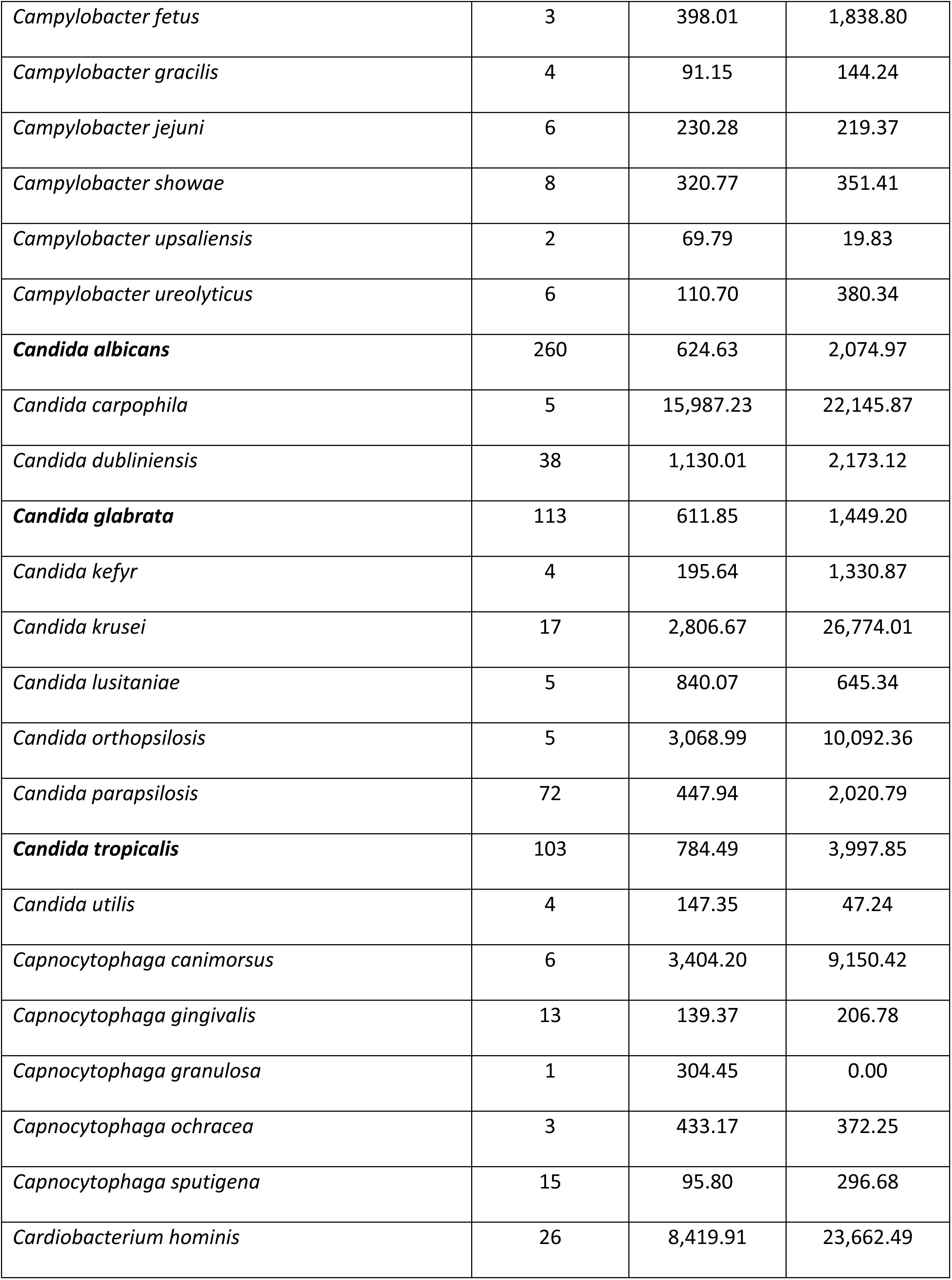
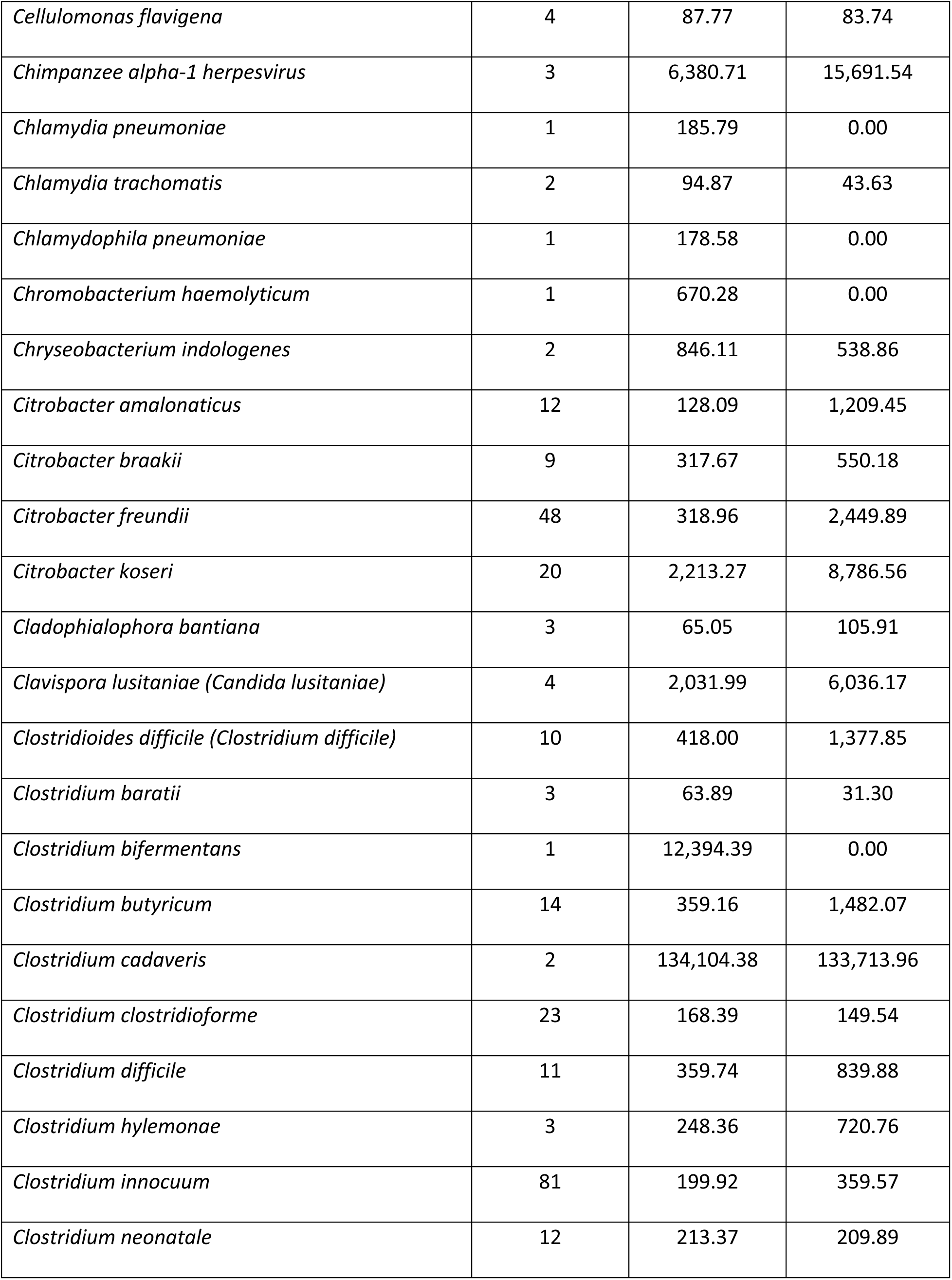
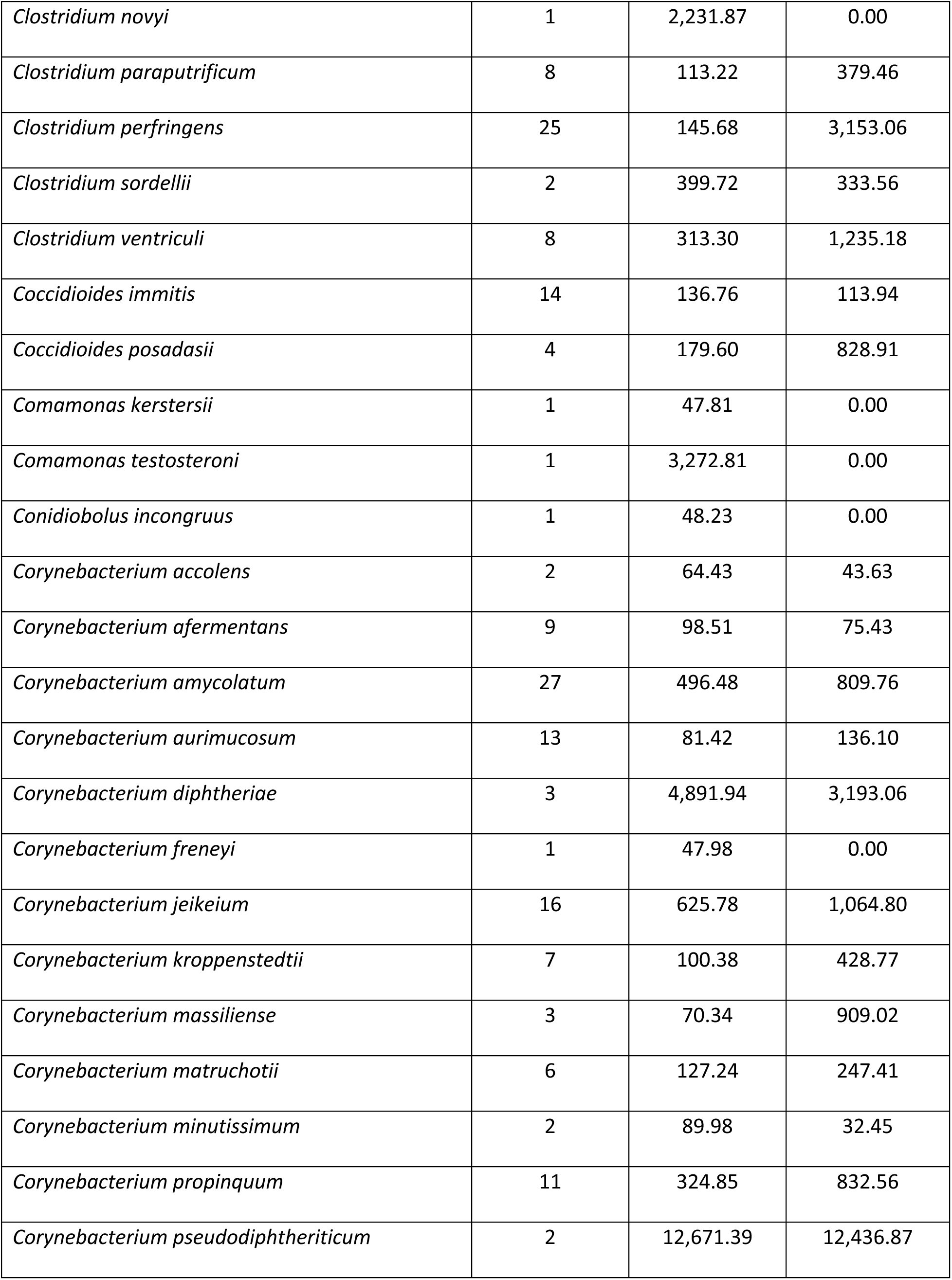
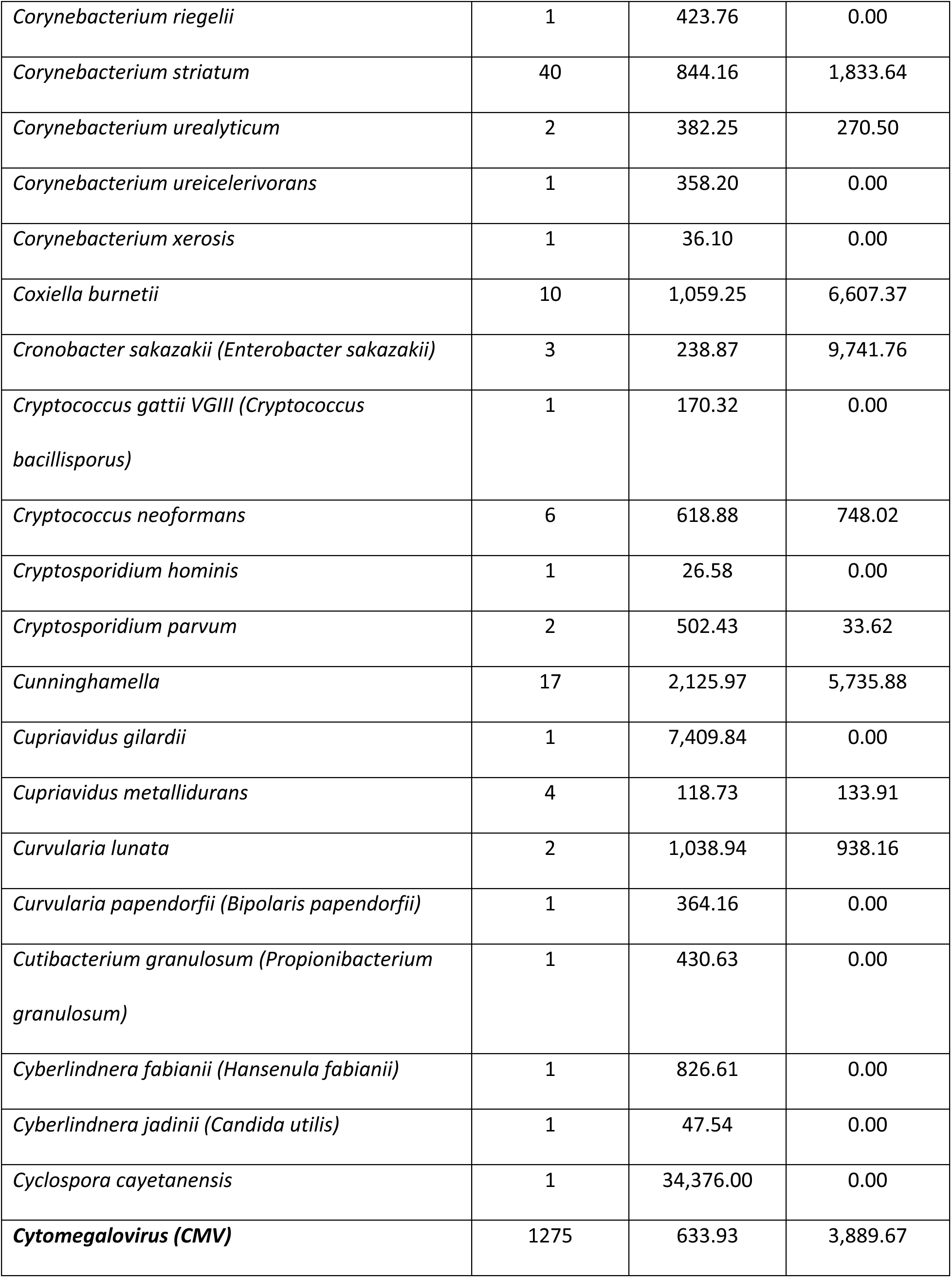
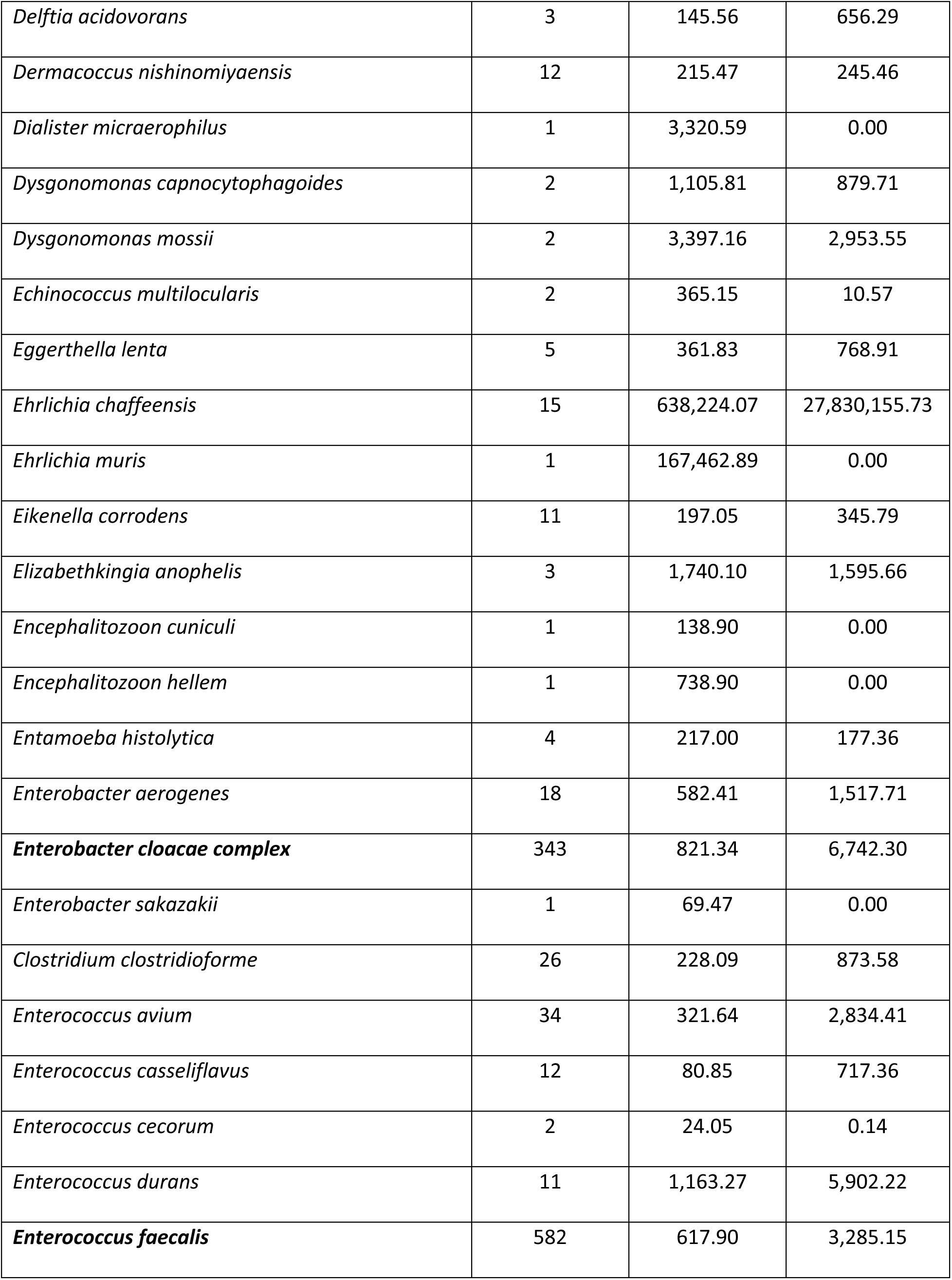
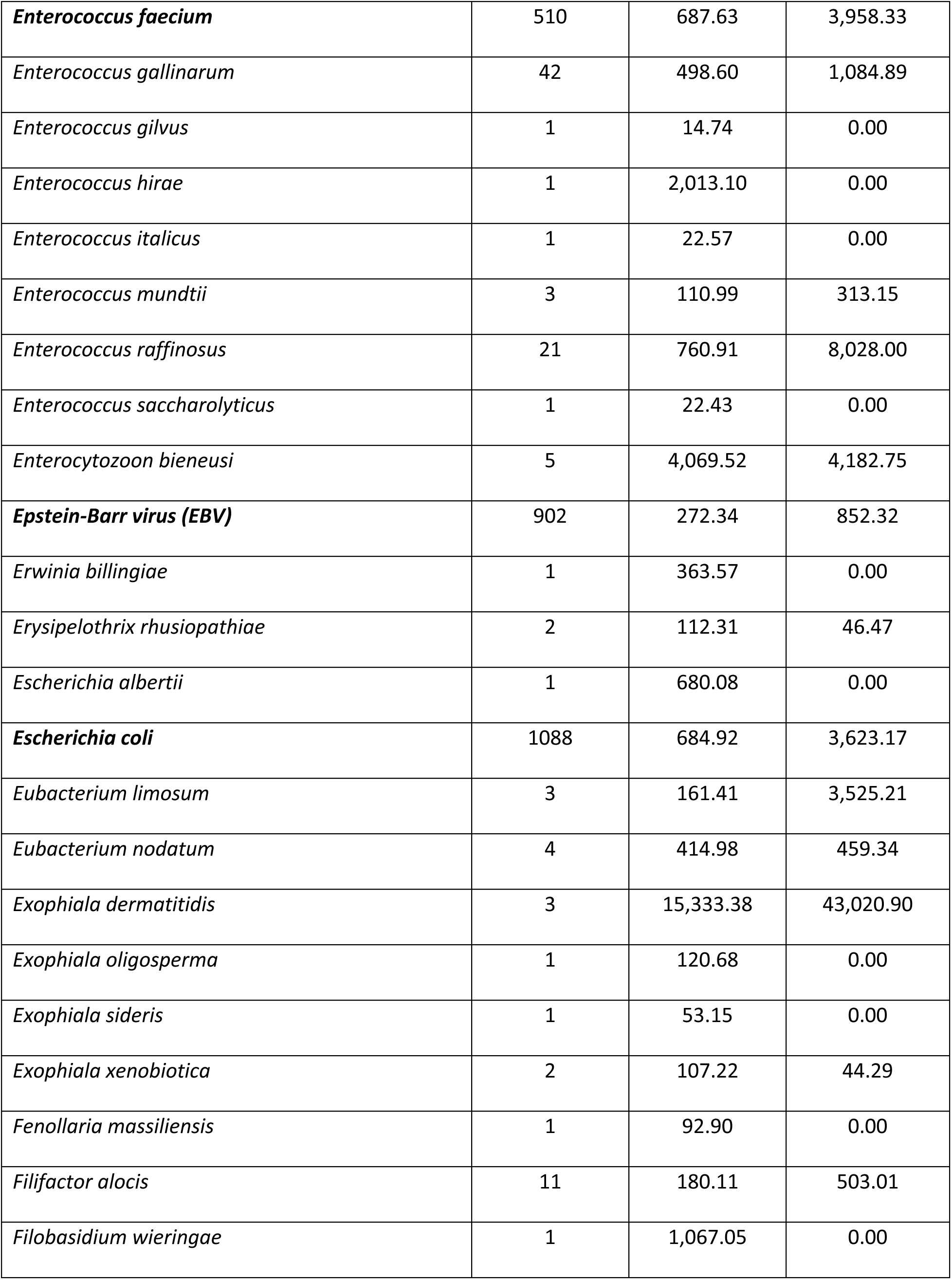
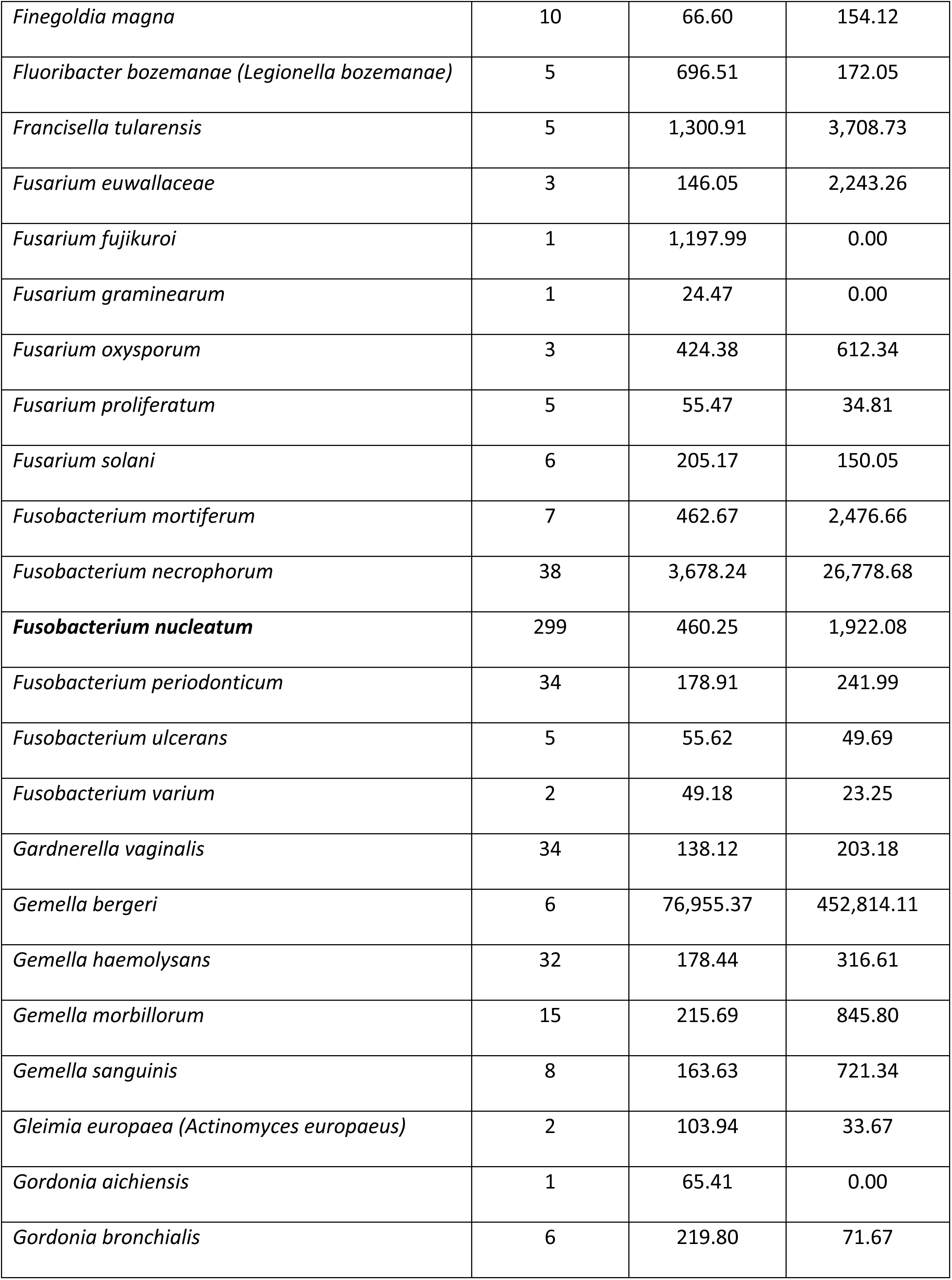
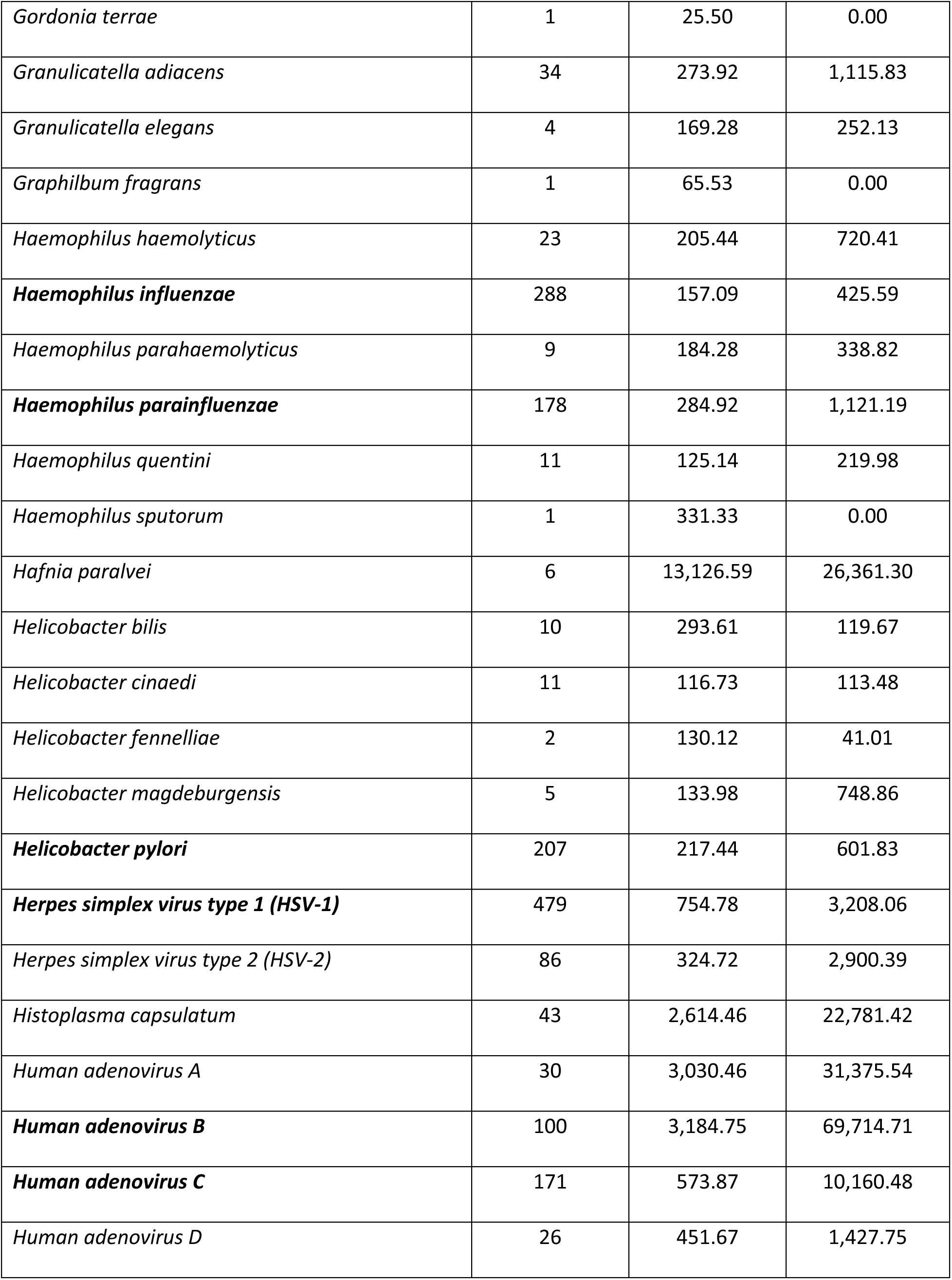
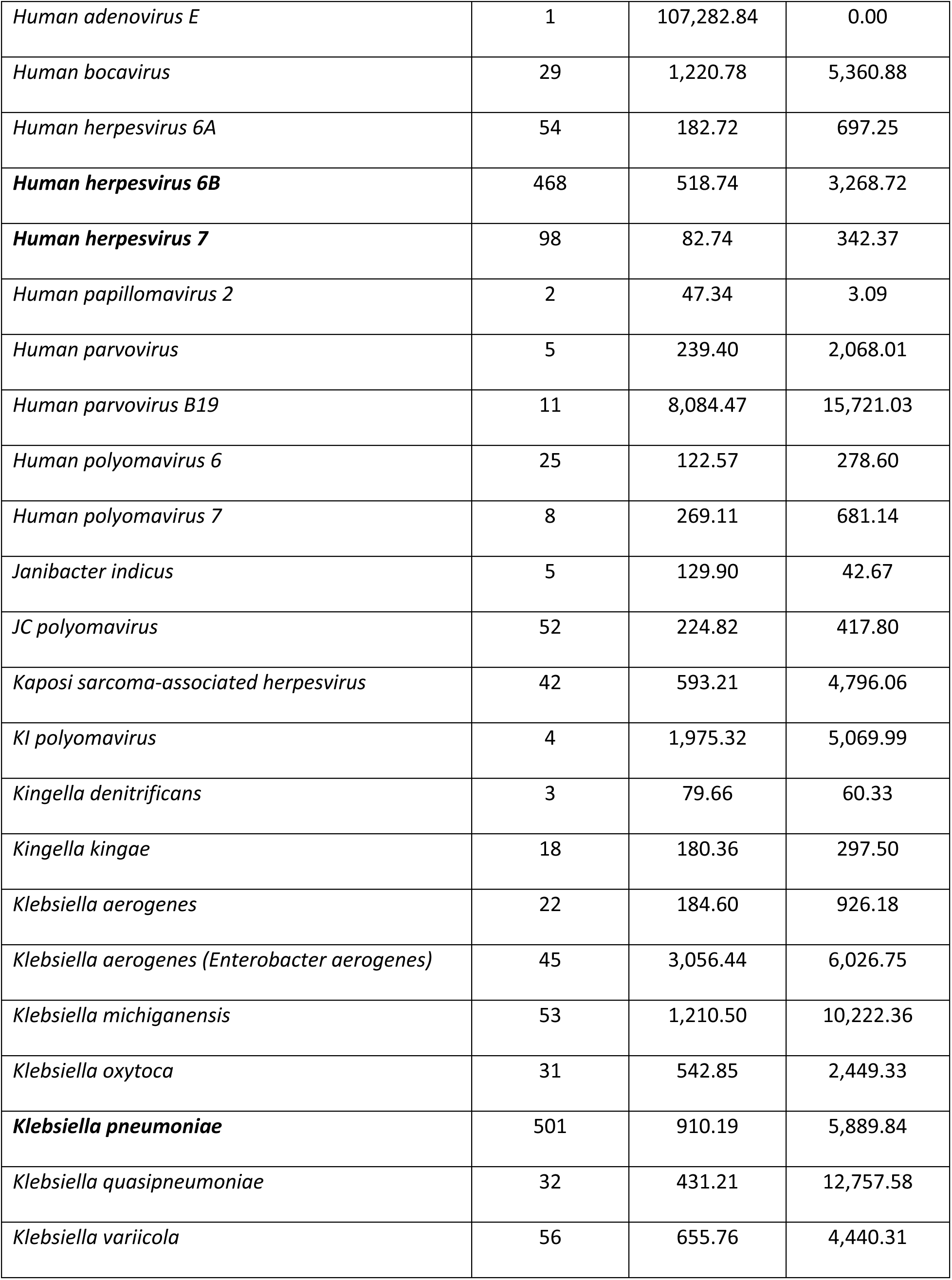
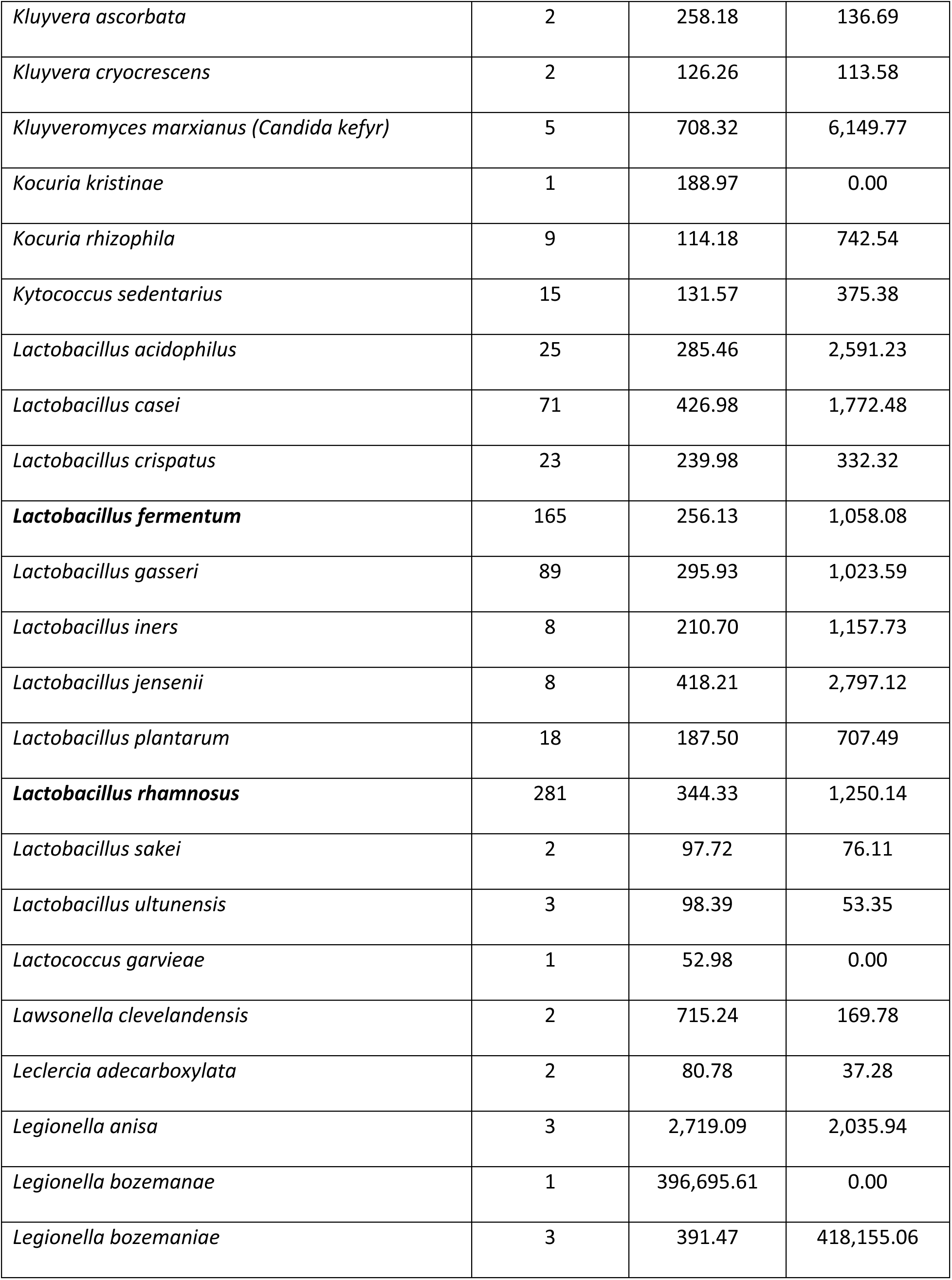
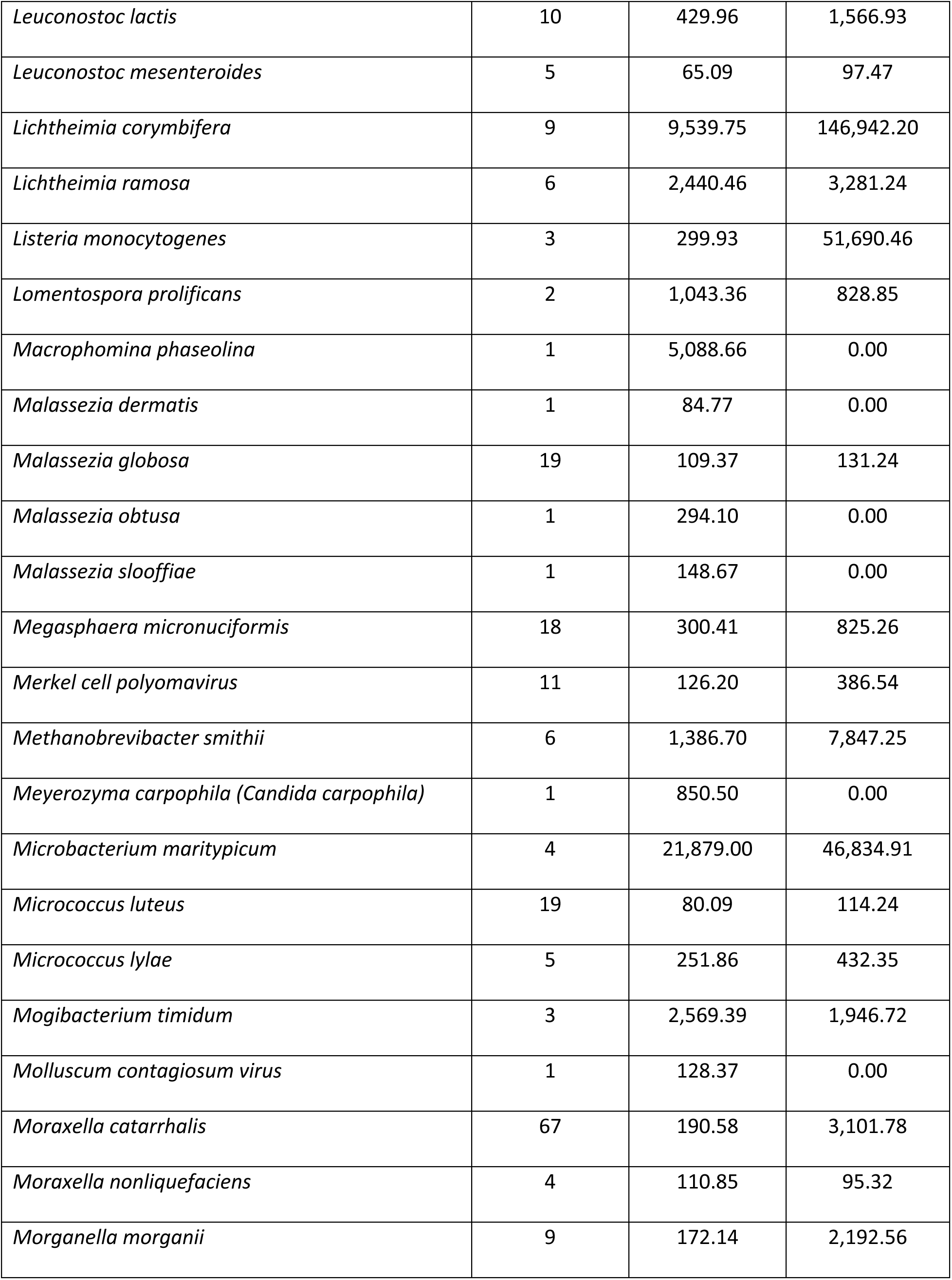
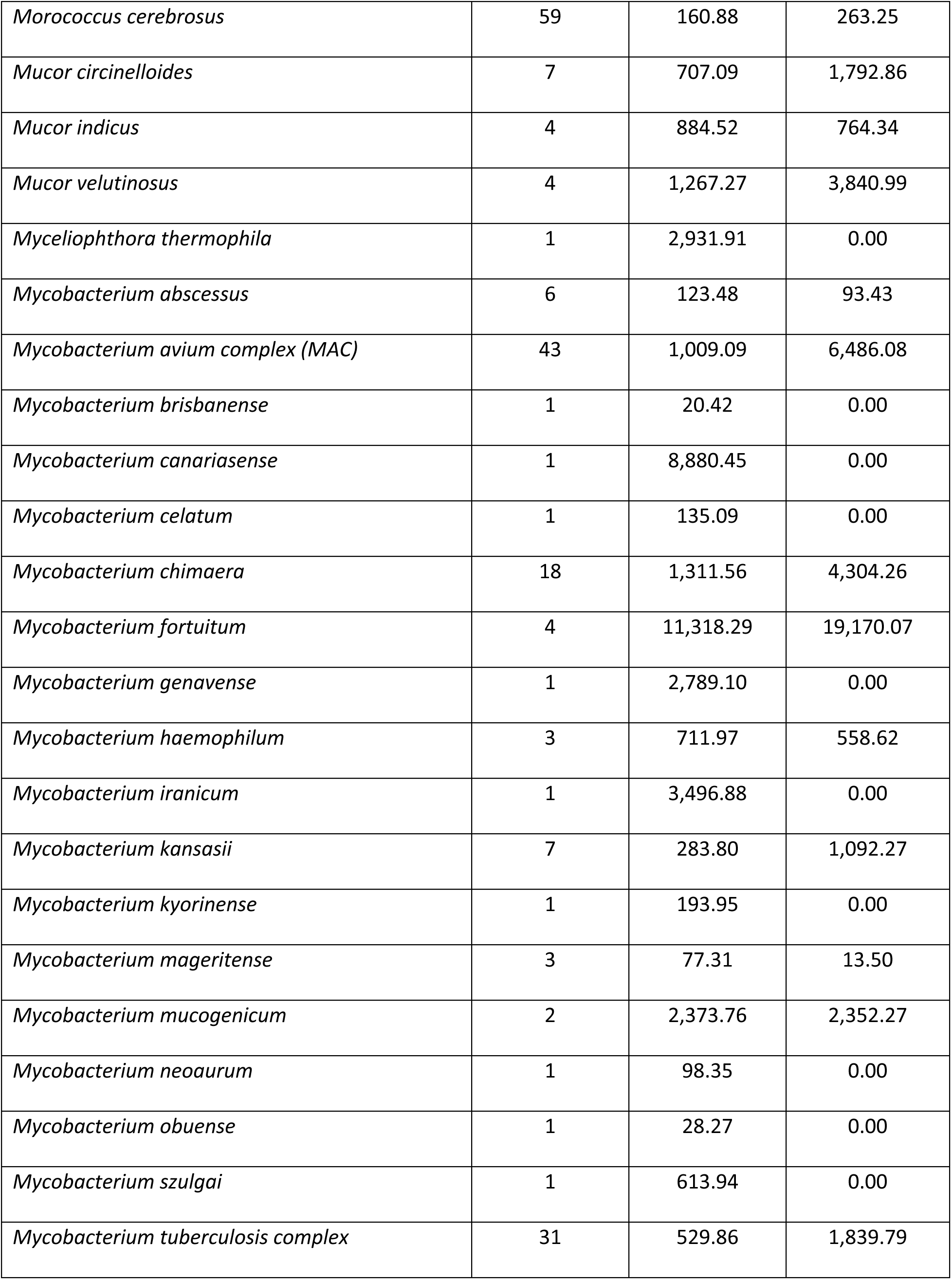
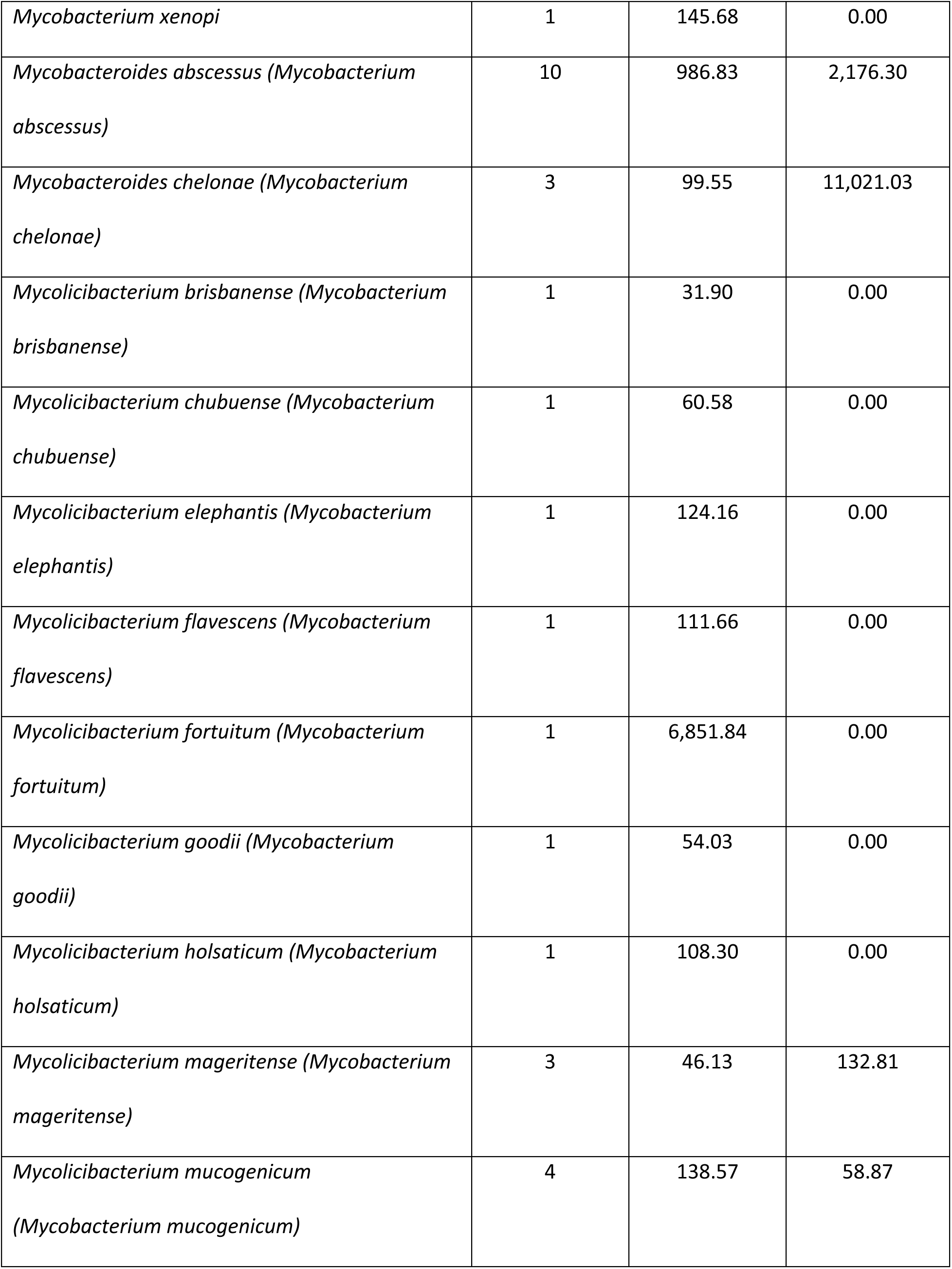
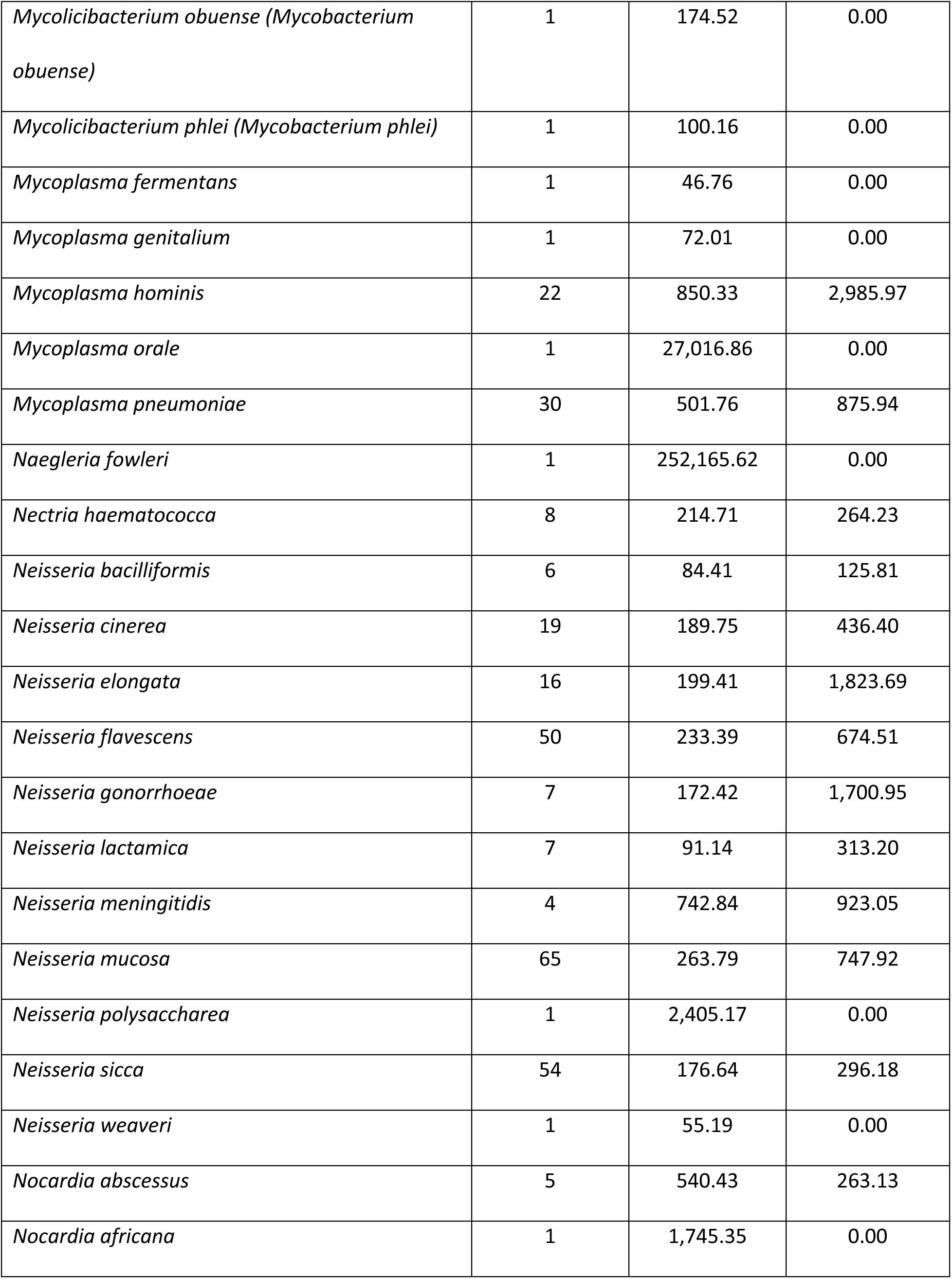
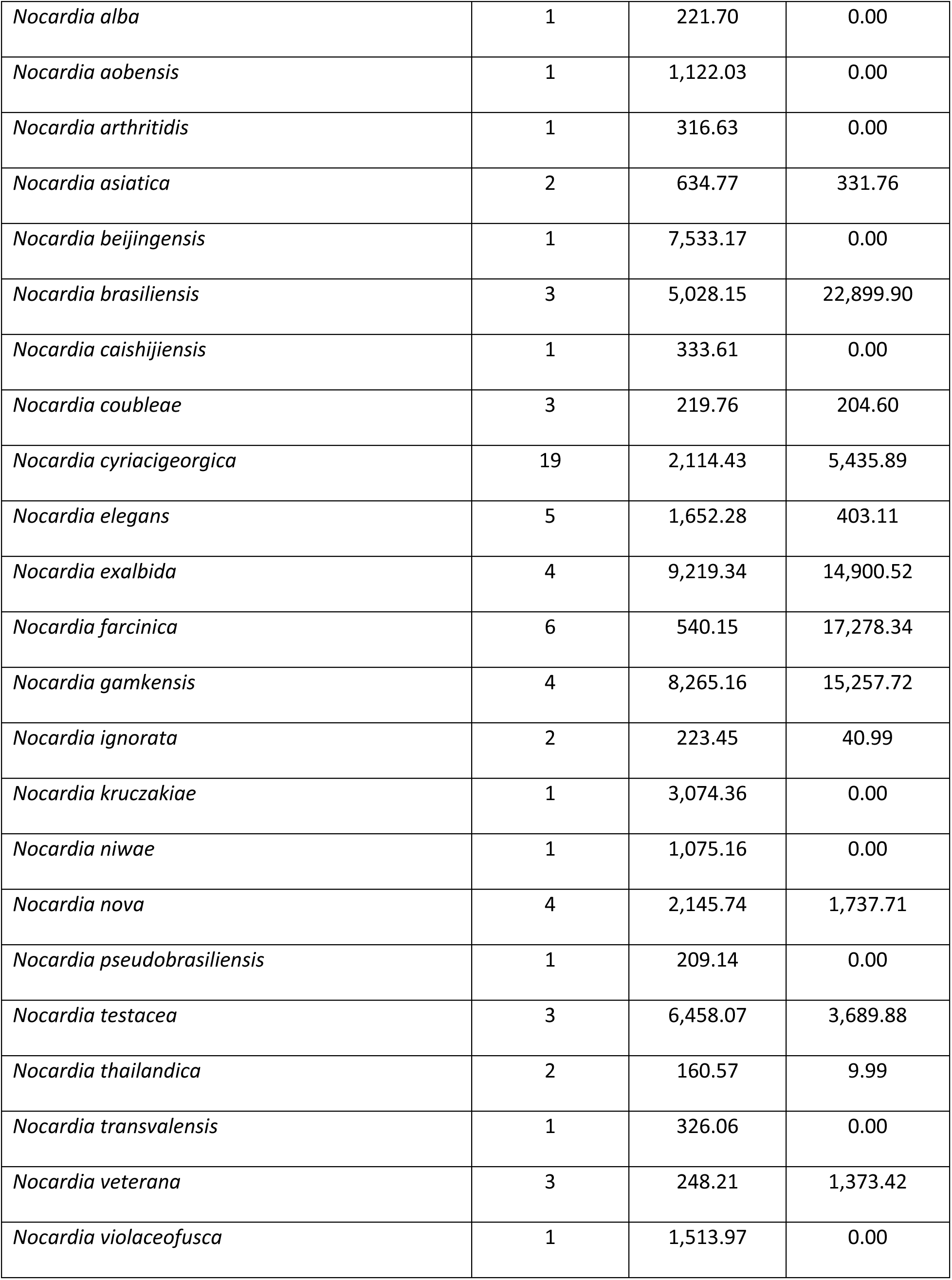
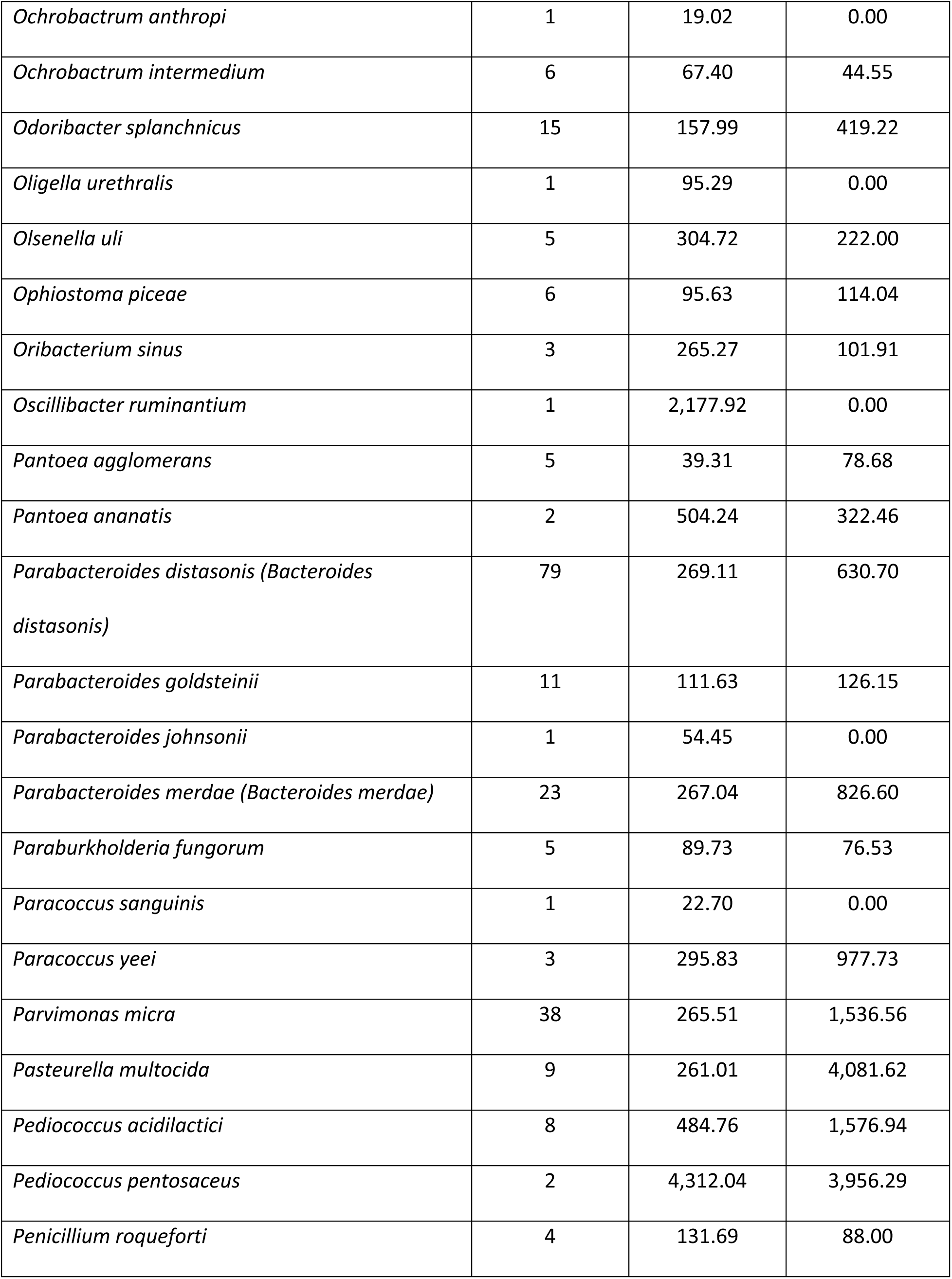
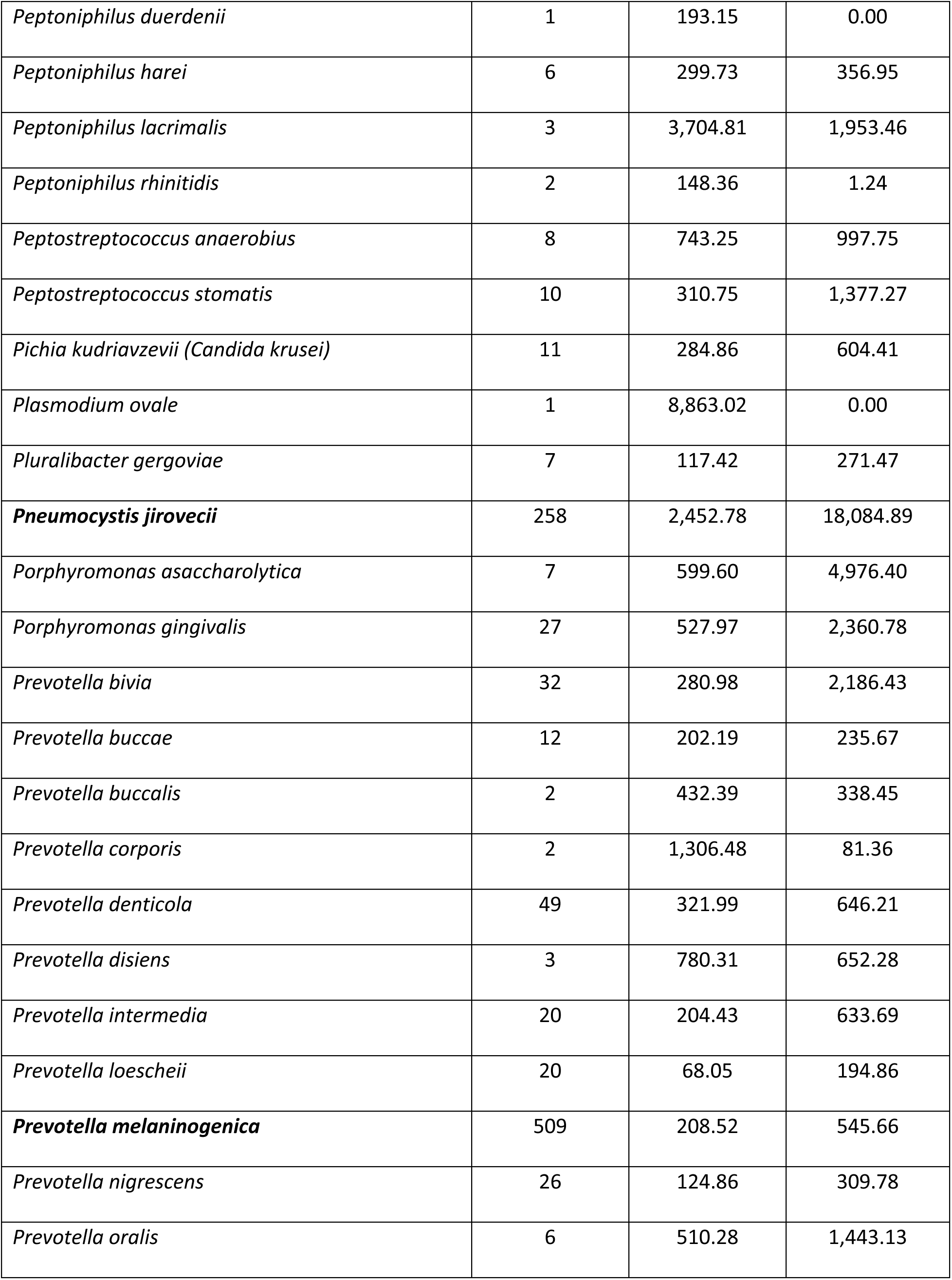
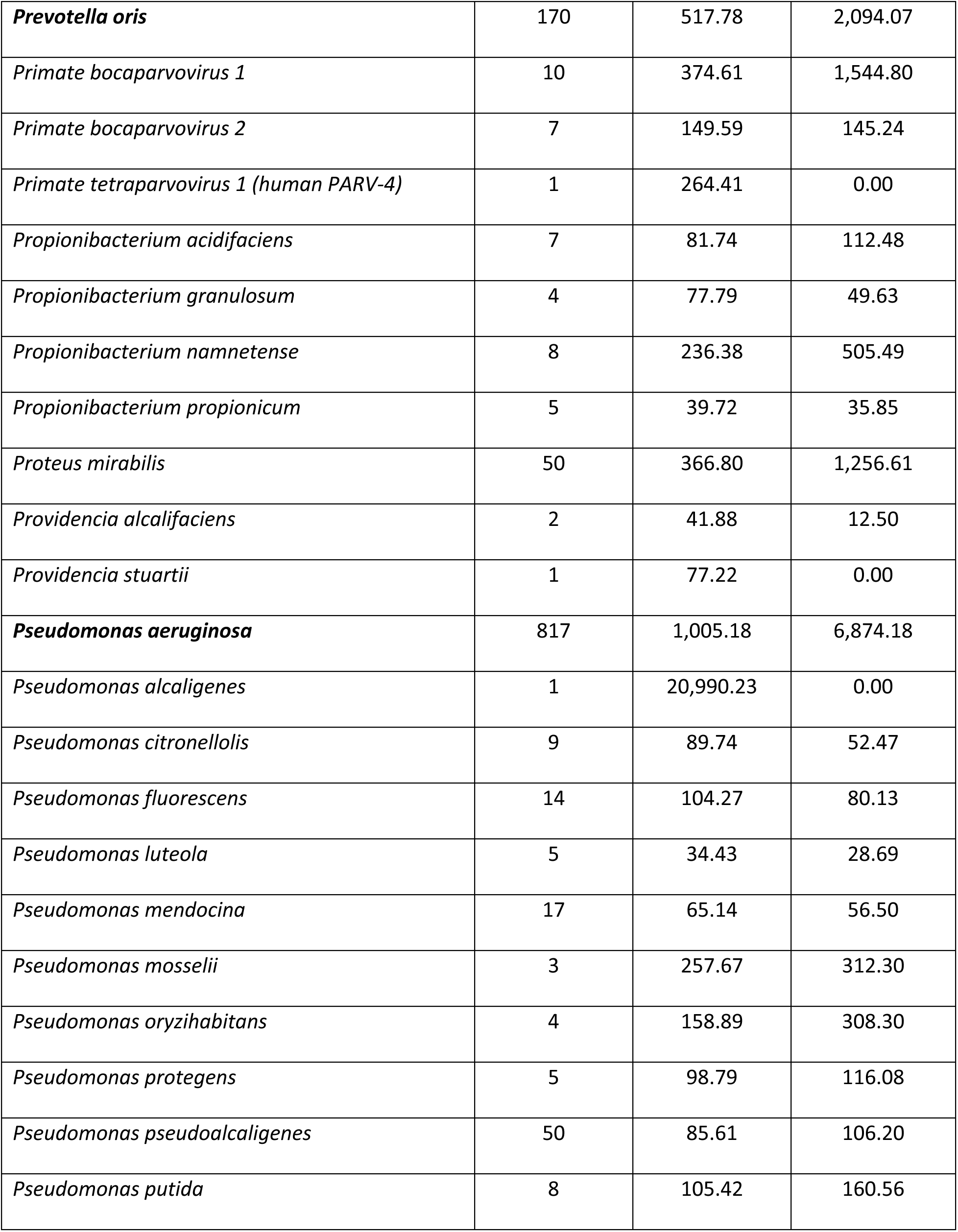
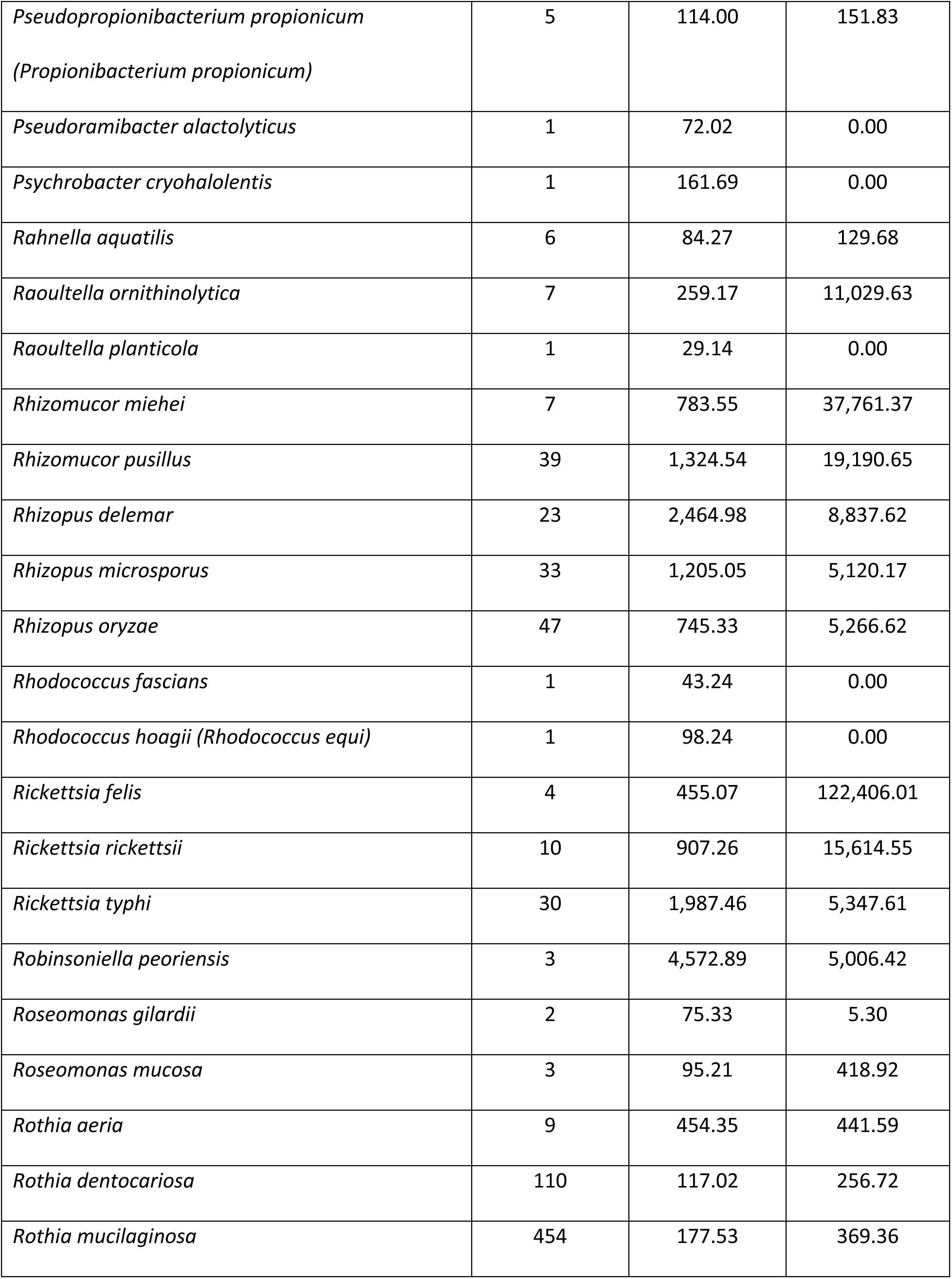
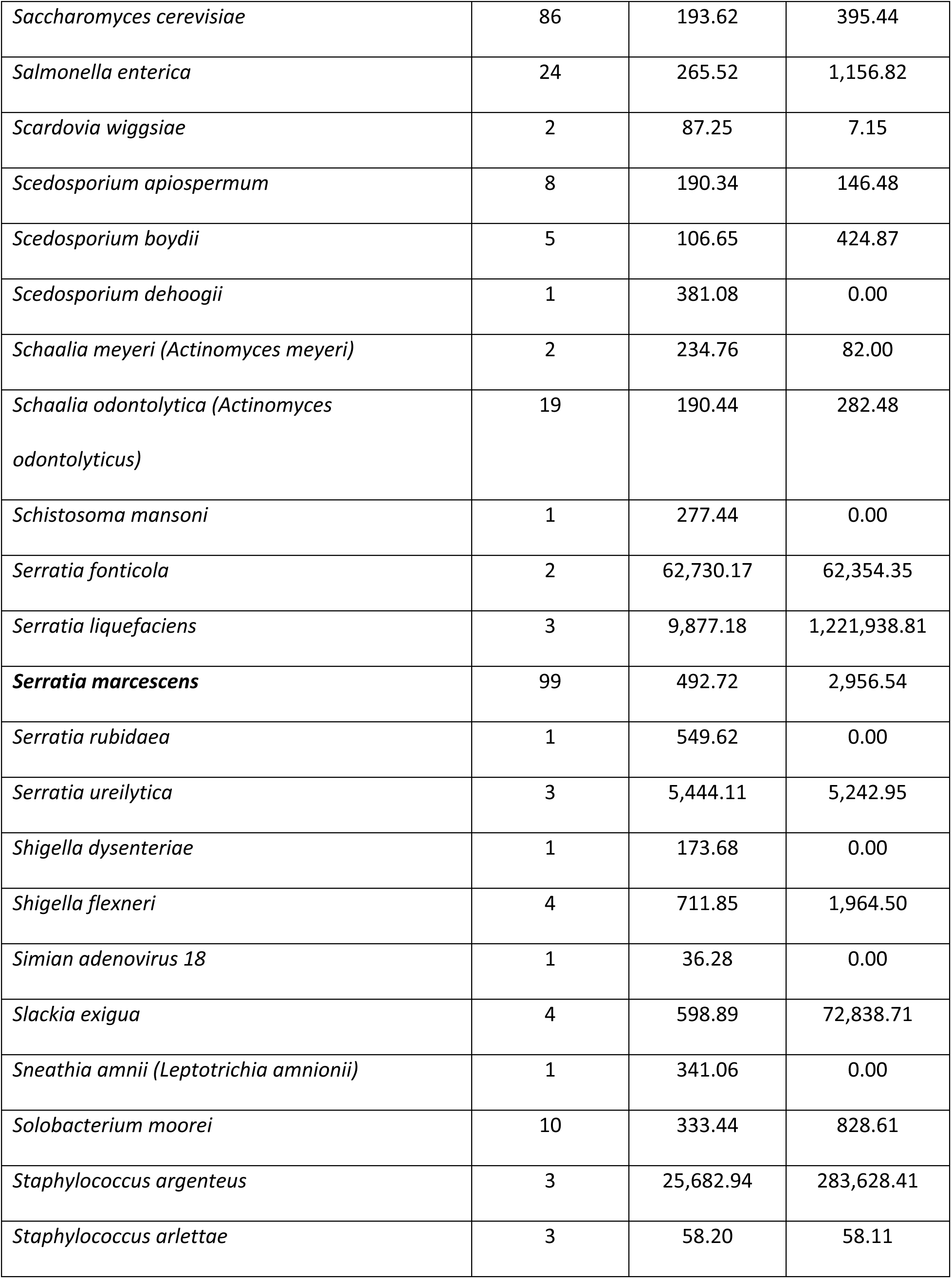
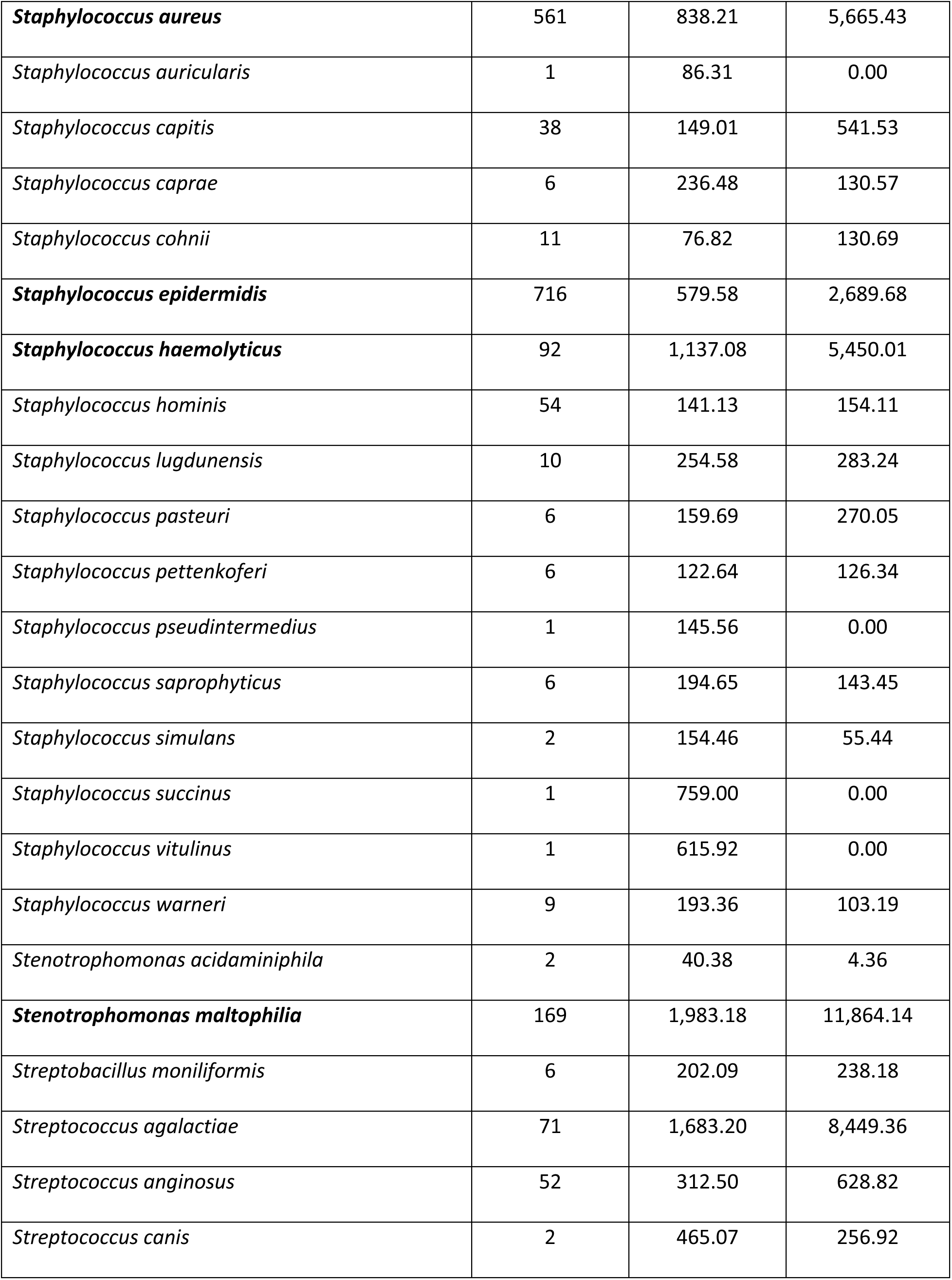
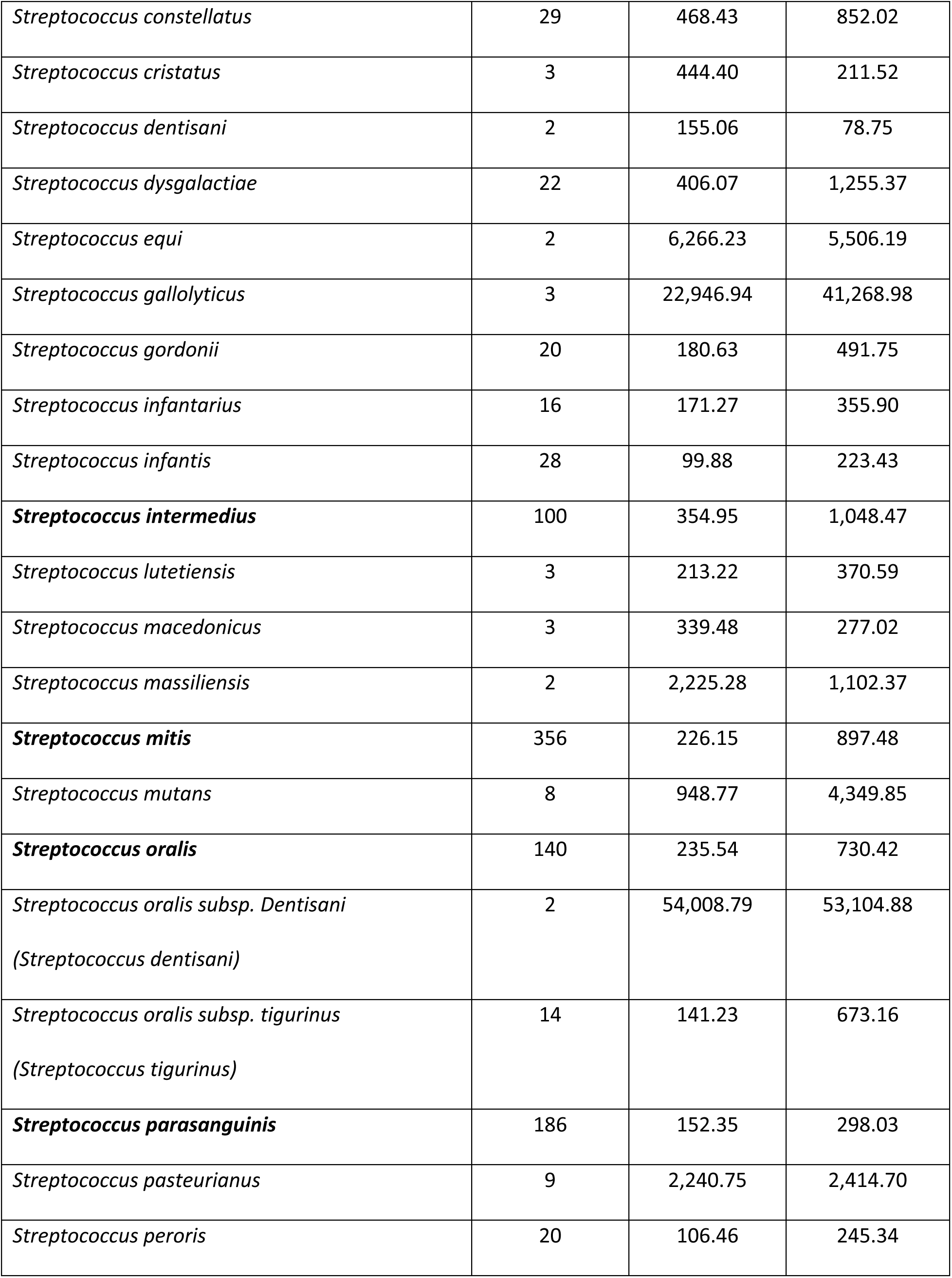
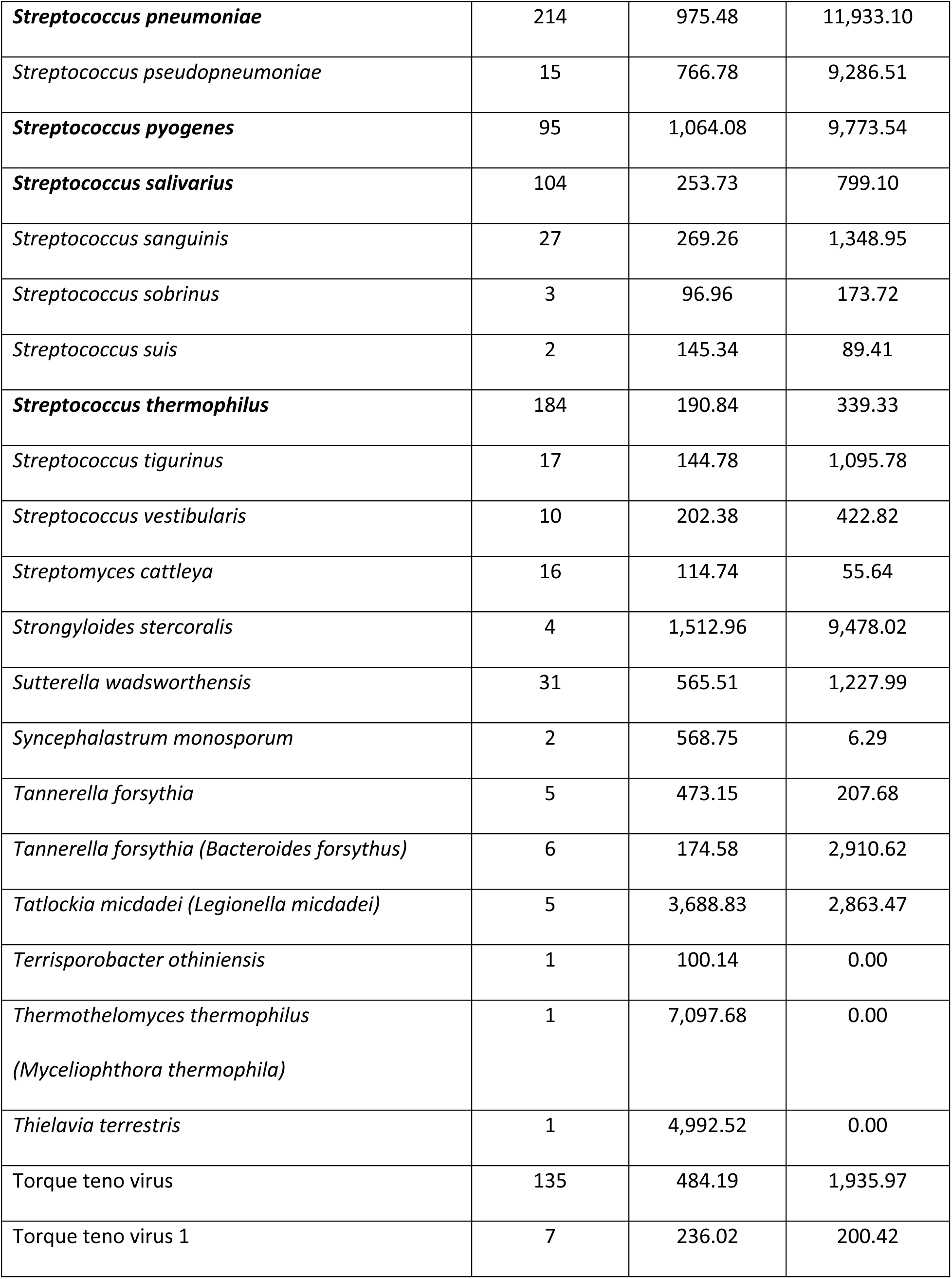
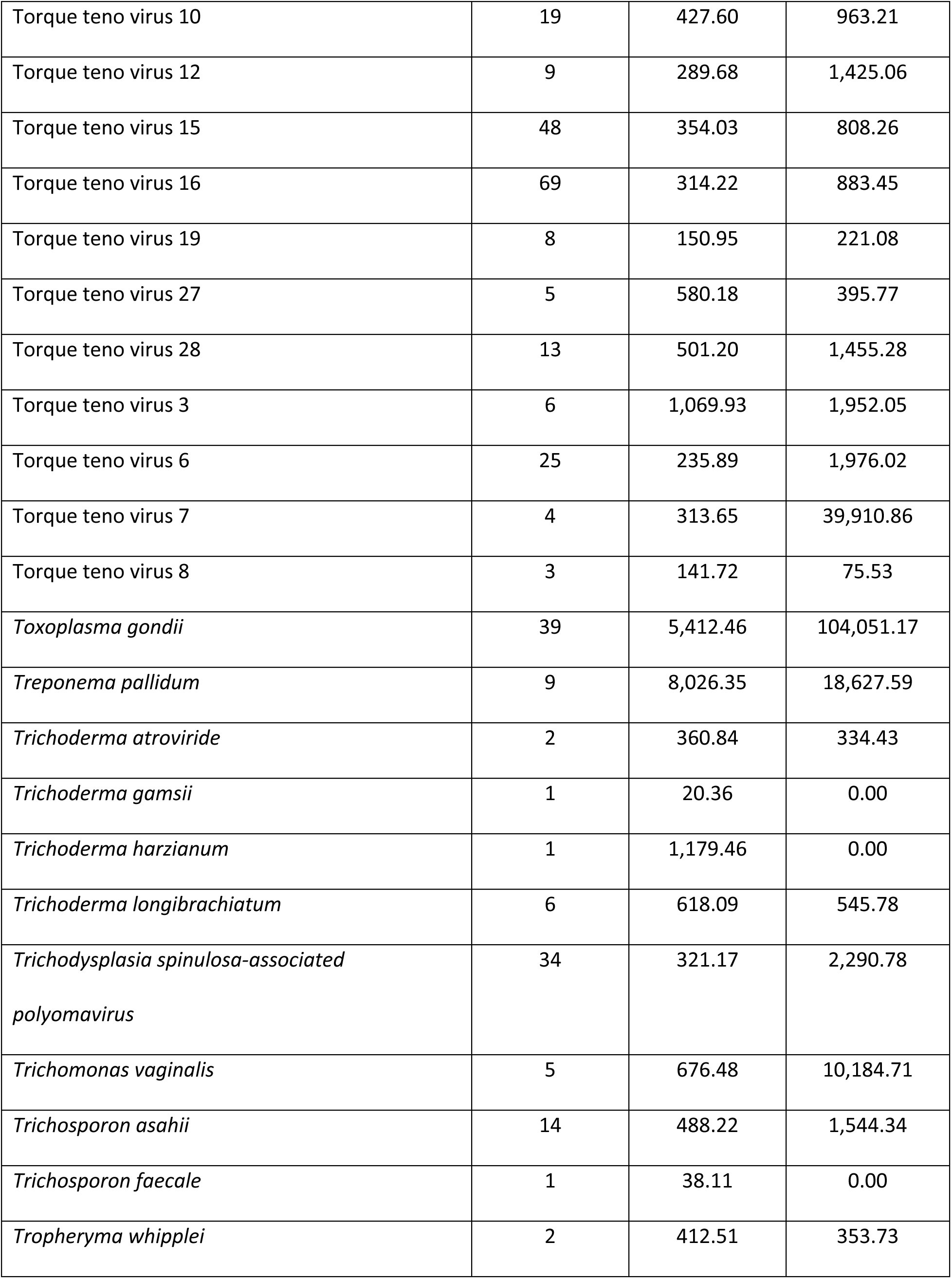
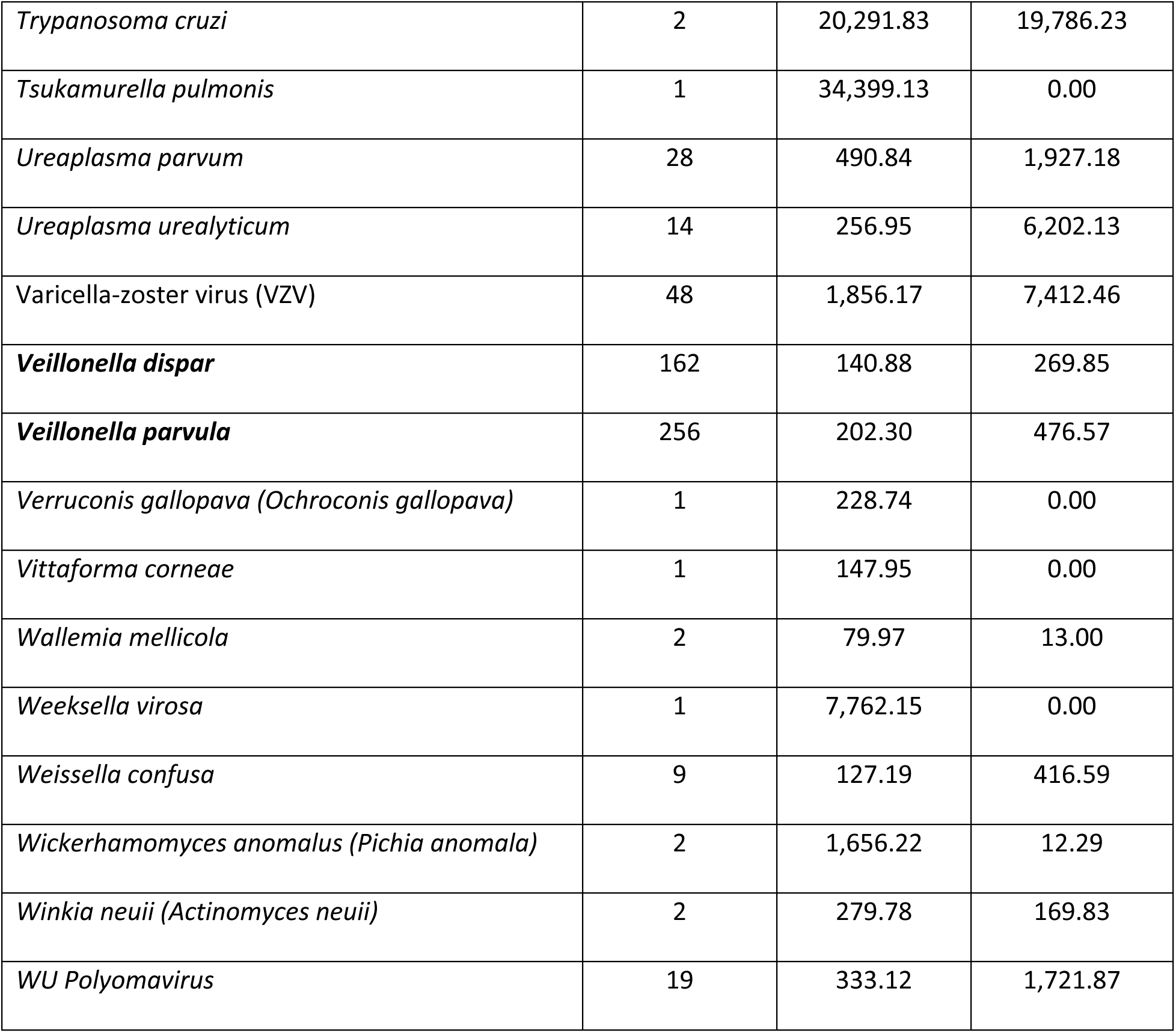
All 701 taxa detected in reports with number detected and median and interquartile range for quantification in molecules/µl (MPM) for each taxon, Apr 2018–Sept 2021. The top 50 detected are highlighted in **bold**.

**Supplemental Table 2.** Reports with co-detections of *Legionella* spp., *Nocardia* spp., and *Mycobacterium* spp., April 2018–September 2021.

| Sample number | Genus | Taxa name | MPM |
| --- | --- | --- | --- |
| 1 | *Legionella* spp. | *L. brunensis* | 401 |
|  |  | *L. hackeliae* | 270 |
|  |  | Epstein-Barr virus (EBV) | 1,405 |
| 2 | *Legionella* spp. | *L. feeleii* | 78,508 |
|  |  | *L. tunisiensis* | 76,445 |
|  |  | *Aspergillus fumigatus* | 594 |
|  |  | *Encephalitozoon hellem* | 739 |
| 3 | *Nocardia* spp. | *N. alba* | 222 |
|  |  | *N. caishijiensis* | 334 |
|  |  | *N. coubleae* | 495 |
|  |  | *N. ignorata* | 182 |
|  |  | *N. thailandica* | 171 |
|  |  | Human adenovirus D | 747 |
|  |  | *Prevotella melaninogenica* | 283 |
| 4 | *Nocardia* spp. | *N. exalbida* | 17,384 |
|  |  | *N. gamkensis* | 22,906 |
| 5 | *Nocardia* spp. | *N. exalbida* | 2,153 |
|  |  | *N. gamkensis* | 1,871 |
| 6 | *Nocardia* spp. | *N. exalbida* | 182 |
|  |  | *N. gamkensis* | 242 |
|  |  | *BK polyomavirus* | 81 |
|  |  | *Pseudomonas aeruginosa* | 498 |
| 7 | ***Nocardia* spp.** | ***N. elegans*** | 131 |
|  |  | ***N. nova*** | 440 |
|  |  | *Actinomyces oris* | 150 |
|  |  | *Bacteroides vulgatus* | 215 |
|  |  | *Corynebacterium striatum* | 1,590 |
|  |  | *Enterococcus faecalis* | 296 |
|  |  | *Lactobacillus fermentum* | 117 |
|  |  | *Lomentospora prolificans* | 215 |
|  |  | *Prevotella buccae* | 138 |
|  |  | *Prevotella oralis* | 209 |
|  |  | *Rothia mucilaginosa* | 874 |
|  |  | *Staphylococcus aureus* | 364 |
|  |  | *Streptococcus salivarius* | 1,032 |
|  |  | *Veillonella parvula* | 122 |
| 8 | ***Nocardia* spp.** | ***N. africana*** | 1,745 |
|  |  | ***N. elegans*** | 6,314 |
|  |  | ***N. nova*** | 3,945 |
|  |  | Cytomegalovirus (CMV) | 12,597 |
| 9 | ***Nocardia* spp.** | ***N. abscessus*** | 619 |
|  |  | ***N. arthritidis*** | 317 |
|  |  | ***N. asiatica*** | 303 |
| 10 | ***Nocardia* spp.** | ***N. elegans*** | 1,556 |
|  |  | ***N. nova*** | 2,720 |
| 11 | ***Nocardia* spp.** | ***N. elegans*** | 1,652 |
|  |  | ***N. nova*** | 1,572 |
| 12 | ***Nocardia* spp.** | ***N. exalbida*** | 16,286 |
|  |  | ***N. gamkensis*** | 14,660 |
|  |  | JC polyomavirus | 337 |
| 13 | ***Nocardia* spp.** | ***N. coubleae*** | 220 |
|  |  | ***N. ignorata*** | 264 |
|  |  | ***N. thailandica*** | 151 |
|  |  | Cytomegalovirus (CMV) | 81 |
|  |  | Human adenovirus D | 692 |
| 14 | ***Nocardia* spp.** | ***N. aobensis*** | 1,122 |
|  |  | ***N. kruczakiae*** | 3,074 |
|  |  | ***N. violaceofusca*** | 1,514 |
|  |  | *Aspergillus fumigatus* | 4,204 |
|  |  | BK polyomavirus | 2,260 |
|  |  | Epstein-Barr virus (EBV) | 49 |
|  |  | JC polyomavirus | 61 |
| 15 | ***Mycobacterium* spp.** | ***M. brisbanense*** | 20 |
|  |  | ***M. mucogenicum*** | 21 |
|  |  | ***M. obuense*** | 28 |
|  |  | *Stenotrophomonas acidaminiphila* | 36 |
| 16 | **Mycobacterium spp.** | ***M. chubuense*** | 61 |
|  |  | ***M. elephantis*** | 124 |
|  |  | ***M. flavescens*** | 112 |
|  |  | ***M. goodii*** | 54 |
|  |  | ***M. holsaticum*** | 108 |
|  |  | ***M. phlei*** | 100 |
|  |  | *Neisseria meningitidis* | 62 |
| 17 | **Mycobacterium spp.** | ***M. avium complex (MAC)*** | 441 |
|  |  | ***M. celatum*** | 135 |
|  |  | ***M. kyorinense*** | 194 |
|  |  | Epstein-Barr virus (EBV) | 155 |
|  |  | *Prevotella melaninogenica* | 398 |
|  |  | *Rothia mucilaginosa* | 913 |
|  |  | *Veillonella dispar* | 236 |
|  |  | *Veillonella parvula* | 122 |
| 18 | **Mycobacterium spp.** | ***M. avium complex (MAC)*** | 8,039 |
|  |  | ***M. chimaera*** | 2,410 |
| 19 | **Mycobacterium spp.** | ***M. avium complex (MAC)*** | 104 |
|  |  | ***M. chimaera*** | 104 |
| 20 | **Mycobacterium spp.** | ***M. complex (MAC)*** | 36,867 |
|  |  | ***M. chimaera*** | 36,862 |

## REFERENCES

1. Levy SE, Myers RM. 2016. Advancements in Next-Generation Sequencing. Annu Rev Genomics Hum Genet 17:95–115.

2. Altschul SF, Gish W, Miller W, Myers EW, Lipman DJ. 1990. Basic local alignment search tool. J Mol Biol 215:403–10.

3. Kowarsky M, Camunas-Soler J, Kertesz M, De Vlaminck I, Koh W, Pan W, Martin L, Neff NF, Okamoto J, Wong RJ, Kharbanda S, El-Sayed Y, Blumenfeld Y, Stevenson DK, Shaw GM, Wolfe ND, Quake SR. 2017. Numerous uncharacterized and highly divergent microbes which colonize humans are revealed by circulating cell-free DNA. Proc Natl Acad Sci U S A 114:9623–9628.

4. Lewinski M, Alby K, Babady E, Butler-Wu S, Dien Bard J, Greninger A, Hanson K, Naccache S, Newton D, Temple-Smolkin RL, Nolte F. *in press*. Exploring the Utility of Multiplex Infectious Disease Panel Testing for Diagnosis of Infection in Different Body Sites: A Joint Report of the Association for Molecular Pathology, American Society for Microbiology, Infectious Diseases Society of America, and Pan American Society for Clinical Virology. The Journal of Molecular Diagnostics.

5. Naureckas Li C, Nakamura MM. 2022. Utility of Broad-Range PCR Sequencing for Infectious Diseases Clinical Decision Making: a Pediatric Center Experience. J Clin Microbiol 60:e0243721.

6. Whiting PF, Rutjes AW, Westwood ME, Mallett S, Deeks JJ, Reitsma JB, Leeflang MM, Sterne JA, Bossuyt PM, Group Q-. 2011. QUADAS-2: a revised tool for the quality assessment of diagnostic accuracy studies. Ann Intern Med 155:529–36.

7. Liu J, Zhang Q, Dong YQ, Yin J, Qiu YQ. 2022. Diagnostic accuracy of metagenomic next-generation sequencing in diagnosing infectious diseases: a meta-analysis. Sci Rep 12:21032.

8. Eichenberger EM, Degner N, Scott ER, Ruffin F, Franzone J, Sharma-Kuinkel B, Shah P, Hong D, Dalai SC, Blair L, Hollemon D, Chang E, Ho C, Wanda L, de Vries C, Fowler VG, Ahmed AA. 2022. Microbial Cell-Free DNA Identifies the Causative Pathogen in Infective Endocarditis and Remains Detectable Longer Than Conventional Blood Culture in Patients with Prior Antibiotic Therapy. Clin Infect Dis doi:10.1093/cid/ciac426.

9. Dworsky ZD, Lee B, Ramchandar N, Rungvivatjarus T, Coufal NG, Bradley JS. 2022. Impact of Cell-Free Next-Generation Sequencing on Management of Pediatric Complicated Pneumonia. Hosp Pediatr 12:377–384.

10. Brenner T, Skarabis A, Stevens P, Axnick J, Haug P, Grumaz S, Bruckner T, Luntz S, Witzke O, Pletz MW, Ruprecht TM, Marschall U, Altin S, Greiner W, Berger MM, Group TICCT. 2021. Optimization of sepsis therapy based on patient-specific digital precision diagnostics using next generation sequencing (DigiSep-Trial)-study protocol for a randomized, controlled, interventional, open-label, multicenter trial. Trials 22:714.

11. Han D, Li R, Shi J, Tan P, Zhang R, Li J. 2020. Liquid biopsy for infectious diseases: a focus on microbial cell-free DNA sequencing. Theranostics 10:5501–5513.

12. Blauwkamp TA, Thair S, Rosen MJ, Blair L, Lindner MS, Vilfan ID, Kawli T, Christians FC, Venkatasubrahmanyam S, Wall GD, Cheung A, Rogers ZN, Meshulam-Simon G, Huijse L, Balakrishnan S, Quinn JV, Hollemon D, Hong DK, Vaughn ML, Kertesz M, Bercovici S, Wilber JC, Yang S. 2019. Analytical and clinical validation of a microbial cell-free DNA sequencing test for infectious disease. Nat Microbiol 4:663–674.

13. Rossoff J, Chaudhury S, Soneji M, Patel SJ, Kwon S, Armstrong A, Muller WJ. 2019. Noninvasive Diagnosis of Infection Using Plasma Next-Generation Sequencing: A Single-Center Experience. Open Forum Infect Dis 6.

14. Benamu E, Gajurel K, Anderson JN, Lieb T, Gomez CA, Seng H, Aquino R, Hollemon D, Hong DK, Blauwkamp TA, Kertesz M, Blair L, Bollyky PL, Medeiros BC, Coutre S, Zompi S, Montoya JG, Deresinski S. 2021. Plasma Microbial Cell-free DNA Next Generation Sequencing in the Diagnosis and Management of Febrile Neutropenia. Clin Infect Dis doi:10.1093/cid/ciab324.

15. Yu J, Diaz JD, Goldstein SC, Patel RD, Varela JC, Reyenga C, Smith M, Smith T, Balls J, Ahmad S, Mori S. 2021. Impact of Next-Generation Sequencing Cell-free Pathogen DNA Test on Antimicrobial Management in Adults with Hematological Malignancies and Transplant Recipients with Suspected Infections. Transplant Cell Ther doi:10.1016/j.jtct.2021.02.025.

16. Francisco DMA, Woc-colburn L, Carlson TJ, Lasco T, Barrett MB, Mohajer MA. 2020. 680. The use of plasma next-generation sequencing test in the management of immunocompetent and immunocompromised patients – a single center retrospective study. Open Forum Infectious Diseases 7:S393–S394.

17. Hogan CA, Yang S, Garner OB, Green DA, Gomez CA, Dien Bard J, Pinsky BA, Banaei N. 2021. Clinical Impact of Metagenomic Next-Generation Sequencing of Plasma Cell-Free DNA for the Diagnosis of Infectious Diseases: A Multicenter Retrospective Cohort Study. Clin Infect Dis 72:239–245.

18. Lee RA, Al Dhaheri F, Pollock NR, Sharma TS. 2020. Assessment of the Clinical Utility of Plasma Metagenomic Next-Generation Sequencing in a Pediatric Hospital Population. J Clin Microbiol 58.

19. Niles DT, Wijetunge DSS, Palazzi DL, Singh IR, Revell PA. 2020. Plasma Metagenomic Next-Generation Sequencing Assay for Identifying Pathogens: a Retrospective Review of Test Utilization in a Large Children’s Hospital. J Clin Microbiol 58.

20. National Center for Health Statistics (CDC). 2015. International Classification of Diseases, Tenth Revision, Clinical Modification (ICD-10-CM). https://ftp.cdc.gov/pub/Health_Statistics/NCHS/Publications/ICD10CM/2022/icd10cm-tabular-2022-April-1.pdf.

21. Agency for Healthcare Research and Quality. October 2021. Clinical Classifications Software Refined (CCSR). Healthcare Cost and Utilization Project (HCUP). Rockville, MD. https://www.hcup-us.ahrq.gov/toolssoftware/ccsr/ccs_refined.jsp.

22. Agency for Healthcare Research and Quality. July 2021. Immunocompromised State Diagnosis and Procedure Codes. https://qualityindicators.ahrq.gov/Downloads/Modules/PQI/V2021/TechSpecs/PQI_Appendix_C.pdf.

23. Agency for Healthcare Research and Quality. July 2021. Intermediate-Risk Immunocompromised State Diagnosis Codes. https://qualityindicators.ahrq.gov/Downloads/Modules/PDI/V2021/TechSpecs/PDI_Appendix_G.pdf.

24. Agency for Healthcare Research and Quality. July 2021. High-Risk Immunocompromised State Diagnosis and Procedure Codes. https://qualityindicators.ahrq.gov/Downloads/Modules/PDI/V2021/TechSpecs/PDI_Appendix_F.pdf.

25. Blauwkamp TA, Ho C, Seng H, Hollemon D, Blair L, Yao JD, Warren D, Russell P, Davis T, Hong DK. 2019. Evaluation of Karius plasma next generation sequencing of microbial cell-free DNA to detect and quantitate Cytomegalovirus, Epstein-Barr Virus, and BK Virus, abstr American Society for Microbiology (ASM) Microbe 2019, San Francisco, CA,

26. Fung M, Teraoka J, Lien K, Seng H, Parham A, Hollemon D, Hong DK, Blair L, Zompì S, Logan AC, Yao JD, Chin-Hong P. 2019. Use of the quantitative Karius® plasma next generation sequencing cell-free pathogen DNA test to detect and monitor Cytomegalovirus infection in allogeneic stem-cell transplant recipients, abstr 2019 TCT: Transplantation & Cellular Therapy Meetings of ASBMT and CIBMTR, Houston, TX,

27. Relman DA, Falkow S, Ramakrishnan L. 2020. Chapter 1: A Molecular Perspective of Microbial Pathogenicity, p 1-11. *In* Bennett JE, Bennett JE, Dolin R, Blaser MJ (ed), Mandell, Douglas, and Bennett’s principles and practice of infectious diseases, Ninth edition. ed, vol 1. Elsevier, Philadelphia, PA.

28. McKinney W. Data structures for statistical computing in python, abstr 9th Python in Science Conference, Austin, TX, June 28 - July 3, 2010.

29. Hunter J. 2007. Matplotlib: a 2D graphics environment. Computing in Science & Engineering. 9(3):90–95. doi:10.1109/MCSE.2007.55.

30. Waskom M, Botvinnik O, O’Kane D, Hobson P, Lukauskas S, Gemperline DC, Augspurger T, Halchenko Y, Cole JB, Warmenhoven J, de Ruiter J, Pye C, Hoyer S, Vanderplas J, Villalba S, Kunter G, Quintero E, Bachant P, Martin M, Meyer K, Miles A, Ram Y, Yarkoni T, Williams ML, Evans C, Fitzgerald C, B, Fonnesbeck C, Lee A, Qalieh A. 2017. mwaskom/seaborn: v0.8.1 (September 2017), September 3, 2017 ed doi:10.5281/zenodo.883859. Zenodo.

31. Ahmed AA, Rosen M, Hong DK, Dalai SC, Macintyre A, Blair L, Lindner M, Balakrishnan S, Bercovici S. Next-generation sequencing of pathogen cell-free DNA in plasma (Karius Test) reveals Nocardia species diversity in clinical infections, abstr ASM Microbe, San Francisco, CA, June 20–24, 2019.

32. Smollin M, Lindner M, Blair L, Arun A, Degner N, Equils O, de Vries CR, Dalai SC, MacIntyre A, Ahmed AA. Rapid, non-invasive detection of Legionella and Nocardia and resolution of species diversity in clinical infections in immunocompromised hosts using the Karius Test, a plasma-based microbial cell-free DNA sequencing test for pathogen identification, abstr 21st ICHS Symposium, Melbourne, Australia, February 17–19, 2021.

33. Graff K, Dominguez SR, Messacar K. 2021. Metagenomic Next-Generation Sequencing for Diagnosis of Pediatric Meningitis and Encephalitis: A Review. J Pediatric Infect Dis Soc 10:S78–S87.

34. Haslam DB. 2021. Future Applications of Metagenomic Next-Generation Sequencing for Infectious Diseases Diagnostics. J Pediatric Infect Dis Soc 10:S112–S117.

35. Huang AL, Hendren N, Carter S, Larsen C, Garg S, La Hoz R, Farr M. 2022. Biomarker-Based Assessment for Infectious Risk Before and After Heart Transplantation. Curr Heart Fail Rep doi:10.1007/s11897-022-00556-z.

36. Bergin SP, Chemaly R, Duttagupta R, Bigelow R, Dadwal S, Hill JA, Lee YJ, Haidar G, Luk A, Drelick A, Chin-Hong PV, Benamu E, Davis T, Wolf O, McClain M, Maziarz E, Madut D, Bedoya A, Gilstrap DL, Todd J, Barkauskas C, Spallone A, McDowell BJ, Small CB, Shariff D, Salsgiver E, Khawaja F, Papanicolaou GA, Spagnoletti J, Van Besien K, English M, Fung M, Rusell P, Ibrahimi S, Pandey S, Adams S, Liang W, Visweswaran A, Ho C, Nemirovich-Danchenko E, Braaten J, Sundermann L, Mughar M, Chavez R, Romano R, Montgomery S, Kumar S, Dalai SC, Cho Y, Ahmed AA, et al. 2022. PICKUP: Pneumonia in the ImmunoCompromised - use of the Karius Test® for the detection of Undiagnosed Pathogens abstr IDWeek 2022, Washington, D.C., October 20, 2022.

37. Farnaes L, Wilke J, Ryan Loker K, Bradley JS, Cannavino CR, Hong DK, Pong A, Foley J, Coufal NG. 2019. Community-acquired pneumonia in children: cell-free plasma sequencing for diagnosis and management. Diagn Microbiol Infect Dis 94:188–191.

38. Eichenberger EM, de Vries CR, Ruffin F, Sharma-Kuinkel B, Park L, Hong D, Scott ER, Blair L, Degner N, Hollemon DH, Blauwkamp TA, Ho C, Seng H, Shah P, Wanda L, Fowler VG, Ahmed AA. 2022. Microbial Cell-Free DNA Identifies Etiology of Bloodstream Infections, Persists Longer Than Conventional Blood Cultures, and Its Duration of Detection Is Associated With Metastatic Infection in Patients With Staphylococcus aureus and Gram-Negative Bacteremia. Clin Infect Dis 74:2020–2027.

39. To RK, Ramchandar N, Gupta A, Pong A, Cannavino C, Foley J, Farnaes L, Coufal NG. 2021. Use of Plasma Metagenomic Next-generation Sequencing for Pathogen Identification in Pediatric Endocarditis. Pediatr Infect Dis J 40:486–488.

40. de Melo Silva J, Pinheiro-Silva R, Dhyani A, Pontes GS. 2020. Cytomegalovirus and Epstein-Barr Infections: Prevalence and Impact on Patients with Hematological Diseases. Biomed Res Int 2020:1627824.

41. Hilt EE, Ferrieri P. 2022. Next Generation and Other Sequencing Technologies in Diagnostic Microbiology and Infectious Diseases. Genes 13:1566.

42. Vaca DJ, Dobler G, Fischer SF, Keller C, Konrad M, von Loewenich FD, Orenga S, Sapre SU, van Belkum A, Kempf VAJ. 2022. Contemporary diagnostics for medically relevant fastidious microorganisms belonging to the genera Anaplasma, Bartonella, Coxiella, Orientia, and Rickettsia. FEMS Microbiol Rev doi:10.1093/femsre/fuac013.

43. Haydour Q, Hage CA, Carmona EM, Epelbaum O, Evans SE, Gabe LM, Knox KS, Kolls JK, Wengenack NL, Prokop LJ, Limper AH, Murad MH. 2019. Diagnosis of Fungal Infections. A Systematic Review and Meta-Analysis Supporting American Thoracic Society Practice Guideline. Ann Am Thorac Soc 16:1179–1188.

44. Centers for Disease Control and Prevention (CDC). Tracking *Candida auris*. https://www.cdc.gov/fungal/candida-auris/tracking-c-auris.html. Accessed December 15, 2022.

45. Lockhart SR, Toda M, Benedict K, Caceres DH, Litvintseva AP. 2021. Endemic and Other Dimorphic Mycoses in The Americas. J Fungi (Basel) 7.

46. Centers for Disease Control and Prevention (CDC). C. gattii Infection Statistics. https://www.cdc.gov/fungal/diseases/cryptococcosis-gattii/statistics.html. Accessed June 5, 2022.

47. Yu VL, Plouffe JF, Pastoris MC, Stout JE, Schousboe M, Widmer A, Summersgill J, File T, Heath CM, Paterson DL, Chereshsky A. 2002. Distribution of Legionella species and serogroups isolated by culture in patients with sporadic community-acquired legionellosis: an international collaborative survey. J Infect Dis 186:127–8.

48. Helbig JH, Uldum SA, Bernander S, Luck PC, Wewalka G, Abraham B, Gaia V, Harrison TG. 2003. Clinical utility of urinary antigen detection for diagnosis of community-acquired, travel-associated, and nosocomial legionnaires’ disease. J Clin Microbiol 41:838–40.

49. Fields BS, Benson RF, Besser RE. 2002. Legionella and Legionnaires’ disease: 25 years of investigation. Clin Microbiol Rev 15:506–26.

50. Phin N, Parry-Ford F, Harrison T, Stagg HR, Zhang N, Kumar K, Lortholary O, Zumla A, Abubakar I. 2014. Epidemiology and clinical management of Legionnaires’ disease. Lancet Infect Dis 14:1011–21.

51. CLSI. 2018. Performance Standards for Susceptibility Testing of Mycobacteria, Nocardia spp., and Other Aerobic Actinomycetes. *In* (ed), CLSI Supplement M62. Clinical and Laboratory Standards Institute, Wayne, PA.

52. Sax H, Bloemberg G, Hasse B, Sommerstein R, Kohler P, Achermann Y, Rossle M, Falk V, Kuster SP, Bottger EC, Weber R. 2015. Prolonged Outbreak of Mycobacterium chimaera Infection After Open-Chest Heart Surgery. Clin Infect Dis 61:67–75.

53. Pollock NR, MacIntyre AT, Blauwkamp TA, Blair L, Ho C, Calderon R, Franke MF. 2021. Detection of Mycobacterium tuberculosis cell-free DNA to diagnose TB in pediatric and adult patients. Int J Tuberc Lung Dis 25:403–405.

54. Goldberg B, Sichtig H, Geyer C, Ledeboer N, Weinstock GM. 2015. Making the Leap from Research Laboratory to Clinic: Challenges and Opportunities for Next-Generation Sequencing in Infectious Disease Diagnostics. mBio 6:e01888–15.

55. Simner PJ, Miller S, Carroll KC. 2018. Understanding the Promises and Hurdles of Metagenomic Next-Generation Sequencing as a Diagnostic Tool for Infectious Diseases. Clin Infect Dis 66:778–788.

56. Wright WF, Simner PJ, Carroll KC, Auwaerter PG. 2022. Progress Report: Next-Generation Sequencing, Multiplex Polymerase Chain Reaction, and Broad-Range Molecular Assays as Diagnostic Tools for Fever of Unknown Origin Investigations in Adults. Clin Infect Dis 74:924–932.

57. Steensels D, Verhaegen J, Lagrou K. 2011. Matrix-assisted laser desorption ionization-time of flight mass spectrometry for the identification of bacteria and yeasts in a clinical microbiological laboratory: a review. Acta Clin Belg 66:267–73.

58. Camargo JF, Ahmed AA, Lindner MS, Morris MI, Anjan S, Anderson AD, Prado CE, Dalai SC, Martinez OV, Komanduri KV. 2019. Next-generation sequencing of microbial cell-free DNA for rapid noninvasive diagnosis of infectious diseases in immunocompromised hosts. F1000Res 8:1194.

59. Edward P, Handel AS. 2021. Metagenomic Next-Generation Sequencing for Infectious Disease Diagnosis: A Review of the Literature With a Focus on Pediatrics. J Pediatric Infect Dis Soc 10:S71–S77.

60. Fishman JA. 2022. Approach to the immunocompromised patient with fever and pulmonary infiltrates. *In* Blumberg EA, Bond S (ed), UptoDate, Waltham, MA.

61. Pierce KK. 2013. Chapter 40 - Immunocompromised Host, p 277–292. *In* Parsons PE, Wiener-Kronish JP (ed), Critical Care Secrets (Fifth Edition) doi:https://doi.org/10.1016/B978-0-323-08500-7.00041-2. Mosby.

62. Goggin KP, Gonzalez-Pena V, Inaba Y, Allison KJ, Hong DK, Ahmed AA, Hollemon D, Natarajan S, Mahmud O, Kuenzinger W, Youssef S, Brenner A, Maron G, Choi J, Rubnitz JE, Sun Y, Tang L, Wolf J, Gawad C. 2020. Evaluation of Plasma Microbial Cell-Free DNA Sequencing to Predict Bloodstream Infection in Pediatric Patients With Relapsed or Refractory Cancer. JAMA Oncol 6:552–556.

63. Solanky D, Ahmed AA, Fierer J, Golts E, Jones M, Mehta SR. 2022. Utility of plasma microbial cell-free DNA decay kinetics after aortic valve replacement for Bartonella endocarditis: case report. Front Trop Dis 3:842100.

64. Ahsouri N, Nieves D, Singh J, Arrieta A. 2022. Serial Microbial Cell-Free DNA Next Generation Sequencing (NGS) As A Means of Diagnosis and Monitoring of Clinical Response To Treatment Of Invasive Fungal Infections (IFI) In Immunocompromised Pediatric Patients, abstr IDWeek 2022, Washington, D.C., October 21, 2022.

65. Griffith BP, Goerlich CE, Singh AK, Rothblatt M, Lau CL, Shah A, Lorber M, Grazioli A, Saharia KK, Hong SN, Joseph SM, Ayares D, Mohiuddin MM. 2022. Genetically Modified Porcine-to-Human Cardiac Xenotransplantation. N Engl J Med 387:35–44.

66. Conville PS, Brown-Elliott BA, Witebsky FG. February 2019. Nocardia, Rhodococcus, Gordonia, Actinomadura, Streptomyces, and other aerobic actinomycetes. In Carroll KC, Pfaller MA, Landry ML, McAdam AJ, Patel R, Richter SS, Warnock DW (ed), Manual of Clinical Microbiology, 12th ed. ASM Press.

67. Caulfield AJ, Elvira, Brown-Elliott BA, Wallace J, Richard J., Wengenack NL. February 2019. Mycobacterium: laboratory characteristics of slowly growing mycobacteria other than Mycobacterium tuberculosis. In Carroll KC, Pfaller MA, Landry ML, McAdam AJ, Patel R, Richter SS, Warnock DW (ed), Manual of Clinical Microbiology, 12th ed. ASM Press.

68. Brown-Elliott BA, Wallace J, Richard J. February 2019. Mycobacterium: clinical and laboratory characteristics of rapidly growing mycobacteria. In Carroll KC, Pfaller MA, Landry ML, McAdam AJ, Patel R, Richter SS, Warnock DW (ed), Manual of Clinical Microbiology, 12th ed. ASM Press.

69. Centers for Disease Control and Prevention (CDC). 2022 National Notifiable Infectious Diseases. https://ndc.services.cdc.gov/search-results-year/. Accessed March 1, 2022.

70. Hoffmann K, Pawlowska J, Walther G, Wrzosek M, de Hoog GS, Benny GL, Kirk PM, Voigt K. 2013. The family structure of the Mucorales: a synoptic revision based on comprehensive multigene-genealogies. Persoonia 30:57–76.

